# Adaptive multi-model ensembles for improved epidemic projections and decision support

**DOI:** 10.64898/2026.06.26.26356648

**Authors:** Stefania Fiandrino, Daniela Paolotti, Clara Bay, Matteo Chinazzi, Jessica T. Davis, Samantha J. Bents, Amanda C. Perofsky, James Turtle, Pete Riley, Michal Ben-Nun, Sean M. Moore, Alex Perkins, Guido Espana, Ajitesh Srivastava, Majd Al Aawar, Shraddha Ramdas Bandekar, Kaiming Bi, Anass Bouchnita, Spencer J. Fox, Lauren Ancel Meyers, Srinivasan Venkatramanan, Przemyslaw Porebski, Aniruddha Adiga, Bryan Lewis, Madhav Marathe, Fardad Haghpanah, Eili Klein, Sara L. Loo, Sung-mok Jung, Claire P. Smith, Lucie Contamin, Harry Hochheiser, Erica C. Carcelén, Emily Howerton, Katriona Shea, Katie Yan, Michael C. Runge, Cecile Viboud, Carl A. B. Pearson, Shaun Truelove, Justin Lessler, Rebecca K. Borchering, Matthew Biggerstaff, Nicolò Gozzi, Alessandro Vespignani

## Abstract

In recent years, the use of multi-model ensemble projections in infectious disease modeling has become an established methodological approach to account for and integrate across uncertainties and structural differences present in individual models. However, the creation of long-term ensemble projections through these coordinated efforts is resource-intensive, demanding the input of multiple research teams and substantial computational power. This typically limits the ability to refine projections, update the selection of plausible epidemic trajectories, or expand the number of scenarios that can be assessed, even as new empirical data become available. To address this challenge, we define an adaptive ensemble approach that, analogously to a multi-model particle filtering method, dynamically selects individual model trajectories based on observed data throughout the epidemic projection period. We demonstrate the effectiveness of this methodology using the U.S. Flu Scenario Modeling Hub (SMH) projections for influenza hospitalizations in the United States during the 2023-2024 and 2024-2025 winter seasons. Our findings show that the adaptive ensemble yields improved predictive accuracy with respect to the original SMH ensemble projections across several scoring rules and geographical resolutions. Furthermore, the adaptive ensemble approach offers two additional applications: i) the dynamic assignment of posterior probabilities to epidemic scenarios, identifying the most plausible scenario, and representing how reality is captured by a combination of scenarios, and ii) the potential use for short-term forecasting. The adaptive ensemble approach is able to identify the most likely scenarios for the 2023-2024 and 2024-2025 U.S. influenza seasons, even in the early stages of the epidemic. It outperforms, retrospectively, a baseline model in short-term forecasting of influenza hospitalizations in the United States during the two seasons across various horizons and scoring rules, showing potential to contribute to real-time collaborative forecasting challenges such as CDC’s FluSight. The proposed approach offers an efficient or low-resource strategy to increase the impact of multi-model epidemic projections by providing real-time support to modeling teams, public health authorities, and decision-makers.

## 1. Introduction

Mathematical and computational models are now part of the toolkit for generating insights that support the management of epidemics and public health threats [1-6]. Drawing from best practices across various disciplines, such as weather forecasting [7-11], the field has increasingly focused on multi-model approaches, leading to the establishment of collaborative modeling hubs [2]. These hubs coordinate consortia of modeling teams, each contributing projections to predefined epidemic scenarios, which provide predictions over extended time horizons (e.g., an entire season) and are conditional on several scenario assumptions [3], or forecasting challenges [12, 14-28]. The outputs from individual models are thus aggregated to form ensemble projections that reflect a consensus among models, improving communication and interpretability. In the context of scenario projections, the ensemble reduces the risk of relying on a single model whose assumptions may not yield accurate projections, and shared scenario assumptions ensure that results across contributing models remain consistent with the same underlying scientific question. The ensemble potentially mitigates the limitations of single models, such as specific assumptions, parameter choices, and data integration challenges [3]. This is possible because model-specific uncertainties are generally independent, arising from differences in model design and assumptions, allowing the ensemble to mitigate these uncertainties rather than compound them. Although ensemble approaches have demonstrated, on average, superior and more stable predictive accuracy compared to individual models [29-30], they require substantial coordination, the contributions of several modeling teams, and considerable computational resources. These challenges are particularly burdensome in scenario projections. Given the anticipatory assumptions, these projections may require frequent updates to maintain relevance as new evidence about underlying phenomena or trends becomes available, to recalibrate models with new data to improve accuracy and realism, placing additional burdens on modeling teams and further complicating the management of hubs, which already demand extensive costs and resources [1].

In this paper, we introduce a methodological approach to efficiently and effectively update long-term epidemiological ensemble projections, thus improving their accuracy, by dynamically selecting individual model trajectories based on real-time observed data. The approach leverages newly available data to generate an *adaptive ensemble*, selecting a subset of trajectories across all models that best align with observed trends, similarly to a multi-model particle filtering or Sequential Monte Carlo (SMC)-like update [2], where candidate trajectories are filtered in real-time based on their consistency with observations. This method has the potential to provide practical enhancements over the existing ensemble approach used in collaborative modeling hubs, which pools together all individual long-term projections into a static ensemble that remains fixed and is never updated.

As a case study, we apply the proposed methodology to the 2023-2024 and 2024-2025 U.S. Flu Scenario Modeling Hub [3] projections for influenza hospitalizations in the United States at both national and subnational levels. Our results show that, based on the median performance across evaluation weeks, the adaptive ensemble achieves improved weighted interval scores and mean absolute errors with respect to the original static ensemble when evaluated on the remaining portion of the season not used for trajectory selection. Interestingly, the adaptive ensemble yields improved overall prediction performance even with minimal observed data for trajectory selection.

The proposed adaptive ensemble approach lends itself to two additional applications. First, by tracking the fraction of trajectories selected from each scenario, we can derive a proxy for the posterior probability of alternative epidemic scenarios. Considering the 2023-2024 and 2024-2025 influenza projections submitted to the U.S. Flu Scenario Modeling Hub, our findings indicate that the adaptive ensemble correctly identifies the scenarios, based on assumptions about influenza strain and vaccination coverage, that most closely align with reality, even in the early stages of the epidemic. Second, the methodology proves to be effective for short-term forecasting during this period. By dynamically selecting trajectories that best reproduce recent influenza hospitalization trends, the adaptive ensemble provides accurate predictions for the short-term evolution of epidemic curves. As a case study, we consider data from the 2023-2024 and 2024-2025 CDC’s FluSight forecasting challenge on influenza hospitalizations in the United States. The adaptive ensemble consistently outperforms the FluSight baseline model in both seasons. Compared to the FluSight ensemble, it shows competitive performance in the 2023-2024 season under certain scoring rules and parameter settings, but no improvements are observed in the 2024-2025 season. Overall, the adaptive ensemble shows the potential to contribute to the FluSight challenge as an additional model.

The proposed approach establishes a general method for dynamically refining epidemiological scenario analyses as new evidence becomes available. Furthermore, it provides a structured way to improve the impact of multi-model epidemic projections by offering real-time, adaptive support to modeling teams, public health authorities, and decision-makers.

## 2. Results

Updating long-term epidemic projections in real time requires an efficient mechanism to identify, among the large pool of trajectories generated before the season, those that are most consistent with emerging observations. The adaptive ensemble addresses this by dynamically selecting, at each epidemiological week, the best-performing trajectories from each contributing model based on their consistency with observed data over the calibration window. Figure 1 illustrates the adaptive ensemble approach: starting from the pool of trajectories, at each week we extend the calibration window by one week, select the best-performing trajectories (top X%) from each model based on a loss function, and generate the updated ensemble. The resulting adaptive ensemble is then evaluated against the static ensemble on the corresponding future observations, repeating this process at each step. Full methodological details are provided in the Methods section. In the main text, we present the results for the 2024-2025 influenza season and discuss key findings from the 2023-2024 season for comparison. Complete details and figures for the 2023-2024 season are provided in the Supplementary Information.

**Figure 1.**
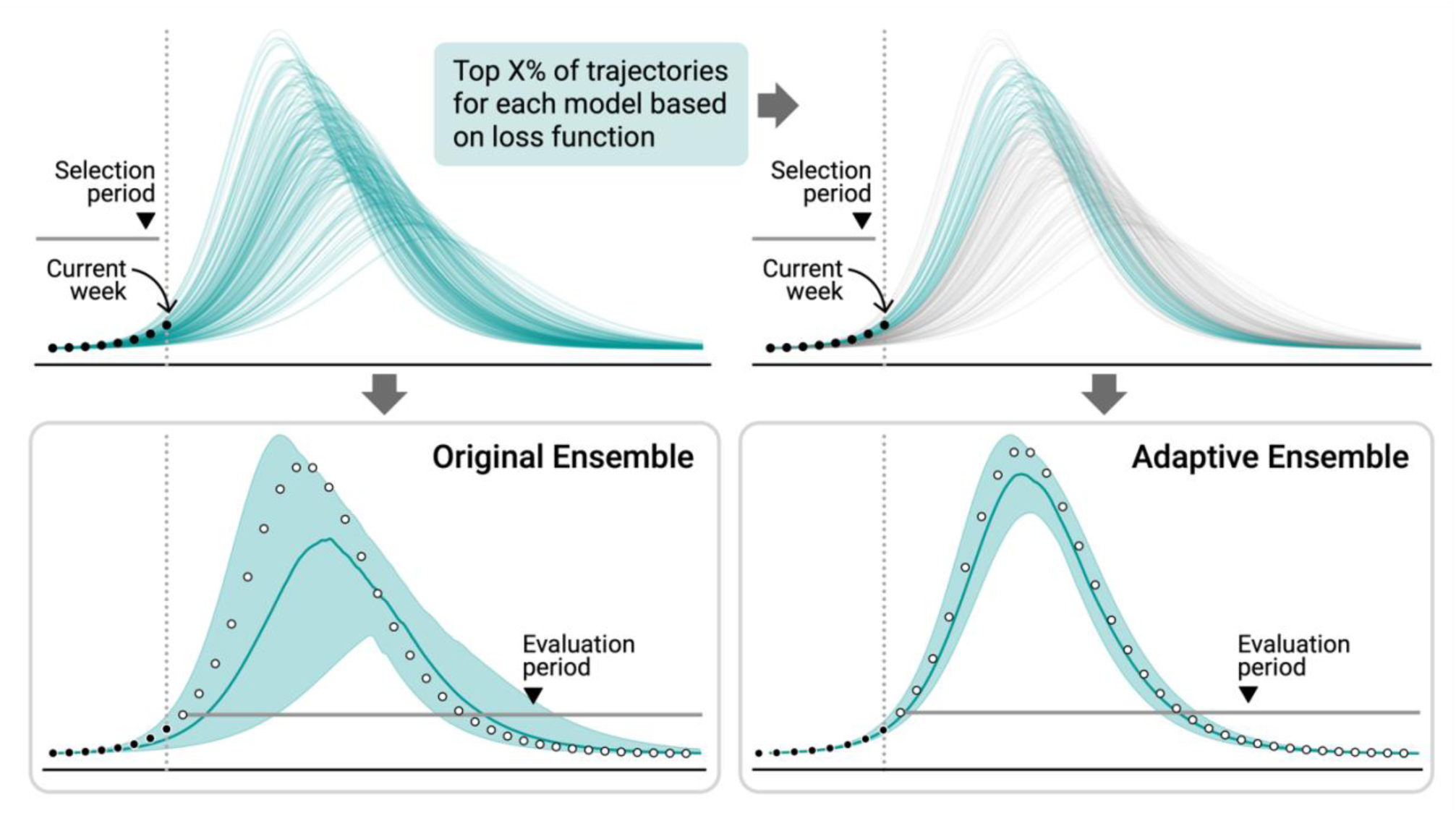
Conceptual overview of the Adaptive Ensemble. Starting from the full pool of trajectories generated prior to the season (top left), the top X% of trajectories from each contributing model are selected at each epidemiological week based on a loss function evaluated over a retrospective calibration window. Retained trajectories are used to construct the adaptive ensemble, and discarded trajectories are shown in grey (top right). The predictive performance of the adaptive ensemble (bottom right) is then evaluated against that of the original static ensemble (bottom left) on future observed data. In bottom panels, white open circles indicate future observed data points.

### 2.1 Evaluating adaptive ensemble for scenario modeling projections

To evaluate the effectiveness of the proposed adaptive ensemble for scenario modeling projections, we apply it to the U.S. 2023-2024 and 2024-2025 Flu Scenario Modeling Hub projections on weekly influenza hospitalizations (see Methods for a description of the dataset). We analyze the performance of the adaptive ensemble considering different percentages of selected trajectories, scoring criteria, and geographical resolutions. Specifically, in the following, we will compare it to the previously developed original ensemble^2^ approach, which is the ensemble computed by aggregating trajectories from all models and scenarios combined over the entire projection period, without any selection [4] (see Methods). Hereafter, the term *ensemble^2^* denotes that the aggregation is performed across both models and scenarios, rather than only across models, as in standard ensembling approaches [5]. In the Supplementary Information, we compare the adaptive ensemble to ensembles computed independently for each scenario, showing that the adaptive ensemble generally outperforms them, with the exception of Scenario E in 2024-2025 and Scenario A in 2023-2024 (Figs. S1-S2).

#### 2.1.1 Adaptive ensemble performance on Flu Scenario Modeling Hub projections

First, we focus on 2024-2025 scenario projections of weekly influenza hospitalizations at the national U.S. level. In each week, we generate the adaptive ensemble considering the weighted Mean Absolute Percentage Error (wMAPE) as loss function for trajectory selection (see Supplementary Information for loss function definitions), and thresholding at 5%, 15%, 25%, 50%, and 75% of trajectories selected. Performance is evaluated on the out-of-sample window (i.e., subsequent data points that were not used for trajectory selection) using the Weighted Interval Score (WIS) and the Mean Absolute Error (MAE) (see Methods).

In Fig. 2A, we show the distribution of the ratio between the WIS of the adaptive ensemble and of the original ensemble^2^ over all weeks in which the adaptive ensemble has been generated across different percentages of selected trajectories. Values below 1 indicate that the adaptive ensemble performs better than the original ensemble^2^ in terms of WIS, and values above 1 indicate worse performance. First, we observe that the adaptive ensemble outperforms the original ensemble^2^ in most cases. Indeed, considering all weekly updates, the median WIS ratio is smaller or equal to 1 for all the tested percentages. Second, we observe a substantial decrease in the variance of WIS ratios by increasing the number of selected trajectories. This is likely explained by the amount of filtering in each level of trajectory selection. When considering 75%, for example, a large proportion of trajectories is retained, thus leading to higher similarity with the original ensemble^2^. On the other hand, the 5% case, being more selective, shows a larger variance in performance with respect to the original ensemble^2^. In Fig. 2B, we repeat the analysis considering the MAE of the median for performance evaluation. Also in this case, the median MAE ratio is smaller than 1 for all percentages considered.

**Figure 2.**
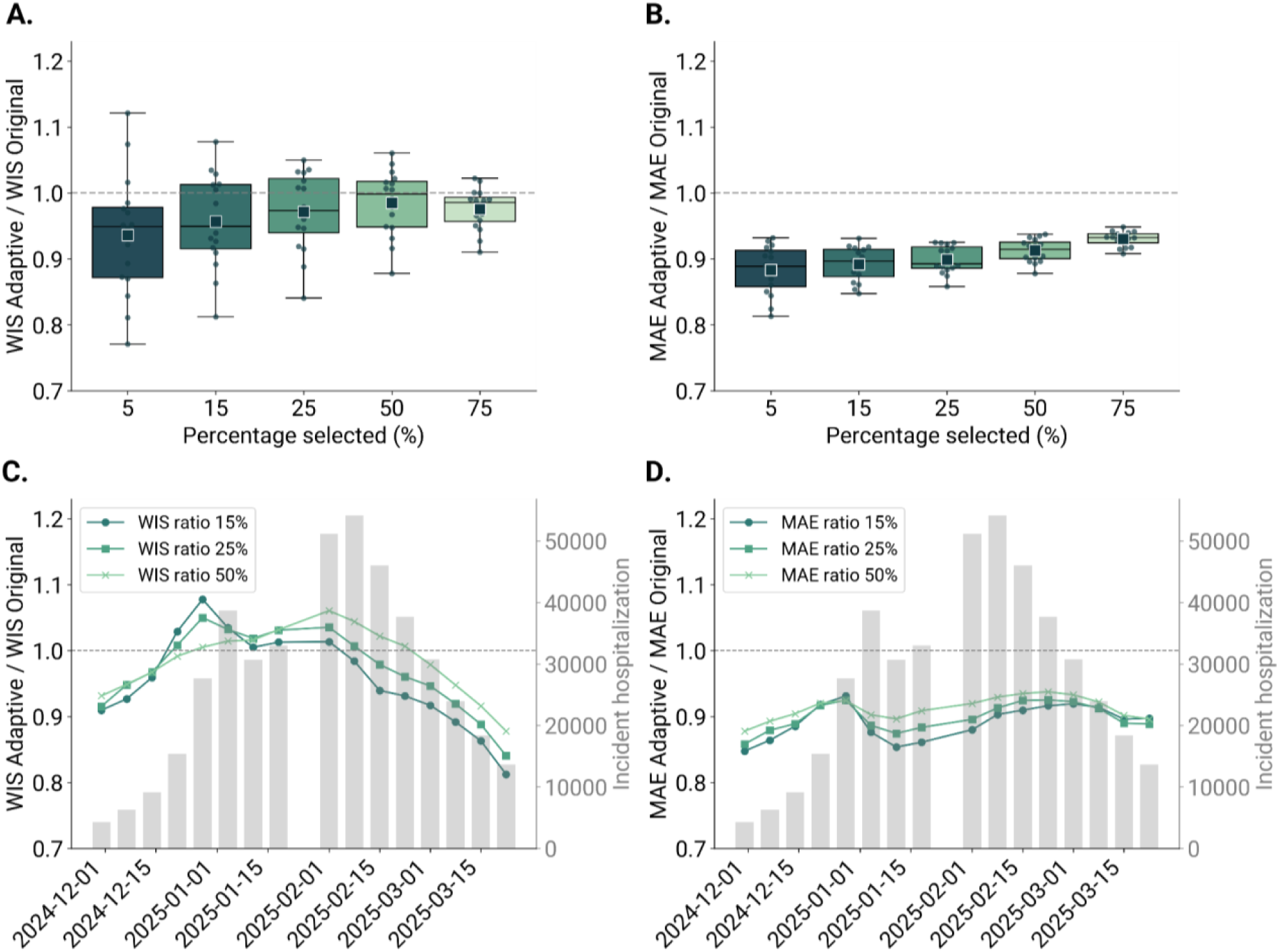
Performance of the adaptive ensemble model on US-level projections from the U.S. Flu Scenario Modeling Hub Round 1 [3], season 2024-2025. A) Ratio between the WIS of the adaptive ensemble and of the original ensemble^2^ across different percentages of trajectory selected and over all weeks. B) Ratio between the MAE of the median of the adaptive ensemble and of the original ensemble^2^ across different percentages of trajectory selected and over all weeks. A ratio below 1 indicates better performance of the adaptive ensemble with respect to the original ensemble^2^. Each boxplot is based on 16 data points, corresponding to the weeks in which the adaptive ensemble was generated. The overlaid swarmplot points correspond to each week and the square markers indicate the mean value. The boxplot boundaries represent the interquartile range (IQR) between the first quartile (Q1) and third quartile (Q3), and the line inside each box indicates the median. The whiskers extend to the furthest data point within 1.5 times the IQR from Q1 and Q3. C) Ratio between the WIS of the adaptive ensemble and of the original ensemble^2^ across selected percentages over time. D) Ratio between the MAE of the median of the adaptive ensemble and of the original ensemble^2^ across selected percentages over time. In the background, the reported weekly incident influenza hospitalizations are shown [5].

For easier interpretation of results, we report in Table 1 the median and interquartile (IQR) range of WIS and MAE ratios presented in Fig. 2 across different percentages of trajectories selected. The median WIS ratio values range from 0.95 (5%) to 1.00 (50%), indicating modest improvements compared to the original ensemble^2^. Interestingly, the upper bound IQR remains always less than or close to 1, with the 25% and 50% selection cases reaching 1.02, demonstrating a consistent performance improvement by the adaptive ensemble across most selection thresholds. In contrast, the median MAE ratio values range from 0.89 (5%) to 0.93 (75%), suggesting greater improvements over the original ensemble^2^ for specific threshold percentages compared to those observed for the WIS. In terms of median values, selecting 5% of trajectories yields the highest improvements for WIS and for MAE. Mean values are also reported as square markers, which are in line with median-based results (Figs. 2A-2B). In the Supplementary Information, we present results for the 2023-2024 season (Figs. S3A-S3B and Table S3), with similar findings: median WIS ratio values range from 0.96 (5%) to 0.99 (50% and 75%), and median MAE ratio values range from 0.79 (75%) to 1.14 (5%). We also repeat the analysis using root mean squared error instead of wMAPE as the loss function for trajectory selection and find analogous results (Figs. S4-S5).

**Table 1.** Performance summary of the adaptive ensemble model on US-level projections from the U.S. Flu Scenario Modeling Hub Round 1 [3], season 2024-2025. The table reports the median and interquartile range (IQR) of WIS and MAE of the median ratios WIS/MAE of the adaptive ensemble and of the original ensemble^2^ for each percentage of trajectory selected. In bold, the best WIS and MAE ratio median values out of the selected percentages. Values below 1 indicate better performance of the adaptive ensemble with respect to the original ensemble^2^.

| Percentage<br>% of selected trajectories | WIS ratio<br>Median (IQR) | MAE ratio<br>Median (IQR) |
| --- | --- | --- |
| 5% | **0.95 (0.87-0.98)** | **0.89 (0.85-0.91)** |
| 15% | 0.95 (0.92-1.01) | 0.89 (0.87-0.91) |
| 25% | 0.97 (0.94-1.02) | 0.89 (0.89-0.92) |
| 50% | 1.00 (0.95-1.02) | 0.91 (0.90-0.93) |
| 75% | 0.99 (0.96-0.99) | 0.93 (0.92-0.94) |

In Fig. 2C and Fig. 2D, we present the weekly WIS and MAE ratios, respectively, comparing the adaptive ensemble to the original ensemble^2^ over time, to evaluate performance without aggregating across the entire season. Specifically, the ratio value at time *t* indicates that the adaptive ensemble was computed by selecting trajectories based on data available up to time *t* and evaluated on all subsequent data points. In the case of WIS, we considered an average over all future data points. The background of the figures displays weekly incident influenza hospitalization in the U.S. during the 2024-2025 season [5]. Data for week 4, 2025 is missing because it was not released within the expected timeframe, and CDC’s National Health Safety Network (NHSN) data reporting resumed the following week. For visualization purposes, we focus on 15%, 25%, and 50% of selected trajectories, while results for the 5% and 75% cases are provided in the Supplementary Information (Fig. S6). For most weeks, the WIS and MAE ratio values remain below 1, indicating that the adaptive ensemble outperforms the original ensemble^2^. The adaptive ensemble does not show WIS improvements immediately before the peak of weekly influenza hospitalizations. Before and close to the peak, the WIS ratios increase, surpassing 1 for six and eight consecutive weeks in the 15% and 50% trajectories selected, respectively. Then, the adaptive ensemble again outperforms the ensemble^2^ in the declining phase of the epidemic. For MAE ratios, values consistently remain below 1 across all considered percentages. In the Supplementary Information (Figs. S3C-S3D, S7), we present results for the 2023-2024 season, showing a decline in performance (both WIS and MAE ratios increasing) during the upward trend in incident influenza hospitalizations. We observe differences in MAE distributions between the two seasons, with overall lower MAE and less weekly variation across the selection thresholds in 2024-2025 than in 2023-2024.

In the Supplementary Information, we have tested the robustness of these results through two sensitivity analyses (see Sections S5.5 and S5.6). First, we explore the effect of differences in the number of submitted trajectories across models (Fig. S8). Unlike the 2023-2024 season, in which each model contributed exactly 100 trajectories and was therefore equally represented, the 2024-2025 season allowed submissions of up to 300 trajectories, resulting in some variability in model representation. We therefore applied a bootstrapping approach to downsample trajectories to equal numbers across models, and the results were consistent with those obtained by retaining all trajectories. Second, we test whether the adaptive ensemble improvements hold when applied to the 65+ age group. We use the 65+ age group for the 2024-2025 season as a case study because a sufficient number of models for constructing an ensemble was available (5 models) [6]. The results (Fig. S9) show that the improvements of the adaptive ensemble with respect to the original ensemble hold at different tested percentages of selections and across both performance metrics. This demonstrates the potential of our approach for age-stratified analyses, when age-stratified projections are available.

We conducted an additional analysis to assess the stability of selected trajectories over time, evaluating how the pool of selected trajectories evolves across time steps. We compute the Jaccard Similarity Index [7], a measure of overlap between two sample sets, comparing: i) the similarity between the pool of trajectories selected at a specific time point and the one selected at the initial time step, and ii) the similarity between the pool of trajectories selected at a specific time point and the one selected at the previous time step. Methodological details and results are provided in the Supplementary Information (Section S4.2 and Figs. S10-S11). Across the board, our findings indicate high overall similarity as well as a gradual increase over time in the similarity of selected trajectories, suggesting that additional data points help stabilize the set of trajectories included in the adaptive ensemble.

We extend the performance analysis to projections of weekly incident influenza hospitalizations across U.S. states and territories during the 2024-2025 season. We exclude Puerto Rico from the analysis, as projections were available from only a single model. In this case, seven out of eight models submitted projections for all states and territories, whereas one model provided projections for only 39 out of 51 states and territories. In the main text, we present the results for the 25% of selected trajectories, and analogous results for the 15% and 50% cases are provided in the Supplementary Information (Figs. S12-S13). Fig. 3A presents the ratios of WIS between the adaptive ensemble and the original ensemble^2^ across all weeks, calculated for each U.S. state and territory and ordered by ascending median values. In more detail, 48 out of 51 U.S. states and territories feature a median WIS ratio lower than 1, indicating the improvements of the adaptive ensembling method also at higher spatial granularities. Fig. 3B presents analogous results considering the MAE of the median instead of the WIS. In the case of MAE ratios, the adaptive ensemble outperformed, in median terms, the original ensemble^2^ in 47 out of 51 U.S. states and territories. In the Supplementary Information, we present the state-level analysis for the 2023-2024 season (Figs. S14-S16), which reveals consistent results across different percentage configurations: at least 38 out of 51 states had median WIS ratios below 1, and 40 out of 51 states had median MAE ratios below 1. However, the 2023-2024 season presents greater heterogeneity in ensemble performance across states compared to the 2024-2025 season, with some states showing substantially higher ratios, particularly in the MAE.

**Figure 3.**
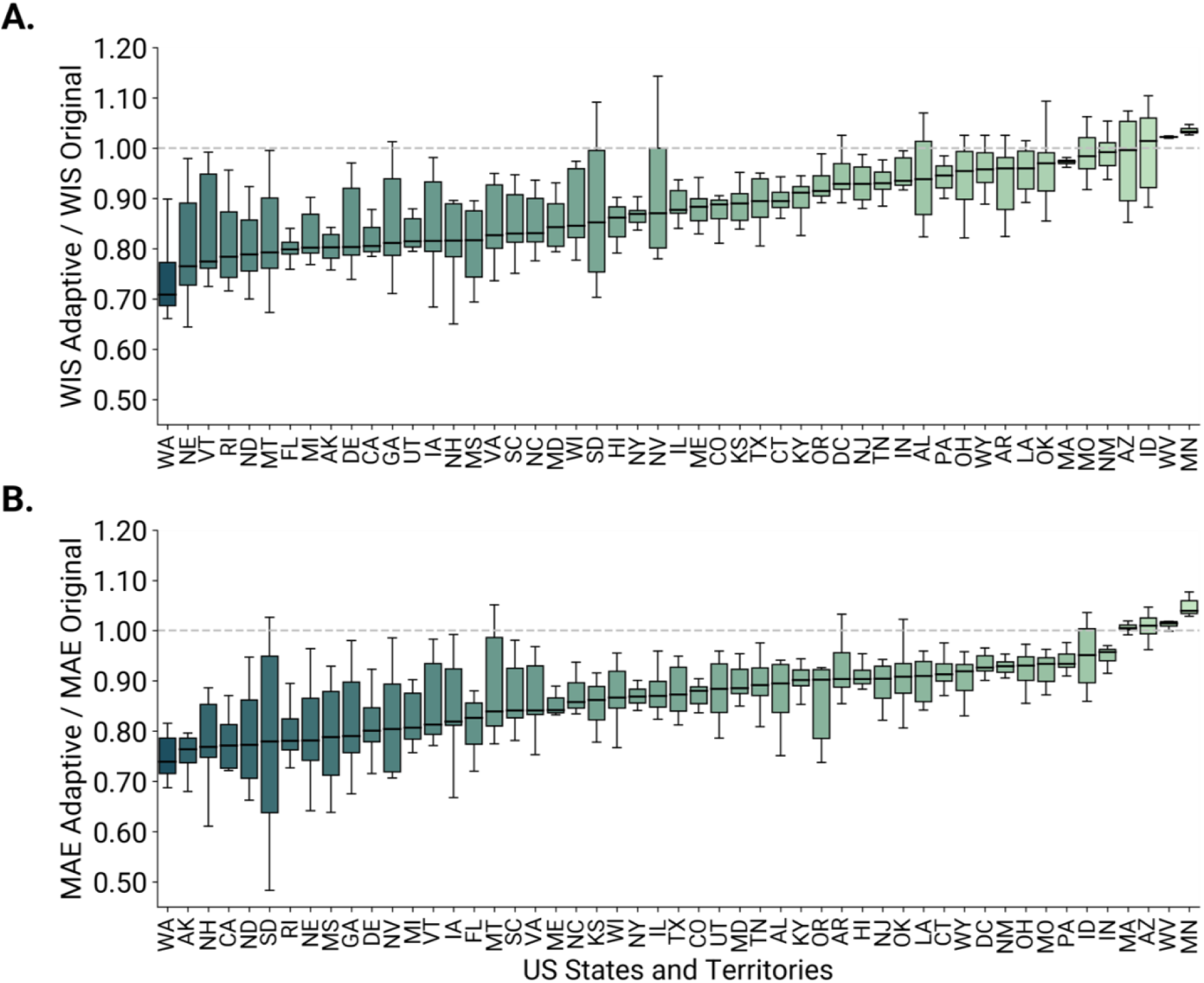
Performance of adaptive ensemble (25% selected trajectories) on projections for U.S. states and territories from the U.S. Flu Scenario Modeling Hub Round 1 [3], season 2024-2025. A) For each U.S. state and territory, we show the ratio between the WIS of the adaptive ensemble and of the original ensemble^2^ for the 25% percentage of trajectory selected and over all weeks. B) Ratio between the MAE of the median of the adaptive ensemble and of the original ensemble^2^ for the 25% percentage of trajectory selected and over all weeks. A ratio below 1 indicates better performance of the adaptive ensemble with respect to the original ensemble^2^. Each boxplot is based on 16 data points, corresponding to the weeks in which the adaptive ensemble was generated. The boxplot boundaries represent the interquartile range (IQR) between the first quartile (Q1) and third quartile (Q3), and the line inside each box indicates the median. The whiskers extend to the furthest data point within 1.5 times the IQR from Q1 and Q3.

#### 2.1.2 Exploring posterior probability of alternative epidemic scenarios

In the following, we present the posterior probability estimates derived from the adaptive ensemble and discuss their correspondence with the epidemiological dynamics and vaccination coverage observed during the study seasons. For each time step, we estimate the posterior probability of each scenario by tracking the fraction of trajectories selected from that scenario, providing a data-driven measure of scenario plausibility (see Methods). Fig. 4 shows the weekly fraction of trajectories originating from each scenario selected by the adaptive ensemble, considering the 25% of trajectories selected for national U.S. level projections for the 2024-2025 season. As explained in the Methods, these fractions can be considered an estimate of the posterior probability of different scenarios based on the surveillance data available. We refer to the Supplementary Information (Fig. S18) for the analogous analysis considering 15% and 50% of the trajectories selected, for the analysis of the 2023-2024 season (Figs. S20-S21), and for the analysis using age-stratified projections for 65+ years old (Fig. S22).

**Figure 4.**
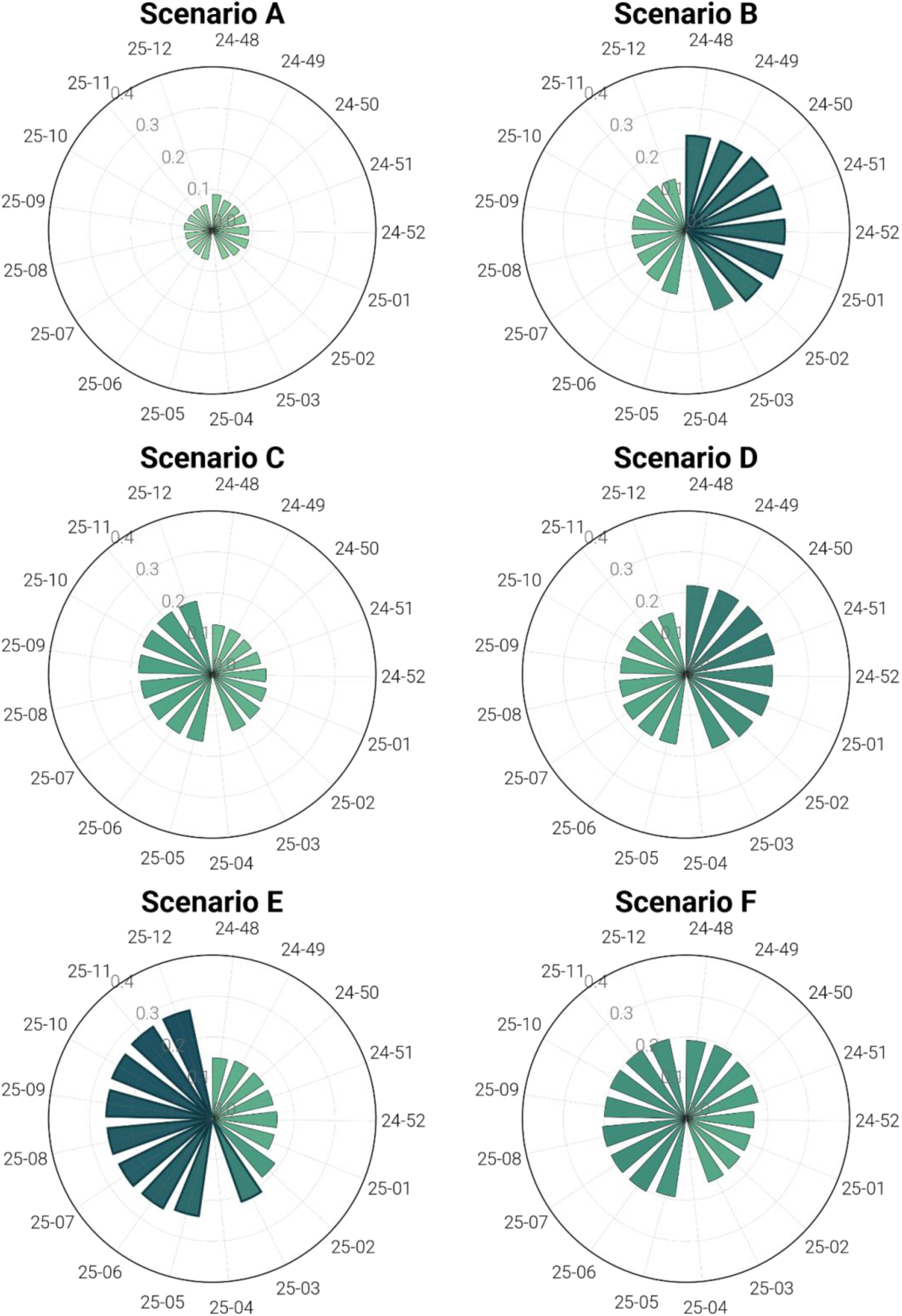
Exploring scenarios posterior probability for 2024-2025 U.S. influenza season, data from U.S. Flu Scenario Modeling Hub Round [3]. The plot shows the weekly fraction of trajectories originating from each scenario selected by the adaptive ensemble (top 25% trajectories selected). Scenarios assuming A/H1N1 dominance in the 2024-2025 season are shown on the right side (B, D, F), and those assuming A/H3N2 dominance are on the left side (A, C, E). Scenarios in the first row assume 20% higher vaccination coverage in all age groups and jurisdictions with respect to 2021/2022 coverage, scenarios in the second row assume a business-as-usual coverage, and scenarios in the third row assume a 20% lower coverage. The labels around the circumference denote epidemiological weeks in year-week format (YY-WW). The color gradient reflects the magnitude of the posterior probability each week, with darker colors indicating higher probabilities and lighter colors indicating lower probabilities.

At the start of the winter season after surpassing the epidemic baseline threshold (late November 2024), scenarios B, D, and F associated with A/H1N1 dominance and varying vaccination coverage levels (higher-than-usual, business-as-usual, and lower-than-usual, respectively), emerge as more plausible than those assuming A/H3N2 dominance. From week 3 of 2025 onward, scenario E, assuming A/H3N2 dominance, becomes the leading scenario followed by scenario F, which assumes A/H1N1 dominance with similarly lower vaccination coverage with respect to the reference season, and then by scenarios C and D (either A/H3N2 or A/H1N1 dominance with business-as-usual coverage). These findings are consistent with the epidemiological characteristics of the 2024-2025 season, in which (i) A/H1N1 and A/H3N2 subtypes co-circulated in the U.S. with comparable cumulative percent positivity [8], and (ii) national vaccination coverage decreased among children yet remained stable among adults [39, 40], reflecting a reality intermediate between the lower-than-usual and business-as-usual coverage scenarios. To support this interpretation, we provide in the Supplementary Information a comparison between the observed cumulative subtype proportions and the posterior scenario contributions over time, for both seasons (Figs. S17, S19). Further details on the actual characteristics of the 2024-2025 season, including the dominant subtype and vaccine uptake, are provided in the Discussion, along with a comparison to the 2023-2024 season.

### 2.2 Evaluating adaptive ensemble for short-term forecasting

We assess the performance of the adaptive ensemble approach for short-term forecasting using data from the 2024-2025 season of the FluSight forecasting challenge [1] (see Methods for details on FluSight). Specifically, we generate retrospective short-term forecasts of weekly incident influenza hospitalizations at the national U.S. level, following the adaptive ensemble methodology for short-term forecasting outlined in the Methods. These forecasts were generated from November 30, 2024, to May 31, 2025, aligning with the timing of the weekly submissions in the FluSight forecasting challenge. The forecasts produced by the adaptive ensemble approach are then compared against the FluSight baseline model, the neutral benchmark for evaluation, and the FluSight ensemble model, which combines forecasts from all participating models that are designated for inclusion in the ensemble.

Fig. 5A presents boxplots summarizing, for different percentages of selected trajectories, the distribution of the ratios between the WIS of the adaptive ensemble forecasts and those of the FluSight baseline and ensemble models, averaged across all forecasting horizons for each forecasting round (each round corresponds to a week). A ratio below 1 indicates that the adaptive ensemble outperformed the reference model in terms of WIS, whereas a ratio above 1 indicates lower performance. Similarly, Fig. 5B presents the same information for the MAE as scoring rule. When compared to the FluSight baseline model, the median WIS ratios are below 1 for all percentages of selected trajectories, indicating that the adaptive ensemble consistently outperforms the baseline in terms of WIS. In contrast, when compared to the FluSight ensemble model, the adaptive ensemble shows worse median performance (WIS ratio above 1) across all percentages of selected trajectories. When performance is measured using the MAE of the median, the results are analogous. Table 2 presents the median and interquartile ranges of WIS and MAE ratios for all percentages of trajectories selected, relative to the FluSight baseline and ensemble models. Across the board, we observe improved performance when lower percentages of trajectories are selected (i.e., trajectory inclusion is more selective). The relevant finding is that the adaptive ensemble remains competitive despite using a fraction of the resources.

**Figure 5.**
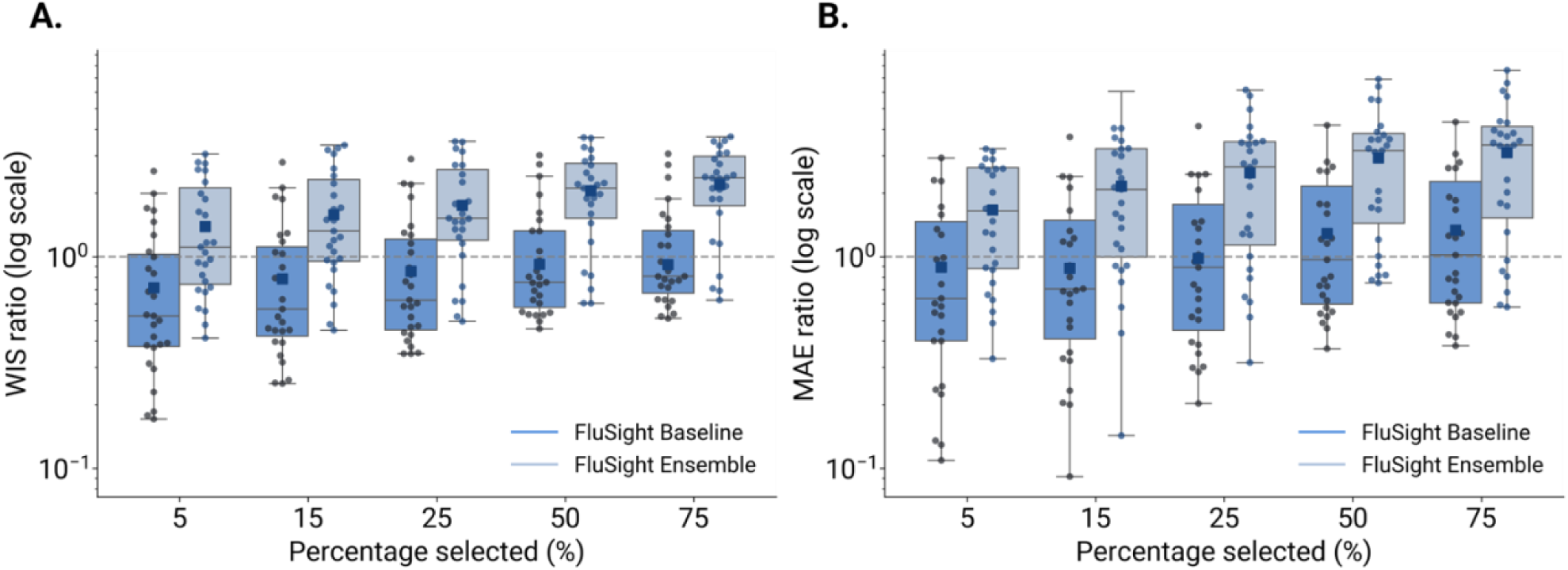
Performance of the adaptive ensemble model in short-term forecasting, data from FluSight season 2024-2025 [1]. A) Ratio between the WIS of the adaptive ensemble and FluSight Baseline or FluSight Ensemble model across different percentages of trajectory selected and over all weeks. B) Ratio between the MAE of the median of the adaptive ensemble and FluSight Baseline or FluSight Ensemble model across different percentages of trajectory selected and over all weeks. A ratio below 1 indicates better performance of the adaptive ensemble with respect to the FluSight Baseline or FluSight Ensemble. Each boxplot is based on 27 data points, corresponding to the weeks in which the adaptive ensemble was generated. The overlaid swarmplot points correspond to each week and the square markers indicate the mean value. The boxplot boundaries represent the interquartile range (IQR) between the first quartile (Q1) and third quartile (Q3), and the line inside each box indicates the median. The whiskers extend to the furthest data point within 1.5 times the IQR from Q1 and Q3.

**Table 2:** Performance summary of adaptive ensemble in short-term forecasting, data from FluSight season 2024-2025 [1]. The table reports the median and interquartile ranges of WIS and MAE ratios for each percentage of trajectory selected. In bold, the best WIS and MAE ratio median values out of the selected percentages; in italics, WIS and MAE ratio median values higher than 1. Values below 1 indicate better performance of the adaptive ensemble with respect to the reference (FluSight Baseline or Ensemble).

| Percentage<br>% of<br>selected<br>trajectories | WIS<br>(Adaptive / FluSight<br>Bas.)<br>Median (50% CI) | WIS<br>(Adaptive / FluSight<br>Ens.)<br>Median (50% CI) | MAE<br>(Adaptive / FluSight<br>Bas.)<br>Median (50% CI) | MAE<br>(Adaptive / FluSight<br>Ens.)<br>Median (50% CI) |
| --- | --- | --- | --- | --- |
| 5% | **0.53 (0.38-1.03)** | **1.11 (0.74-2.12)** | **0.64 (0.40-1.46)** | **1.65 (0.88-2.64)** |
| 15% | 0.54 (0.42-1.11) | 1.32 (0.95-2.31) | 0.71 (0.41-1.48) | 2.08 (1.00-3.23) |
| 25% | 0.62 (0.45-1.21) | 1.52 (1.20-2.58) | 0.89 (0.45-1.77) | 2.65 (1.14-3.48) |
| 50% | 0.76 (0.58-1.32) | 2.11 (1.52-2.77) | 0.97 (0.60-2.15) | 3.17 (1.44-3.82) |
| 75% | 0.81 (0.67-1.33) | 2.35 (1.74-2.98) | 1.02 (0.60-2.27) | 3.37 (1.52-4.12) |

In the Supplementary Information, we present the equivalent analysis stratified by forecasting horizon (Figs. S23-S24) and visualize the 4-week-ahead forecasts generated by the adaptive ensemble for different percentages of selected trajectories (Figs. S25-S26). The results are consistent, showing that in general, the adaptive ensemble achieves better performance in median terms at each horizon when compared to the FluSight baseline, and it maintains lower performance than the FluSight ensemble. In the Supplementary Information, we also compare the adaptive ensemble to the FluSight baseline and ensemble models in terms of coverage probability (Figs. S27-S28). The results indicate that the adaptive ensemble tends to exhibit overconfidence across all forecasting horizons, with prediction coverages generally lower than the nominal ones. In contrast, the FluSight ensemble shows prediction coverages that are more closely aligned with the nominal values. More specifically, when assessing coverage mismatch for 50% and 90% coverage (Table S4), the FluSight Ensemble demonstrates the smallest absolute deviations in all cases.

In the Supplementary Information, we present the full results of the retrospective short-term forecasting analysis for the 2023-2024 season, including additional details on the time period, the number of submission rounds, and participating teams in the FluSight challenge. In this case study, we observe similar performance in terms of median WIS, and no improvements relative to the FluSight ensemble, except for 5% selection case. Interestingly, MAE ratios consistently fall below 1 across all percentages of trajectories included in the adaptive ensemble, when compared to the FluSight baseline and the FluSight ensemble, indicating improved performance with respect to both according to this scoring rule (Figs. S29-S31). Additionally, the adaptive ensemble exhibits underconfidence, in contrast to the behavior observed for the 2024-2025 season (Figs. S32-S35 and Tab. S6).

## 3. Discussion

In this study, we developed a methodological approach for epidemic projections that leverages adaptive multi-model ensembles. The approach dynamically selects trajectories generated at the start of the season under predefined epidemiological scenarios, with selection progressively informed by observed data as it becomes available. We evaluated the proposed methodology using scenario projections submitted to the U.S. Flu Scenario Modeling Hub for the 2023-2024 and 2024-2025 influenza seasons. We showed that, on average, the proposed adaptive ensemble outperforms a static ensemble based on all trajectories across different scoring rules and influenza seasons. This pattern also holds at higher spatial granularities. Nonetheless, we acknowledge a reduction in adaptive ensemble performance in complex phases of the season, specifically at the onset and during the peak in the 2023-2024 season, and just before the epidemic peak in the 2024-2025 season. Early in the season, this is potentially linked to the scarcity of data available for trajectory selection. Around the epidemic peak, performance may decline due to the increased complexity of the epidemic phase, where individual models tend to struggle, impacting the overall performance of the ensemble. This limitation is also reflected in some state-level results, particularly in seasons characterized by atypical epidemic dynamics, where no individual model adequately captures key epidemic features (e.g., peak timing, peak magnitude); in these cases, selecting trajectories based on past fit may reduce performance compared to the static ensemble, which benefits from retaining a broader range of trajectories. The performance of the adaptive ensemble varies across evaluation metrics, as each captures distinct aspects of predictive accuracy: improvements in WIS reflect better calibration and sharpness of the full predictive distribution, whereas improvements in MAE indicate more accurate point forecasts. These differences reflect how the filtering procedure interacts with performance metrics. The MAE evaluates only the point forecast, similarly to the metric used for filtering: when the top-performing trajectories on past data also capture future dynamics, filtering stabilizes median predictions; otherwise, retaining the full trajectory set may be preferable. The WIS evaluates the full predictive distribution, and the filtering is driven by a point accuracy metric, which does not explicitly optimize for calibration and sharpness. When the pool of trajectories adequately represents future dynamics, filtering produces better-calibrated intervals than the static ensemble; conversely, if only a few trajectories capture future behavior, the filtered set may fail to represent future uncertainty, leading to poorly calibrated intervals. In this situation, the static ensemble benefits from a broader and more diverse trajectory pool. In the 2024-2025 season, these dynamics are reflected in the observed results. The filtered trajectories were clustered around the future epidemic trend, stabilizing the median prediction early and possibly explaining why MAE appears less dispersed even at very low selection percentages. The peak in WIS around the 50% threshold may likely reflect a transitional regime in which the filtered ensemble produces intervals that are neither narrow enough to benefit from sharpness nor diverse enough to ensure calibration, whereas both smaller and larger selection percentages avoid this unfavorable trade-off.

We showed that the proposed methodology lends itself to additional interesting applications. First, the adaptive ensemble naturally provides estimates for the posterior probability of individual scenarios based on observed data. Although identifying the most plausible scenario remains challenging even retrospectively, the posterior scenario analysis generally aligned with observed epidemiological patterns during the 2023-2024 and 2024-2025 influenza seasons. In the 2024-2025 season, most influenza viruses detected in the U.S. were influenza A, with A/H1N1 and A/H3N2 subtypes co-circulating with comparable cumulative positivity across different weeks [8]. Early in the season, the adaptive ensemble most frequently selected trajectories from scenarios assuming A/H1N1 dominance, and scenario E, assuming A/H3N2 dominance, also maintained a non-negligible posterior probability. From late February onward, scenario E (A/H3N2) became the most likely, followed by scenario F (A/H1N1), consistent with the co-circulation of both subtypes during the 2024-2025 influenza season. Determining which vaccination coverage scenario best matches reality is more complex. During 2024-2025, national vaccination coverage decreased among children aged 6 months-17 years (49.2% vs. 55.5% in 2022-2023 [9]) and remained stable among adults aged 18+ years (46.7% vs. 46.2% [0]). This supports identifying scenarios assuming reduced coverage (E and F) as the most likely, which were the ones most frequently chosen by the adaptive ensemble, followed closely by business-as-usual scenarios (C and D). In contrast, the 2023-2024 season was characterized by early and sustained dominance of influenza A/H1N1 [2]. In over 85% of the 25 analyzed weeks, the adaptive ensemble most frequently selected trajectories from A/H1N1-dominant scenarios, consistent with surveillance data [3]. Vaccination coverage during this season declined slightly among children (54% vs. 57% in 2021-2022 [9]) but increased modestly among adults (48% vs. 45% [0]), with a notable 11% relative increase among those aged 65+. These vaccination trends suggest an overall vaccination coverage closer to or slightly higher with respect to the reference season. This observation, combined with an A/H1N1-dominant season, points to scenarios B and D as the most representative of the actual season. Notably, these were also the scenarios most frequently selected by the adaptive ensemble. Some of these assumptions can be directly verified as the season progresses, but the results highlight the potential of the proposed approach in identifying the most plausible scenarios. This suggests potentially interesting applications, particularly for assumptions that are more difficult to validate directly, such as the effectiveness of interventions. In this context, this approach can inform the development of dashboards, offering policymakers real-time insights into how the evolving epidemiological situation aligns with expectations and supporting the iterative design of multi-round scenarios.

Second, the proposed methodology can be directly applied to short-term forecasting. A retrospective analysis using data and forecasts from the 2023-2024 and 2024-2025 FluSight challenge shows that the adaptive ensemble outperforms the FluSight baseline model but underperforms relative to the FluSight ensemble. This comparison, however, is structurally asymmetric. The FluSight ensemble aggregates forecasts from up to 50 independently maintained models, each ingesting new surveillance data weekly and many specifically optimized for short-term forecasting through repeated participation in the challenge. It represents a substantial distributed effort involving dozens of research teams operating in real time. The adaptive ensemble, by contrast, draws from eight models whose trajectories were generated once before the season for the purpose of scenario projections, a fundamentally different task. Given these differences, the competitive performance achieved with a fraction of the resources is notable. In 2024-2025, the underperformance compared to FluSight ensemble may also be influenced by the atypically severe influenza season, characterized by a markedly high peak magnitude, which reached levels not seen since 2017-2018 [44-45]. A severe season is likely underrepresented in the available scenario trajectories, which may not capture comparable dynamics, limiting the adaptive ensemble’s ability to select trajectories that align with observations. Nonetheless, during the more moderately severe 2023-2024 season, the adaptive ensemble demonstrated competitive performance compared to the FluSight ensemble for specific scoring rules and forecasting horizons. Overall, this demonstrates the potential for reusing long-term epidemic trajectories from scenario modeling to generate short-term forecasts that can strengthen the overall hub ensemble by submitting the adaptive ensemble as a contributing forecasting model.

The present work comes with the usual limitations of retrospective analysis. Although we used data as they would have been available during the 2023-2024 and 2024-2025 influenza seasons, this retrospective analysis does not fully address the challenges of real-time infectious disease modeling, such as reporting delays and operational challenges [6]. To test the impact of such limitations and to evaluate our approach in realistic contexts, we actively contribute adaptive ensemble short-term forecasts to collaborative forecasting hubs in both the U.S. and Europe, with results consistently indicating competitive performance in fully prospective settings [47-50].

Among the different percentages of trajectories selected, no single percentage consistently performed best across scoring rules and predictive tasks. This work offers a starting point and provides some intuition on the overall behavior of performance across a fixed range of percentages: results suggest a decrease in the variance of the WIS ratios as the number of retained trajectories increases. When more trajectories are retained, the adaptive ensemble performs similarly to the original static ensemble^2^, yielding little or no gain. Conversely, higher selectivity (i.e., retaining fewer trajectories) allows for larger improvements, but at the cost of increased fragility and more likely inconsistent results across rounds. However, future research could explore the trade-offs between selecting a specific set of trajectories and the information lost by excluding others. These trade-offs may vary depending on the predictive task (e.g., long-term scenario modeling versus short-term forecasting), the number and independence of contributing models, the specific set of scenarios considered, and the specific phase of the current epidemic. Future work could also investigate adaptive, data-driven strategies (e.g., reinforcement learning) to dynamically adjust the selection threshold over time. Also, future research could explore alternative strategies for selecting the pool of trajectories for the adaptive ensemble, beyond relying uniquely on individual trajectory performance. The *optimal* set of trajectories may not consist of those with the best standalone performance. Advanced techniques, such as machine learning-based optimization or Shapley value-based approaches [1], could provide effective and interpretable solutions for this objective. Alternatively, the trajectory selection procedure can be extended to more complex strategies that adjust the prior on each scenario and model according to empirical data or expert inputs. Finally, a key goal of scenario projections is the comparison of disease outcomes between scenarios [3] (e.g., project the number of hospitalizations averted by vaccination vs a counterfactual non-intervention scenario). When counterfactual non-intervention (i.e., no vaccination) is a possibility, trajectory selection could potentially be applied towards this goal, as more observations become available. Although the adaptive ensemble approach shows potential to offer insights into the most likely scenarios, this has been demonstrated using only two seasons and one disease. Further evaluation across additional seasons and diseases could strengthen these findings.

It is important to note that the adaptive ensemble approach is not agnostic to the purpose of scenario design [3]. Scenarios that are not explicitly designed to capture uncertainty, such as those contrasting alternative interventions or exploring counterfactuals, are not intended to represent the full range of plausible outcomes. As a result, it may not be possible to assign meaningful priors to these scenarios or to ensure that posterior analyses yield insights aligned with real-world conditions. In other words, understanding the objectives and structure of the scenario design is essential for determining the applicability of adaptive ensemble methods, particularly for posterior scenario distributions and short-term forecasting.

Another potential limitation of the adaptive methodology lies in the trade-off between the accuracy gains that could be achieved by fully re-running models with updated data and the practical challenges of doing so in real time. In future seasons, this trade-off could be explicitly evaluated by requesting more frequent scenario updates from participating teams and comparing the performance of these updated ensembles to that of the adaptive approach presented here. Such comparisons would help to contrast potential improvements in accuracy versus the associated computational and organizational costs.

An open question concerns how the performance of the adaptive ensemble scales with the number of contributing models, and whether accuracy stabilizes beyond a certain ensemble size. An additional aspect, which could be explored further, is the extent to which specific models are consistently selected within the adaptive ensemble. An alternative approach could be to select trajectories across all models, rather than selecting the same proportion from each model separately. The frequency with which trajectories from a given model are selected may provide insight into model performance, reflecting factors such as structural assumptions, calibration, and underlying epidemiological hypotheses. The adaptive ensemble approach can therefore be extended to estimate model posterior probabilities, enabling a more formal assessment of each model’s contribution over time. This direction may offer a complementary perspective on evaluating epidemic scenario projections within a trajectory-based framework, and provide insights to guide the development and refinement of individual models.

In conclusion, we proposed a novel methodological approach that adaptively improves long-term epidemic ensemble projections, with the potential to offer reliable insights into the most likely scenarios. The method also demonstrates the possibility of reusing scenario modeling epidemic trajectories to contribute to short-term forecasting tasks. Although additional research and validation may further strengthen this approach, the current methodology supports informed decision-making under uncertainty, to support both epidemic modelers and stakeholders.

## 4. Materials and Methods

In the following, we describe the data and the methodology used to develop and evaluate the adaptive ensemble. We then detail its application to both long-term scenario projections and short-term forecasts, along with the approach used to estimate posterior probabilities of epidemic scenarios.

### 4.1 Data

#### 4.1.1 Flu Scenario Modeling Hub scenario projections

We consider long-term projections submitted to the U.S. Flu Scenario Modeling Hub round (i.e., a coordinated modeling exercise under shared assumptions) of seasons 2023-2024 and 2024-2025 [3]. These projections were contributed by multiple modeling teams through the hub’s GitHub repository [2]. Prior to each influenza season, modeling teams participating in the hub were asked to submit weekly incident influenza hospitalization projections from September 3, 2023, to June 1, 2024, for the 2023-2024 season, and from August 11, 2024, to June 7, 2025, for the 2024-2025 season, at both national and subnational levels in the United States. Each modeling team contributed model projections for each of the six scenarios reported in Fig. 6. The scenario design [3], defined by the Scenario Modeling Hub, considered two main axes. One scenario axis accounted for different dominant virus subtypes, with the season being dominated either by influenza A/H3N2 or A/H1N1. The other scenario axis considered varying levels of vaccination coverage: 20% higher, the same (“business as usual”), and 20% lower coverage across all ages and jurisdictions compared to a reference season, specifically 2021-2022 for the 2023-2024 projections, and 2022-2023 for the 2024-2025 ones (Fig. 6). The reference seasons were selected based on the availability of coverage data at the start of the season and serve only to define baseline vaccination coverage values [39, 40]. This leads to six distinct scenarios (labeled from A to F), each representing a combination of dominant virus subtype and vaccination coverage assumptions.

**Figure 6.**
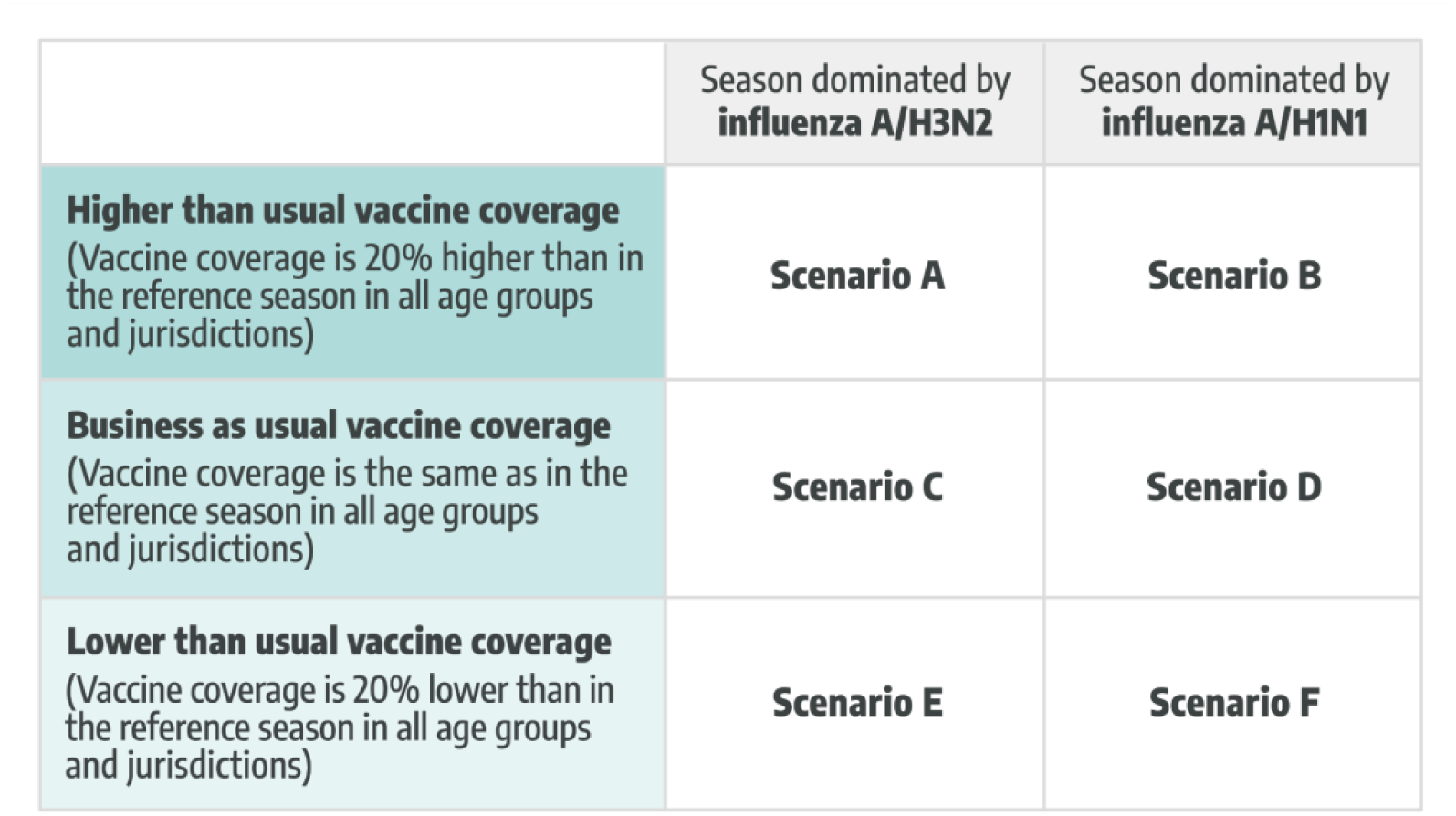
Scenarios for Round 1 of seasons 2023-2024 and 2024-2025, U.S. Flu Scenario Modeling Hub. Summary of assumptions for different scenarios [3]. Six scenarios are defined describing the interaction between vaccination coverage and the dominant influenza A virus subtype. Due to data availability, assumptions related to vaccination coverage are based on the 2021-2022 season for the 2023-2024 projections, and on the 2022-2023 season for the 2024-2025 projections.

Modeling teams were required to assume vaccine effectiveness (VE) of 40% against medically attended influenza illnesses and hospitalizations, in line with evidence from the 2017-2018 and 2019-2020 seasons [52, 54-55]. Assumptions about VE against infection and transmission, prior immunity, seeding intensity, timing, geographic distribution, and the mix of circulating strains at the start of the projection period were left to the teams’ discretion. More information on the specifics of the scenario rounds is reported in the Supplementary Information and the U.S. Flu Scenario Modeling Hub GitHub Repository [2].

For the 2023-2024, eight teams contributed projections from nine models, whereas in the 2024-2025 round, seven modeling teams contributed projections from eight models. Only models that submitted projections for at least the U.S. national level are considered here; this exclusion criterion led to the exclusion of one model for the 2024-2025 season. Most models submitted projections for both the national and subnational levels, with subnational referring to the state level. In the 2023-2024 season, of the nine models, seven were mechanistic (including two metapopulation and one agent-based models), one was semi-mechanistic, and one was statistical. Overall, seven out of nine models incorporated age stratification, and six out of nine provided projections at both national and subnational resolution. In the 2024-2025 season, of the eight models, six were mechanistic (including two metapopulation and two agent-based models), one was semi-mechanistic, and one was statistical. Two models did not incorporate age stratification, but all models provided projections at both national and subnational resolution. Each model provided from 100 to 300 representative individual trajectories for each scenario and geographic unit, with each trajectory representing a stochastic realization of weekly influenza hospital admissions, and a minimum of 100 trajectories being required (see Table S1 for details on the number of trajectories submitted by each model). Teams provided projections for the overall population and, optionally, for specific age groups. In this study, we focus on projections for the overall population. Additionally, we excluded from the analysis the weeks where the overall U.S. influenza-like-illness incidence rate was below the baseline threshold determined and reported by the Centers for Disease Control and Prevention through the FluView Weekly U.S. Influenza Surveillance Report [6]. Using this criterion, we consider 20 weeks for the 2023-2024 season from November 11, 2023 to March 22, 2024, and 17 weeks for the 2024-2025 season from November 30, 2024 to March 23, 2025. After submission, individual model trajectories related to each scenario and geographic unit were combined into ensemble projections using a Linear Opinion Pool (LOP) approach, which averages the cumulative probabilities for each incident hospitalization value projected by individual models [7]. This equates to pooling individual trajectories across all models, creating a single aggregate probability distribution of hospitalizations for each week. The LOP approach includes two different methods of aggregating projections: a trimmed version, which excludes the two outermost values to reduce the variance, and an untrimmed version, where all values are included in the averaging [5]. Here, we consider the untrimmed version as it preserves all values.

In this work, we refer to the *original ensemble^2^* as the ensemble computed using the LOP procedure, considering all the trajectories from all models and scenarios combined, without any selection, thus including the full set of original trajectories. The term *ensemble^2^* denotes that the aggregation is performed across both models and scenarios, rather than only across models, as in standard ensemble approaches [5]. This method aligns with previous research demonstrating that an *ensemble^2^* approach (i.e., integrating all scenarios) can improve predictive performance [4]. This *original ensemble*^2^ serves as a benchmark for evaluating the performance improvements achieved by the adaptive ensemble approach. In this context, it represents a static version of the adaptive ensemble, as it does not incorporate updates based on observed data. It is worth noting that this original ensemble differs from the untrimmed ensembles published by the U.S. Flu Scenario Modeling Hub, which are computed separately for each scenario, resulting in six distinct ensembles. The reason for our choice is that by combining all scenarios, we aim to streamline comparisons with the adaptive ensemble. However, for completeness, we also compare the adaptive ensemble to the ensembles computed separately for each scenario in the Supplementary Information, finding similar performance trends.

#### 4.1.2 FluSight short-term forecasts

For the evaluation of the proposed approach in the short-term forecasting task, we consider data and forecasts from the 2023-2024 and 2024-2025 seasons of the CDC FluSight challenge. Forecasts were submitted weekly and made available through the FluSight GitHub repository [1]. Each week during the influenza season, typically running from October through May of the next year, participating teams are invited to submit national and jurisdiction-specific probabilistic nowcasts and forecasts on weekly confirmed influenza hospital admissions for the prior week, the current week, and the following three weeks. Here we consider only forecasts at the national U.S. level. The FluSight forecasting hub produces two real-time outputs each week: a baseline model and an ensemble model. The *FluSight baseline*, used as a neutral benchmark for model evaluation, consistently forecasts the last observed value as the median, with prediction intervals based on historical data. The *FluSight ensemble* aggregates forecasts from independent individual models [8], many of which are specifically designed and iteratively optimized for short-term forecasting. The combined forecast is computed as the median across corresponding quantiles from contributing models. During the 2023-2024 season, 38 models submitted over 30 forecasting rounds, from October 14, 2023, to May 5, 2024, and in 2024-2025, 50 models contributed to FluSight throughout 27 forecasting rounds, from November 23, 2024 to May 28, 2025. Each forecasting round corresponds to a week [30, 59-60]. Not all models contributed forecasts for every round. More information on individual models contributing to FluSight is available in Table S2.

### 4.2 Adaptive ensemble

In this section, we describe the methodology for constructing the adaptive ensemble. Being conditional predictions of future outcomes, scenario projections depend on specific assumptions about uncertain drivers of future dynamics. Because the impact of such drivers becomes increasingly uncertain over longer time horizons, scenario projections are useful for exploring best- and worst-case possibilities. In other words, they typically bracket the range of what is likely to occur. For this reason, the set of scenario projections provides a pool of possible future trajectories. As new evidence becomes available (e.g., data points), we generate the adaptive ensemble by selecting those trajectories that most closely align with the observations. The intuition is that this allows us to retain the diversity of scenario-based thinking while adaptively retaining those trajectories that are more consistent with reality.

Specifically, we propose two similar but distinct methods: one designed for adaptive scenario projections and the other for short-term forecasting. In both cases, we adopt the following notation. For each scenario *s*, the set of trajectories contributed by the model *m* is indicated as *T*^*s*,*m*^ = {*ĥ*_*i*_^*s*,*m*^ ∶ *i* ∈ {1, …, *R*}}, where *ĥ*_i_^*s*,*m*^ is the *i-th* model trajectory (e.g., one stochastic realization of the number of weekly hospital admissions over time), and R denotes the total number of trajectories per model and scenario. We also indicate as *T*^*m*^ = ⋃_*s*_ *T*^*s*,*m*^ the set of all trajectories for the model *m* (i.e., aggregating trajectories from all scenarios for a fixed model), as *T*^*s*^= ⋃_*m*_ *T*^*s*,*m*^the set of all trajectories for scenario *s* (i.e., aggregating trajectories from all models for a fixed scenario), and as *T* = ⋃_*s*,*m*_ *T*^*s*,*m*^ the set of all models’ trajectories contributing to all scenarios (i.e., aggregating trajectories from all models and scenarios). We indicate reported data points between time *t*_0_ and *t* as *h*_*t*0:*t*_. We then introduce a general loss function *δ*(⋅), and the set of the loss function values computed between the reported data and all trajectories for a scenario *s* and model *m* in the time window *t*_0_ to *t* as 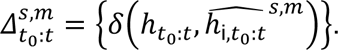 We define 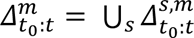 as the set of the loss function values of all trajectories for a model *m* across all scenarios, 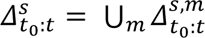 as the set of the loss function values of all trajectories for a scenario *s* across all models, and 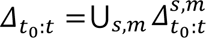 as the set of the loss function values of all trajectories.

In both scenario modeling and short-term forecasting, the trajectory selection algorithm is applied iteratively at each time step as new data become available. Consequently, a new adaptive ensemble is generated at each time step using all available reported data up to that point for trajectory selection. It is important to note that the selection procedure at different time steps is completely independent; therefore, the trajectories included in the subset of the adaptive ensemble at one time step may or may not be included in the subsequent steps. To accurately reflect real-world data constraints in real-time infectious disease modeling, such as incomplete data at the time of reporting, we use unrevised reported data for trajectory selection (i.e., data available at the time of selection, which may be subject to varying degrees of backfilling) while evaluating the performance of the adaptive ensemble against the revised and finalized data.

#### 4.2.1 Adaptive ensemble for long-term scenario projections

In the case of scenario modeling projections, we aim to generate the adaptive ensemble projection for the remainder of the projection window as follows. At time *t*, a new data point *h*_*t*_ becomes available. Then we select the set *T*_*t*_ of trajectories that satisfy the following condition:

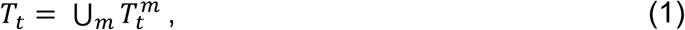

with

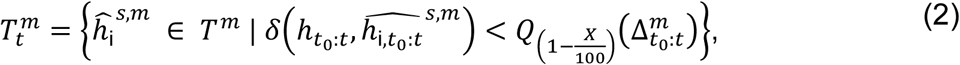

where *T*_*t*_^*m*^ is the set of trajectories at time *t* selected for each model *m*, and *Q*_*α*_ (·) is a function that returns the top *α*-th percentile of the quantity to which it is applied. In simple terms, we select the top *X*% of trajectories based on the loss function *δ*(⋅) across all scenarios for each model independently. Then we compute the adaptive ensemble by applying a linear opinion pool (LOP) approach on the selected trajectories. That is, for each model *m* we estimate the cumulative density function (CDF) *F*_*t*_^*m*^ of different hospitalization values considering the selected trajectories for that model (*T*_*t*_^*m*^). The CDF represents the probability that the projected outcome (i.e., influenza hospitalizations) will be less than or equal to a given value, providing a probabilistic summary of selected trajectories for each model at a specific time point. Then, the CDF of the adaptive ensemble (AE) will simply be a weighted average of individual models CDFs:

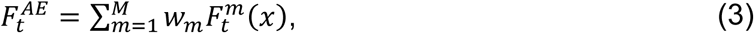

where *M* is the number of models, and *w_m_* is the weight of model *m*. In the following, we adopt the same weighting approach used in the original U.S. Flu Scenario Modeling Hub projections. Specifically, the unnormalized weight is set to one for each team, ensuring equal initial weighting across teams. If a team submits two models, each model is assigned half the weight to maintain parity at the team level. Finally, the quantiles of the adaptive ensemble are computed as the inverse of the computed CDF. For example, the median of the adaptive ensemble computed at time *t* will be (*F_t_*^*AE*^)^−1^(0.5).

The adaptive ensemble approach is analogous to using an Approximate Bayesian Computation (ABC) rejection algorithm for calibrating each model [1]. The loss function threshold serves as the tolerance for the ABC rejection criterion, effectively selecting the posterior distribution of trajectories for each model. This analogy assumes that each modeling team has implicitly defined a prior distribution within their original projection set, informed by their assumptions and scenario guidelines. With this approach, we ensure an equal proportion of trajectories is selected from each model. This is implemented in order to maintain model variability and diversity within the adaptive ensemble, a feature driving ensemble performance, particularly for long-term scenario projections. Additionally, the adaptive ensemble is computed by selecting trajectories from the combined pool of all scenarios [4], excluding counterfactual scenarios (i.e., scenarios that assume unrealistic conditions and are considered for comparison). This approach enables the derivation of a proxy for scenario posterior probability, as explained further below in Section 4.2.2.

In the following, we use the weighted mean absolute percentage error (wMAPE) as the loss function for trajectory selection in the case of long-term scenario modeling projections. The wMAPE is defined as:

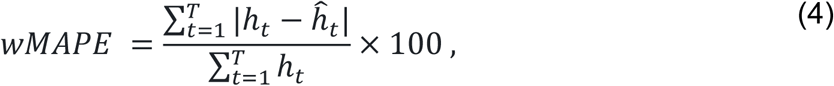

where T is the evaluation period length, *ĥ*_*t*_ is the projected and *h*_*t*_ is the actual value.

In the Supplementary Information, we repeat the analysis using root mean squared error (RMSE) instead of wMAPE as the loss function for trajectory selection.

#### 4.2.2 Estimating epidemic scenarios posterior probability

The adaptive ensemble is constructed by selecting trajectories from a combined pool of projections spanning all scenarios. Arguably, this approach has the potential to improve adaptive ensemble performance by increasing the pool of trajectories for selection [1].

Additionally, reality rarely aligns perfectly with any single scenario but instead exists in an intermediate space between scenarios. From this perspective, it seems more appropriate for the ensemble to represent a weighted mixture of different scenarios [8]. On the other hand, this approach may lead to a loss of interpretability due to the mixing of multiple scenarios. To address this issue, we determine the plausibility of each scenario as part of the trajectory selection procedure; more precisely, we keep track of the fraction of trajectories selected from each scenario in the case of adaptive ensemble at each time step *t*. It is important to note that the interpretation of posterior scenario probabilities depends on the underlying scenario design choices [3]. For example, counterfactual scenarios should generally be excluded from ensembling or assigned lower weights, as they are not intended to reflect likely real-world outcomes. Similarly, scenarios designed to explore specific policy implementations or interventions may not correspond to actual conditions, limiting the effectiveness of the adaptive strategy. However, in this specific work, such scenarios were not included in either 2023-2024 or 2024-2025 rounds of the U.S. Flu Scenario Modeling Hub and therefore do not present a concern in this analysis.

Formally, we compute the fraction of trajectories originating from the scenario *s* contributing to the adaptive ensemble at time *t* as 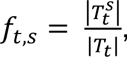 where *T*^*s*^ is the set of trajectories originating from the scenario *s* included in *T*_*t*_ and |⋅| returns the size of a set (i.e., number of trajectories). Finally, by considering the analogy of this approach with an ABC rejection algorithm, *f*_*t*,*s*_ yields an estimate of the posterior probability of each scenario *s* based on the information available at time *t*.

#### 4.2.3 Adaptive ensemble for short-term forecasting

In the case of short-term forecasting, instead of generating the adaptive ensemble for the remainder of the projection window, we focus on forecasting the next four weeks and proceed as follows. At time *t*, a new data point *h*_*t*_ becomes available. Then we select the set of trajectories *T*_*t*_ that satisfies the following condition:

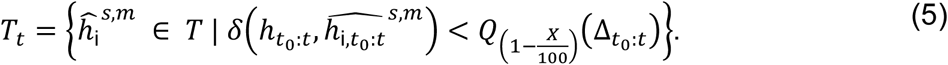

In simple terms, we select the top *X*% of trajectories based on the loss function *δ*(⋅) across all scenarios and models. Finally, the quantiles of the adaptive ensemble are computed directly from the set of selected trajectories. For instance, the median of the adaptive ensemble is represented as *Q*_0.5_(*T*_*t*_). We then consider the projections between *t* and *t* + *h* (where *h* is the forecasting horizon) as the short-term forecast of the adaptive ensemble.

There are two key differences between this approach and the adaptive ensemble methodology used for scenario modeling. In the scenario modeling case, trajectories were selected independently for each model, ensuring equal representation of models in the ensemble. Here, models are represented unequally in the final set of selected trajectories.

This adjustment is intentional, as the short-horizon nature of the forecasting exercise prioritizes selecting trajectories that best align with recent observed trends, rather than preserving model variability. Second, the quantiles of the adaptive ensemble are computed directly from the set of selected trajectories, rather than by first constructing the CDF using the LOP procedure as done for scenario modeling. We opt for such an approach because computing quantiles directly from the pool of trajectories is equivalent to an unweighted LOP.

The forecasts produced by the adaptive ensemble are then compared against two benchmark models: i) the FluSight baseline model, which serves as a neutral benchmark by consistently forecasting the last observed value as the median and estimating prediction intervals based on historical data, and ii) the FluSight ensemble, which aggregates forecasts from contributing models and serves as the FluSight hub’s official output for external reporting and public dissemination.

In the case of trajectory selection for short-term forecasting, we use a generalized version of the wMAPE, which gives more importance to recent data points. It is defined as:

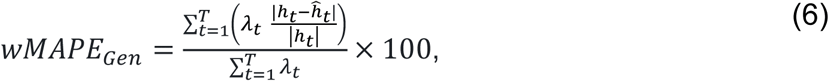

where T is the evaluation period length, 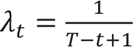 is a linear decay weight, which gives more importance to recent data, *ĥ*_*t*_ is the projected value and *h*_*t*_ is the actual value.

### 4.3 Scoring rules for performance evaluation

After generating the adaptive ensemble, we evaluate its performance on the out-of-sample window (i.e., data points that were not used for trajectory selection) using different scoring rules, namely the Weighted Interval Score (WIS), the Mean Absolute Error (MAE), and prediction interval coverage.

We note how, in the case of scenario modeling, the evaluation is done on all future data points, but in the case of short-term forecasting, only on the 1- to 4-weeks-ahead forecasting horizons (i.e., the FluSight targets of the current week, and up to three weeks ahead).

The WIS approximates the continuous ranked probability score (CRPS) [2]. For a given (1 − *α*) prediction interval of a forecast *F*, the interval score (IS) is defined as:

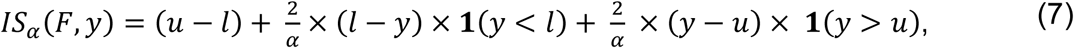

where **1** is the indicator function, *l* (*u*) denotes the lower (upper) limit of the prediction interval, and *y* is the actual observed value. The WIS expands the interval score to several prediction intervals at different levels and it is defined as:

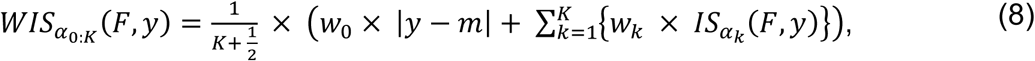

where *K* is the number of prediction intervals considered, and *m* is the prediction median. Here we consider 11 prediction intervals (*α*_*k*_ = 0.02, 0.05, 0.10, 0.20, 0.30, 0.40, 0.50, 0.60, 0.70, 0.80, 0.90) and, following a common approach in the literature we set 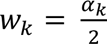 and 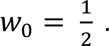 Note that the WIS is defined for each time step; therefore, in the following we consider WIS values averaged over the evaluation period if not specified otherwise.

The MAE of the median is defined as:

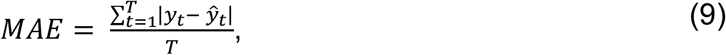

where *y*_*t*_ are the actual values and *y*^_*t*_ are the predicted medians in the evaluation period of length T.

The prediction interval coverage is a measure used for assessing the calibration of a model, which describes how closely forecasted probabilities align with observed frequencies. It defines the fraction of times that actual data fall within the corresponding prediction intervals. Specifically, in a perfectly calibrated model a 50% predictive interval will contain exactly 50% of the observations, meaning the actual coverage matches the nominal coverage. Mathematically, for N data points *y*_*i*_, and for a given (1 − *α*) prediction interval, the coverage is computed as follows:

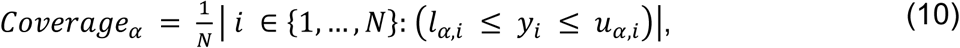

where *l*_*α*,*i*_ and *u*_*α*,*i*_ are the lower and upper bounds of the prediction interval of the i-th forecast.

## Declarations

### Code Availability

The code to reproduce all the analyses and figures is available in https://github.com/sfiandrino/adaptive_ensemble

### Data Availability

The data to reproduce all the analyses are available in https://github.com/sfiandrino/adaptive_ensemble, https://github.com/cdcepi/FluSight-forecast-hub/tree/main/auxiliary-data/target-data-archive, https://github.com/midas-network/flu-scenario-modeling-hub

## Acknowledgements

We gratefully acknowledge the Flu Scenario Modeling Hub and the FluSight Influenza Forecast Hub.

## Disclaimer

The findings and conclusions in this report are those of the authors and do not necessarily represent the official position of the Centers for Disease Control and Prevention. The content of this publication does not necessarily reflect the views or policies of the National Institutes of Health or the Department of Health and Human Services, nor does mention of trade names, commercial products or organizations imply endorsement by the U.S. Government.

