## Supplementary Information for "Adaptive multi-model ensembles for improved epidemic projections and decision support"

### S1. Summary of individual models that submitted projections in Round 1 2024-2025 and 2023-2024 of the U.S. Flu Scenario Modeling Hub (SMH)

#### CDDEP FluCompModel

**Team:** CDDEP (Center for Disease Dynamics, Economics & Policy)
**Model:** FluCompModel
**Contributors:** Fardad Haghpanah, Eili Klein

**General Description:**
FluCompModel is an age-stratified compartmental model of influenza transmission with a Markov Chain Monte Carlo (MCMC) wrapper for parameter estimation. The model evaluates the potential impact of increased vaccine effectiveness and prior immunity on hospitalizations and mortality in the United States.

**Methods:**

- Calibration: The model relies on weekly reported cases, hospitalizations, and mortality data from the CDC for model calibration. It uses MCMC for parameter estimation. The projections are calculated based on simulations using the obtained posterior parameter samples.
- Non-fixed parameters: Transmission coefficient [0, 1], vaccine response (delay) rate [0, 0.1], asymptomatic infectiousness rate [0, 0.5], symptomatic infectiousness rate [0, 0.5], hospitalization ratio [0, 0.01], hospital discharge rate [0, 0.2], death rate [0, 05], prior immunity [0.25, 0.35], reporting ratio [0, 0.5].
- Age groups: 0-17, 18-64, 65+.
- Contact matrix/mixing structure: the model considers contact matrices from Prem et al., 2017 [1].

**Assumptions:**

- Seasonality: Seasonal forcing was estimated based on human behavior using a cosine function fitted to indoor activity pattern data [2]. The estimated seasonal forcing dynamically affects the transmission coefficient through time.
- Vaccine: Two levels of vaccine response are considered: i) protection against infection and transmission; ii) protection against hospitalization only. Age specific values were calculated based on the Centers for Disease Control and Prevention (CDC) post season vaccine effectiveness (VE) estimates and

other studies [3]. VE-infection for H1N1: 40%, 40%, 39%; for H3N2: 55%, 38%, 20%. VE-hospitalization for both subtypes: 65%, 55%, 50%, respectively for 0-17, 18-64, and 65+ age groups. Vaccine efficacy (VE) delay is accounted through a delay parameter.
- Immunity: Duration of natural immunity is assumed to be longer than a flu season, and duration of vaccine immunity is assumed to follow an exponential distribution with a mean of 300 days. The initial estimate of immunity is sampled from a uniform distribution U(0.25, 0.35).
- Subtypes: The model is single-strain, and no interactions between strains/subtypes are implicitly modeled.
- Non-pharmaceutical interventions (NPIs): NPIs not implemented in the model.

**Contributions:**
- Seasons: 2023-2024
- Geographies: 1 (iso2code: US)

**Reference paper:**
Haghpanah, F., Hamilton, A. & Klein, E. Modeling the potential health impacts of delayed strain selection on influenza hospitalization and mortality with mRNA vaccines. *Vaccine X* **14**, 100287 (2023) [4].

#### MOBS_NEU GLEAM_FLU

**Team:** MOBS_NEU (MOBS Lab at Northeastern University)
**Model:** GLEAM_FLU
**Contributors:** Matteo Chinazzi, Jessica T. Davis, Clara Bay, Alessandro Vespignani

**General Description:**
GLEAM_FLU is an age-structured stochastic, metapopulation model that includes metropolitan area demographic data, short-range commuting flows, domestic air traffic, and age-specific contact patterns. The model works at the resolution of roughly 400 aggregated subpopulations to provide state-level results. A natural history of the disease for infectious and asymptomatic individuals and specific compartmentalization for vaccinated individuals are used.

**Methods:**
- Calibration: The calibration is performed using an Approximate Bayesian Computation (ABC) rejection approach comparing the weekly model-

estimated hospitalizations with those reported by the U.S. Department of Health and Human Services, respectively. The distance between surveillance data and model estimates is measured using the weighted mean absolute percentage error and/or residuals. The selected stochastic realizations generate a posterior distribution.

- Non-fixed parameters: Effective reproduction number, seasonality multiplier, starting date/initial conditions, infection hospitalization rate (ranges depend on scenario), residual population immunity.
- Age-groups: 0-4, 5-17, 18-49, 50-64, 65+.
- Contact matrix/mixing structure: Contact rates are explicitly modeled using contact matrices from Mistry et al. (2021) [5]. Contact rates change due to school calendar (school closures/calendars follow observed data).

**Assumptions:**

- Seasonality: The changes in transmissibility due to seasonality are explicitly modeled following Balcan et al. 2010 [6].
- Vaccine: Vaccines are modeled in a leaky way with efficacies as specified by the Scenario Modeling Hub (SMH) scenarios.
- Immunity: Duration of natural immunity holds for current flu season and initial estimates of immunity depend on SMH scenarios.
- Subtypes: The model is single strain and no interactions between strains/subtypes are implicitly modeled.
- Non-pharmaceutical interventions: NPIs not implemented in the model.

**Contributions:**

- Seasons: 2023-2024, 2024-2025
- Geographies: 52 (iso2code: 01, 02, 04, 05, 06, 08, 09, 10, 11, 12, 13, 15, 16, 17, 18, 19, 20, 21, 22, 23, 24, 25, 26, 27, 28, 29, 30, 31, 32, 33, 34, 35, 36, 37, 38, 39, 40, 41, 42, 44, 45, 46, 47, 48, 49, 50, 51, 53, 54, 55, 56, US)

**Reference paper:**

Davis, J. T. *et al.* Cryptic transmission of SARS-CoV-2 and the first COVID-19 wave. *Nature* **600**, 127–132 (2021) [7].

#### NIH FLUD

**Team:** NIH (Fogarty International Center, National Institutes of Health)
**Model:** FluD
**Contributors:** Samantha Bents, Cécile Viboud

**General Description:**

FLUD is an ordinary differential equation (ODE) based age-structured deterministic model following the SEIRS (Susceptible-Exposed-Infectious-Recovered-Susceptible) compartmental structure.

**Methods:**

- Calibration: The model fits age-specific hospitalization rates, transmission coefficient, and seasonality parameters to historical time series using maximum likelihood estimation. This gives 95% CI for each parameter from which sampled when projecting forward. The model is calibrated to FluSurv-NET and the U.S. Department of Health and Human Services (HHS) hospitalization data.
- Non-fixed parameters: transmission coefficient [0.01, 0.05], seasonality parameters (amplitude of seasonal forcing [0.01, 0.03], phase shift [0, $2\pi$], age-specific hospitalization rates [0,1], NPI reduction in transmission [0, 50%]
- Age groups: 0-4, 5-19, 20-49, 50-64, 65+.
- Contact matrix/mixing structure: The model includes POLYMOD contact matrix [1].

**Assumptions:**

- Seasonality: The model considers seasonality parameters (phase shift, amplitude of seasonal forcing) within a cosine function in the transmission expression.
- Vaccine: This model does not include a vaccine compartment, but it incorporates the impact of vaccination in a post-processing step. A regression over multiple years of historical flu burden against annual VE and coverage is applied to recover effect sizes of covariates with 95% CI. The effect sizes are then applied post-processing to the hospitalization burden estimates generated by the model.
- Immunity: The duration of natural immunity holds for 2 years.
- Subtypes: The model is calibrated to separate AH1 and AH3 time series. These time series are calculated by multiplying historic CDC Flusurv-NET time series by CDC FluView weekly subtype rates.
- Non-pharmaceutical interventions: NPIs not implemented in the model.

**Contributions:**

- Seasons: 2023-2024
- Geographies: 1 (iso2code: US)

**Reference paper:**

Hogan, A. B. *et al.* Potential impact of a maternal vaccine for RSV: a mathematical modelling study. *Vaccine* **35**, 6172–6179 (2017) [8].

#### NIH Flu_TS

**Team:** NIH (Fogarty International Center, National Institutes of Health)
**Model:** Flu_TS
**Contributors:** Amanda Perofsky, Cécile Viboud

##### General Description:

Flu_TS uses time series regression models with AutoRegressive Integrated Moving Average (ARIMA) or AutoRegressive Moving Average (ARMA) errors and exogenous covariates for seasonal cumulative vaccination coverage, seasonal vaccine effectiveness, weekly A/H3N2 virus circulation, and weekly estimated influenza infections. For individual states, seasonality is modeled with Fourier terms (dynamic harmonic regression); for the U.S., seasonality is captured by the structure of an automatically selected ARIMA/ARMA model. This approach requires providing values for the exogenous covariates during the projection period.

##### Methods:

- Calibration: The model uses historical data on influenza hospitalizations, influenza-like illness rates, influenza A subtype circulation, and vaccine coverage and effectiveness for calibration. Influenza-like illness rates are used to derive the weekly estimated influenza infections covariate rather than entering the model as a separate regressor. The out-of-season projections for historical seasons prior to the COVID-19 pandemic are used to determine the optimal model training period.
- Non-fixed parameters: The non-seasonal orders – the number of autoregressive terms (p), the degree of differencing (d), and the number of moving-average terms (q) – and, for the United States, their seasonal counterparts (P, D, Q), are not fixed in advance. They are selected automatically using a variation of the Hyndman-Khandakar (HK) algorithm for automatic ARIMA modeling (the ARIMA function in the fable R package): the differencing order is determined by repeated Kwiatkowski–Phillips–Schmidt–Shin (KPSS) tests, and the AR and MA orders are then chosen to minimize the corrected Akaike Information Criterion (AICc) on the differenced series, using a stepwise search through the model space rather than an exhaustive evaluation of every order combination. The United States and state models differ only in how seasonality is handled, and therefore in which orders the search considers: for the United States, the search ranges over both non-seasonal and seasonal orders, whereas for individual states seasonality is instead modeled with Fourier terms (dynamic harmonic regression) and only

the non-seasonal orders of the ARIMA/ARMA errors are selected. The number of Fourier terms is itself tuned (see Seasonality section).

- Age groups: no age-stratification.
- Contact matrix/mixing structure: no mixing structure.

**Assumptions:**

- Seasonality: For the U.S., the Hyndman–Khandakar (HK) algorithm may return a seasonal ARIMA, a non-seasonal ARIMA, or an ARMA model depending on the data, so an explicit seasonal term is not always present. For states, the optimal number of Fourier terms was determined by 1) training models on historical weekly hospitalizations up to the 2019-2020 season and comparing AICc values across models with different orders of Fourier terms, and 2) conducting out-of-season projections for recent seasons prior to COVID-19 (2016-17, 2017-18, 2018-19, and 2019-20) and ranking the median out-of-season accuracy (Root Mean Square Error (RMSE) and Continuous Ranked Probability Score (CRPS)) of models with different orders of Fourier terms. The final models for states include 12 Fourier terms.
- Vaccine: Only VE against symptoms is considered. For historical seasons, adjusted vaccine effectiveness estimates for influenza-like-illness (ILI) or acute-respiratory-infection (ARI) are obtained from the CDC's influenza VE website or published observational studies, the majority of which were test-negative case control design and focused on general populations or healthy adult populations in North America or Europe. If more than one VE study was available for a particular historical season, the seasonal weighted mean VE is considered, with individual VE values weighted by the inverse of their 95% confidence intervals. One fixed VE value per season was used as an exogenous covariate in model training and projections. Projections used vaccine effectiveness (40%), as dictated by the SMH. For model training, the model relies on national and state-specific monthly vaccination coverage estimates for individuals aged 6+ months during seasons 2010-2011 to 2021-2022, retrieved from publicly available data from the National Center for Immunization and Respiratory Diseases; for each historical season, the final cumulative coverage value is used for the United States and each state. For the projection period, final cumulative vaccination coverage for each location is extracted from the coverage curves provided by the SMH.
- Immunity: Immunity is not explicitly represented in the time series model. Population susceptibility enters indirectly through the weekly estimated influenza infections covariate, which is produced by a semi-mechanistic renewal model (epidemia R package) that assumes an initial susceptible fraction of 0.65 [9], treats infection as removing individuals from the susceptible pool (no within-season waning of natural immunity), and does not

implement a vaccine effect on susceptibility (the vaccine signal enters the time series model separately through the VE covariate).

- Subtypes: Strain- or subtype-specific hospitalizations have not been trained or projected. However, they account for the proportion of samples testing positive for A/H3N2 (from CDC FluView) as an exogenous covariate in model training and projections. The focus on percent positive for A/H3N2 rather than percent positive for influenza A is because A/H3N2 dominant seasons tend to be more severe than A/H1N1 dominant seasons.
- Non-pharmaceutical interventions: NPIs not implemented in the model.

**Contributions:**

- Seasons: 2023-2024, 2024-2025
- Geographies:
  - 2023-2024: 13 (iso2code: 47, 13, 24, 41, 08, 09, 35, 39, 36, 06, 26, 27, US)
  - 2024-2025: 39 (iso2code: 01, 04, 05, 06, 08, 09, 13, 15, 16, 17, 18, 20, 21, 22, 23, 24, 25, 26, 27, 28, 29, 30, 31, 35, 36, 37, 39, 40, 42, 45, 46, 47, 48, 50, 51, 53, 54, 55, US)

#### University of Notre Dame FRED

**Team:** University of Notre Dame
**Model:** FRED
**Contributors:** Guido Espana, Sean Moore, Alex Perkins

**General Description:**
FRED is an agent-based model that uses a synthetic population with demographic and geographic characteristics of each state's real population, including age, household composition, household location, and occupation.

**Methods:**

- Calibration: The model uses the sobol sequence to generate 1000 parameter sets for each state. Fit the model to 2017-18 and 2019-20 seasons separately. Likelihood calculated for cumulative age-specific attack rates from each year and age-specific cumulative hospitalization rates from Fluserv.
- Non-fixed parameters: Transmissibility (0.1, 2.0), nursing home importation rate (0.01, 0.1), same-age bias in neighborhood contact rate (0.01, 0.1), neighborhood contact rate (0.08, 1.18), age-specific hospitalization rates (0.0005, 0.06).
- Age groups: 0-4, 5-17, 18-49, 50-64, 65+.

- Contact matrix/mixing structure: The model includes contact matrices.

**Assumptions:**

- Seasonality: Human contact rate is adjusted using seasonality estimates.
- Vaccine: VE affecting transmission, infection and hospitalization is strain specific: H1N1: 0.4 (all ages) H3N2: 0.68 (0-9), 0.32 (10-17), 0.33 (18-49), 0.3 (50-64), 0.17 (65+). The VE delay is set to 14 days.
- Immunity: The duration of natural immunity holds for seasonal projections. The duration of vaccine immunity follows exponential distribution with mean of 180 days. The initial estimate of immunity is drawn from a uniform distribution of (0.335, 0.35).
- Subtypes: The model considers one subtype (influenza A).
- Non-pharmaceutical interventions: NPIs not implemented in the model.

**Contributions:**

- Seasons: 2023-2024, 2024-2025
- Geographies: 52 (iso2code: 01, 02, 04, 05, 06, 08, 09, 10, 11, 12, 13, 15, 16, 17, 18, 19, 20, 21, 22, 23, 24, 25, 26, 27, 28, 29, 30, 31, 32, 33, 34, 35, 36, 37, 38, 39, 40, 41, 42, 44, 45, 46, 47, 48, 49, 50, 51, 53, 54, 55, 56, US)

**Reference paper:**

Espana, G. *et al.* Impacts of K-12 school reopening on the COVID-19 epidemic in Indiana, USA. *Epidemics* **37**, 100487 (2021) [10].

#### PSI M2

**Team:** Predictive Science Inc (PSI)
**Model:** M2
**Contributors:** Ben-Nun M., Turtle J., Riley P.

**General Description:**

M2 is a mechanistic model with S[Sv]I[Iv]HR compartments, where 'v' subscripts indicate vaccinated, and each compartment is age-stratified.

**Methods:**

- Calibration: The model uses MCMC fit to FluSurvNet total network values for exemplar seasons.
- Non-fixed parameters: Reproduction number [1.1, 1.4], infection hospitalization ratio [0.008, 0.02], initial number of infections [1, N/1E5].

- Age groups: four age groups are modeled/fit (0-17, 18-49, 50-64, 65+), but only one (0-130yrs) is submitted.
- Contact matrix/mixing structure: The model includes contact matrices based on the Mobs-Lab mixing-patterns github.

**Assumptions:**

- Seasonality: An arbitrary seasonal forcing function was fit. Additionally, projection timings were filtered such that they roughly match historic ILI+ peak distributions.
- Vaccine: VE affecting transmission is assumed to be 1/2 of VE against hospitalization—this was tuned in the 2024-25 submission such that model VE better matched observed VE. Vaccine efficacy delay is not considered.
- Immunity: The duration of natural immunity and of vaccine immunity is not modeled. Initial estimates of population susceptible are assumed 70%.
- Subtypes: Subtypes are not modeled.
- Non-pharmaceutical interventions: NPIs are not explicitly modeled.

**Contributions:**

- Seasons: 2023-2024, 2024-2025
- Geographies: 52 (iso2code: 01, 02, 04, 05, 06, 08, 09, 10, 11, 12, 13, 15, 16, 17, 18, 19, 20, 21, 22, 23, 24, 25, 26, 27, 28, 29, 30, 31, 32, 33, 34, 35, 36, 37, 38, 39, 40, 41, 42, 44, 45, 46, 47, 48, 49, 50, 51, 53, 54, 55, 56, US)

#### University of Southern California SIkJalpha

**Team:** University of Southern California (USC)
**Model:** SIkJalpha
**Contributors:** Ajitesh Srivastava, Majd Al Aawar

**General Description:**
Discrete heterogeneous rate model where rates are learned using regression weighing the recently seen data higher. Past seasons' rates are used as a proxy for seasonality.

**Methods:**

- Calibration: The calibration is based on a previous select season. Data inputs were taken from HHS & Flusurvnet.
- Non-fixed parameters: Modeling is done at the infection level assuming a range of hospitalization rates: (0.5% to 3.5%).
- Age groups: 0-4, 5-17, 18-49, 50-64, 65-130.

- Contact matrix/mixing structure: no mixing structure is considered.

**Assumptions:**

- Seasonality: Seasonality is estimated by looking at the past seasons' rates
- Vaccine: VE affecting transmission and infection is not modeled. VE against hospitalization and coverage contribute to higher population immunity which in turn reduces hospitalizations. VE delay is set to 14 days.
- Immunity: The duration of natural immunity and of vaccine immunity is not modeled. Initial estimate of immunity was set to 33% for prior seasons, some optimization is then done to estimate initial immunity for this season.
- Subtypes: Subtypes are not modeled.
- Non-pharmaceutical interventions: NPIs not implemented in the model.

**Contributions:**

- Seasons: 2023-2024, 2024-2025
- Geographies: 57 (iso2code: 01, 02, 04, 05, 06, 08, 09, 10, 11, 12, 13, 15, 16, 17, 18, 19, 20, 21, 22, 23, 24, 25, 26, 27, 28, 29, 30, 31, 32, 33, 34, 35, 36, 37, 38, 39, 40, 41, 42, 44, 45, 46, 47, 48, 49, 50, 51, 53, 54, 55, 56, 60, 66, 69, 72, 78, US)

#### UT ImmunoSEIRS

**Team:** University of Texas (UT)
**Model:** ImmunoSEIRS
**Contributors:** Shraddha R Bandekar, Kaiming Bi, Anass Bouchnita, Spencer J. Fox, Lauren Ancel Meyers, UT COVID-19 Modeling Consortium.

**General Description:**

ImmunoSEIRS is a new two-viruses (H1N1 and H3N2) model that explicitly tracks the immunity caused by natural infections and vaccination and its impact on the average chances of infection, hospitalization, and death.

**Methods:**

- Calibration: The calibration is based on least squares and the outcomes are deaths and hospitalizations.
- Non-fixed parameters: Hospitalization rates, and death rates.

- Age groups: 0-4, 5-11, 12-18, 19-49, 50-64, 65+.
- Contact matrix/mixing structure: The model is contact matrix based between different age groups, for weekday, school closure day, and weekend or holiday, we used different contact matrix.

**Assumptions:**

- Seasonality: The seasonality is based on contact behavior difference in weekday, school closure day, and weekend or holiday, and humidity impact
- Vaccine: VE affecting transmission is not modeled. The model assumes VE against both hospitalizations and infection. VE against hospitalization is modeled with ranges different for each year and round. VE against infection is set to 21% across all age groups, based on the reference values provided by the hub. Vaccine efficacy delay is not considered.
- Immunity: The duration of natural immunity is set to 18 months half time. The duration of vaccine immunity is set to 6 months half time. Initial estimate of immunity is simulated on last season, then calculated based on model.
- Subtypes: H1 and H3 are modeled. Type B is not modeled.
- Non-pharmaceutical interventions: NPIs not implemented in the model.

**Contributions:**

- Seasons: 2023-2024, 2024-2025
- Geographies: 51 (iso2code: 01, 02, 04, 05, 06, 08, 09, 10, 12, 13, 15, 16, 17, 18, 19, 20, 21, 22, 23, 24, 25, 26, 27, 28, 29, 30, 31, 32, 33, 34, 35, 36, 37, 38, 39, 40, 41, 42, 44, 45, 46, 47, 48, 49, 50, 51, 53, 54, 55, 56, US)

**Reference paper:**

Bouchnita, A., Bi, K., Fox, S. J. & Meyers, L. A. Projecting Omicron scenarios in the US while tracking population-level immunity. *Epidemics* **46**, 100746 (2024) [12].
Bi, K., Bandekar, S. R., Bouchnita, A., Cramer, A., Fox, S. J., Borchering, R. K., Biggerstaff, M. & Meyers, L. A. Estimated impact of 2022–2023 influenza vaccines on annual hospital burden in the United States. *Proc. Natl Acad. Sci. USA* **122**, e2505175122 (2025) [13].

#### UVA Biocomplexity Institute FluXSim

**Team:** University of Virginia - UVA Biocomplexity Institute (UVA)
**Model:** FluXSim
**Contributors:** Srini Venkatramanan, Aniruddha Adiga, Przemek Porebski, Brian Klahn, Benjamin Hurt, Bryan Lewis, Madhav Marathe

**General Description:**
FluXSim is a metapopulation simulation over age- and spatially stratified synthetic contact network. The model is based on a synthetic contact network at the state level, aggregated to the county level and five age groups.

**Methods:**

- Calibration: Latin Hypercube design is constructed, and particles are evaluated using an exponentially weighted (alpha=0.975) absolute error normalized by similarly weighted ground truth. Fitting is done separately for each scenario and state. Top 50 particles satisfying the peak constraint with the lowest error are selected and combined with hospitalization ratios to create the replicates.
- Non-fixed parameters: Transmissibility is parameterized through a combination of season width, season swing, and season midpoint. These are calibrated with the recent HHS ground truth.
- Age groups: 0-4, 5-17, 18-49, 50-64, 65+.
- Contact matrix/mixing structure: Age and spatial contact matrices obtained from the synthetic population for each state.

**Assumptions:**

- Seasonality: Seasonality modeled as a sinusoidal forcing with calibratable parameters for season duration, dynamic range of transmissibility, and midpoint.
- Vaccine: Vaccine efficacy is applied at infection stage by age group. Vaccine efficacy delay is set to 14 days.
- Immunity: No in-season waning modeled for both natural immunity and vaccine immunity. Initial estimate of immunity is set at 40% for H3 strain and 33% for H1.
- Subtypes: Only influenza A is modeled. Prior immunity settings are different between subtypes. Burden estimates from reference seasons are used to derive age stratified IHR and IFR for H1 and H3. Because age-stratified estimates were similar, used ratio of all age IHR and IFR to scale down H1 numbers.
- Non-pharmaceutical interventions: NPIs not implemented in the model.

**Contributions:**

- Seasons: 2023-2024, 2024-2025
- Geographies:
    - season 2023-2024: 1 (iso2code: US)
    - season 2024-2025: 52 (iso2code: 01, 02, 04, 05, 06, 08, 09, 10, 11, 12, 13, 15, 16, 17, 18, 19, 20, 21, 22, 23, 24, 25, 26, 27, 28, 29, 30,

31, 32, 33, 34, 35, 36, 37, 38, 39, 40, 41, 42, 44, 45, 46, 47, 48, 49, 50, 51, 53, 54, 55, 56, US)

#### UVA Biocomplexity Institute EscapeFlu

**Team:** University of Virginia - UVA Biocomplexity Institute (UVA)
**Model:** EscapeFlu
**Contributors:** Parantapa Bhattacharya, Srini Venkatramanan, Bryan Lewis, Jiangzhuo Chen, Stefan Hoops, Madhav Marathe

**General Description:**
The Epidemic Simulator Compiler and Programming Environment (ESCAPE) is a meta-simulator framework for creating custom high-performance stochastic agent-based contagion simulators. ESCAPE defines the Epidemic Simulator Language (ESL), a domain-specific language for concisely describing such simulators. The 2024-SMH-ESCAPE-Flu model is an agent-based stochastic simulator written in ESL, that simulates an age and spatially stratified Flu contagion model over a synthetic contact network. The contagion model is a compartmental model with eight compartments: Susceptible, Exposed, Infectious (symptomatic), Infectious (asymptomatic), Medical visit, Hospitalized, Dead, and Recovered. The transition and transmission probabilities were parameterized by age, contact duration between individuals, and state-specific seasonality factors.

**Methods:**
- Calibration: The 2024-SMH-ESCAPE-FLu model calibration process used multi-objective Bayesian optimization to fit the following variables for each scenario and each state: baseline transmissibility, seasonality importance for transmissibility, and seeding schedule. The two objectives that were used for calibration were matching attack rate target (25%) and shape of the hospitalization curve. The hospitalization curve targets were obtained from previous year's patient reported hospitalization data.
- Non-fixed parameters: Baseline transmissibility factor (9.06e-6 - 1.09e-4); seasonality factor (2.04e-2 - 8.21e-2); daily seeding probability (9.03e-3 - 2.66e-2).
- Age groups: The model considered the following age groups: 0-4, 5-17, 18-49, 50-64, 65-130.
- Contact matrix/mixing structure: The 2024-SMH-ESCAPE-Flu model used a population digital twin of the whole United States where each individual was a separate agent in the simulation. This digital twin was constructed from many public and proprietary data sources. The contact matrix / mixing structure arose from the physical proximity (close co-location) of the agents

as they came in contact with each other during their daily schedule. The details of this digital twin can be found in [14].

**Assumptions:**

- Seasonality: The 2024-SMH-ESCAPE-Flu model explicitly modeled seasonality by learning a seasonality curve that scaled the transmission probability over the season. This seasonality curve was learned separately for each state and used the FluSurv-NET dataset of the representative seasons (specified in the scenario). For states without sufficient data, the whole U.S. dataset was used to fit the seasonality curve. Specifically, a quadratic version of Model A (Eq. 6) from [15] was used to fit the seasonality curve.
- Vaccine: Vaccine effectiveness was considered in two forms: reduction in susceptibility to infection (30%) and reduction in likelihood of needing medical treatment if symptomatic (as given in the scenario description). Vaccines took effect 14 days after receiving the shot, and its effects waned continuously such that 7% of the gained immunity was lost per month starting at 14 days of receiving the shot.
- Immunity: 30% of the population was considered to have prior immunity for H3 scenarios, whereas 35% of the population was considered prior immune for the H1 scenarios. Prior immune agents where not excluded from receiving vaccine.
- Sybtypes: The model did not consider flu variant subtypes.
- Non-pharmaceutical interventions: The model did not implement any NPIs.

**Contributions:**

- Seasons: 2024-2025
- Geographies: 51 (iso2code: 01, 04, 05, 06, 08, 09, 10, 11, 12, 13, 15, 16, 17, 18, 19, 20, 21, 22, 23, 24, 25, 26, 27, 28, 29, 30, 31, 32, 33, 34, 35, 36, 37, 38, 39, 40, 41, 42, 44, 45, 46, 47, 48, 49, 50, 51, 53, 54, 55, 56, US)

For the 2023-2024 influenza season, eight teams contributed projections from nine models, and in the 2024-2025 round, seven modeling teams contributed projections from eight models. Each model provided from 100 to 300 representative individual trajectories for each scenario and geographic unit, with each trajectory representing a stochastic realization of weekly influenza hospital admissions. Table S1 lists the individual models, the seasons to which they were submitted, and the number of trajectories provided by each.

| Team | Model | # trajs. 2023/2024 for each scenario and geographical resolution | # trajs. 2024/2025 for each scenario and geographical resolution |
|---|---|---|---|
| **CDDEP** | FluCompModel | 100 | / |
| **Fogarty International Center, National Institutes of Health** | FluD | 100 | / |
| | Flu_TS | 100 | 300 |
| **MOBS_NEU** | GLEAM_FLU | 100 | 300 |
| **PSI** | M2 | 100 | 300 |
| **University of Notre Dame** | FRED | 100 | 200 |
| **University of Southern Carolina** | SIkJalpha | 100 | 100 |
| **UT** | ImmunoSEIRS | 100 | 100 |
| **UVA Biocomplexity Institute** | EscapeFlu | / | 120 |
| | FluXSim | 100 | 100 |

**Table S1: Number of trajectories submitted to the Scenario Modeling Hub for the 2023-2024 and 2024-2025 influenza season.** The table reports the number of trajectories submitted by each model for each scenario and geographical resolution.

### S2. Specifications and assumptions for Round 1 of seasons 2024-2025 and 2023-2024 of the U.S. Flu Scenario Modeling Hub

#### S2.1 Epidemic scenarios

Round 1 of season 2024-2025 of the U.S. Flu Scenario Modeling Hub provided early season projections for incident influenza hospitalizations in the United States at both the national and subnational levels. Six scenarios describing the interaction of the levels of vaccination coverage based on vaccination rates reported in 2022-2023 (first dimension) with the dominant influenza A subtype (second dimension) considered as follows:

| | **Season dominated by influenza A/H3N2, natural history indexed on 2017-18 season.** | **Season dominated by influenza A/H1N1, natural history indexed on 2019-20 season.** |
|---|---|---|
| **Higher than Usual Vaccine Coverage**<br>● Vaccine coverage is **20% higher than in the 2022-23** flu season in all age groups and jurisdictions. (ie coverage for age group *a* and jurisdiction *j* is 1.20x the coverage for 2023-23). Overall, the US coverage is about 58% in this scenario. | **Scenario A** | **Scenario B** |
| **Business as Usual Vaccine Coverage**<br>● Vaccine coverage is **the same as in the 2022-23** flu season in all age groups and jurisdictions. Overall, the US coverage is about 49% in this scenario. | **Scenario C** | **Scenario D** |
| **Low Vaccine Coverage**<br>● Vaccine coverage is **20% lower than in the 2022-23** flu season in all age groups and jurisdictions (x0.80 coverage in 2022-23). Overall, the US coverage is about 39% in this scenario. | **Scenario E** | **Scenario F** |

Similarly, Round 1 of the 2023-2024 season focused on early-season projections of hospitalization trends, using comparable assumptions. The six scenarios were based on 2021-2022 vaccination rates and the expected dominant influenza A subtype, as follows:

| | **Season dominated by influenza A/H3N2, indexed on age distribution of 2017-18 season.**<br><br>**VE = 40% against medically attended illnesses and hospitalizations, VE drops in older age groups** | **Season dominated by influenza A/H1N1, indexed on age distribution of 2019-20 season.**<br><br>**VE = 40% against medically attended illnesses and hospitalizations, similar VE across all age groups** |
|---|---|---|
| **Higher than Usual Vaccine Coverage**<br>● Vaccine coverage is **20% higher than in the 2021-22** flu season in all age groups and jurisdictions. (20% is a relative change, ie a 50% coverage for age group *a* and jurisdiction *j* in 2021-22 translates to a 50%*1.20=60% coverage for 2023-24). Overall, the US coverage is about 60% in this scenario. | **Scenario A** | **Scenario B** |
| **Business as Usual Vaccine Coverage**<br>● Vaccine coverage is **the same as in the 2021-22** flu season in all age groups and jurisdictions. Overall, the US coverage is about 50% in this scenario. | **Scenario C** | **Scenario D** |
| **Low Vaccine Coverage**<br>● Vaccine coverage is **20% lower than in the 2021-22** flu season in all age groups and jurisdictions. Overall, the US coverage is about 40% in this scenario. | **Scenario E** | **Scenario F** |

#### S2.2. Projection period and geographic scope

For the 2024-2025 season, projections considered a 43-week period, running August 11, 2024, to June 7, 2025. Projections for all states and territories were not required, but projections for the whole United States were recommended.

For the 2023-2024 season, projections covered a 39-week period, running September 3, 2023, to June 1, 2024. Although national projections were recommended, state and territorial projections were optional.

#### S2.3 Targets

Targets for Round 1 of season 2023-2024 were overall and age-specific (recommended age strata: 0-4, 5-17, 18-49, 50-64, and 65+) weekly incident influenza hospitalizations (based on HHS COVID and flu reporting system) and weekly influenza deaths (from the CDC multiplier model). Teams were required to send their projections in the form of individual stochastic trajectories (100 to 300 for each model) for each target, geography, and population segment considered. The submission of quantiles in addition to trajectories was optional.

Teams were also given the opportunity to submit additional optional targets (only if submitting quantiles in addition to trajectories):

- Cumulative hospitalizations
- State-level peak hospitalizations
- State-level timing of peak hospitalizations

#### S2.4 Assumptions regarding influenza subtype dominance, severity and age dynamics

The age dynamics and severity of the flu season were indexed based on two exemplar seasons dominated by A/H3N2 (2017-18) and A/H1N1 (2019-20). Nonetheless, teams were not required to assume that the timing of the projected 2024-2025 and 2023-2024 seasons could not vary with respect to the dynamics of the exemplary seasons. In other words, teams were left the opportunity to project early epidemics if model assumptions led to that because of factors such as prior immunity, seeding, or contact assumptions. More in detail:

1. The three scenarios assuming A/H3N2 dominance (A, C, E) were based on epidemiologic patterns observed during the 2017-2018 season. Teams were advised to use data from this season (including but not limited to ILI, percentage of positives for influenza, FluSurv-NET hospitalizations) to estimate the effective reproductive number ($R_e$) at the start of the season and the fraction of hospitalizations in the different age groups. For reference, around 65% of observed hospitalizations in this season were among seniors and 6% were in children [16].

2. The three scenarios assuming A/H1N1 dominance (B, D, F) were based on the 2019-2020 season. Similarly, teams were advised to base the overall burden and fraction of hospitalizations in the different age groups on data from this season. For reference, during the 2019-20 epidemic, around 40% of observed hospitalizations were among seniors and around 15% of hospitalizations were observed in children [17]. Additionally, teams were asked to disregard the potential effect of COVID-19 at the very tail end of the 2019-20 season.

The inclusion of Influenza B activity (observed to some extent in both exemplar seasons) in these considerations was left at the discretion of participating teams.

#### S2.5 Assumptions regarding vaccine effectiveness

Teams were required to assume an all-age vaccine effectiveness (VE) of 40% against medically attended influenza illnesses and hospitalizations, in line with evidence from the 2017-18 and 2019-20 exemplar seasons. Teams having developed age-stratified models were given the option to use the age-specific VE reported for these two seasons. Assumptions about VE against infection and transmission were left at the teams' discretions.

#### S2.6 Assumptions regarding vaccine coverage

Weekly state-level coverage to be used in scenarios A-F was provided for each of the following age groups: 0-4yr, 5-12yr, 13-17yr, 0-17yr, 18-49yr, 50-64yr, 65+yr. These estimates were based on influenza vaccination rates reported in 2022-2023 or 2021-2022 for business-as-usual scenarios (C, D), with 20% correction up (scenarios A, B) and down (scenarios E, F) to match scenario assumptions. The 20% changes were applied to each age group and jurisdiction.

#### S2.7 Other assumptions

**Prior Immunity.** Prior influenza immunity was assumed to be a combination of residual immunity from previous infections and previous seasonal vaccinations. The exact specifications of prior immunity were left at the discretion of each team, depending on model structure, but the following suggestions were provided:

- At the onset of a typical influenza season (all subtypes combined), modeling has estimated that around 30-35% of the population has prior immunity (i.e., 65-70% susceptible), the effective reproduction number ranges from 1.2-1.4, and the attack rate is between 8-25% [9]. The 2009 pandemic, which was marked by the emergence of a new strain to which individuals under the age of 50 years were susceptible, was associated with greater transmission (cumulative attack rates 32% over 2009) and decreased prior immunity compared to a regular season (prior immunity in 2009 was ~25% instead of ~33%).
- Teams were allowed to vary prior immunity by virus subtype, age, or other demographic characteristics, and state.

**COVID-19 Interactions.** No major interactions with future COVID-19 surges (immunological, social, behavioral) were considered in this round.

**Influenza strains.** Hospitalization and death targets were intended to include the impact of all influenza subtypes combined. Influenza A/H3N2 was presumed to dominate in scenarios A, C, E and A/H1N1 in scenarios B, D, F. Subtype-specific models were allowed. Intrinsic transmissibility (i.e., $R_0$) was assumed to be the same independently from the season, and the only changes in effective transmission of either A/H3N2 or A/H1N1 were assumed to originate from differences in prior immunity and social mixing.

**Vaccine immune waning.** Teams were advised to include waning of vaccine-induced protection, as vaccine-induced immunity has been found to decrease rapidly over the course of an influenza season [18].

**Age groups**. Age-stratification was optional. Suggested age-strata were 0-4, 5-17, 18-49, 50-64, and 65+.

**State-level variability.** Differences in the dynamics in different states and territories were assumed to originate from different vaccination coverage and demographic. Variability in the age distribution of hospitalizations between states was permitted, provided it aggregated to match the scenario definition for the United States as a whole. Additionally, variability in severity and prior immunity between states was allowed.

**Cross-protection of subtype immunity.** Cross-protection of subtype immunity was left at the discretion of the teams.

**Seasonality.** Teams were left free to include their best estimates of influenza seasonality in their model, no specific level of seasonal forcing was prescribed.

**Non-pharmaceutical interventions (NPIs).** No reactive NPIs to COVID-19 or influenza were assumed, inclusion of low-level masking was allowed at teams' discretion.

**Seeding of influenza.** Seeding intensity, timing, and geographic distribution was left at the discretion of the teams.

**Initial conditions.** The mix of circulating strains at the start of the projection period was left at the discretion of the teams based on their interpretation of the scenarios. Variation in initial prevalence between states was also left at the teams' discretion.

#### S3. Summary of individual models that submitted short-term forecasts to FluSight during the 2024-2025 and 2023-2024 seasons

The FluSight forecasting hub is a short-term forecasting challenge coordinated by the CDC, in which modeling teams submit weekly forecasts of confirmed influenza hospitalizations, typically from October through May. During the 2023-2024 season, 38 models participated across more than 30 forecasting rounds; during the 2024-2025 season, 50 models participated across 27 rounds. Table S2 lists the individual contributing models and the number of rounds covered by each during the respective season.

| Model | # rounds 2023/2024 (2023-10-14 to 2024-05-05) | # rounds 2024/2025 (2024-11-23 to 2025-05-28) |
|---|---|---|
| **CEPH-Rtrend_fluH** | 30 | 26 |
| **CFA_Pyrenew-Pyrenew_HE_Flu** | / | 11 |
| **CFA_Pyrenew-Pyrenew_H_Flu** | / | 19 |
| **CMU-TimeSeries** | 28 | 26 |
| **CMU-climate_baseline** | / | 25 |
| **CU-ensemble** | 30 | 25 |
| **FluSight-baseline** | 30 | 26 |
| **FluSight-ensemble** | 30 | 26 |
| **FluSight-lop_norm** | 30 | 26 |
| **FluSight-trained_mean** | / | 20 |
| **FluSight-trained_med** | / | 20 |
| **Gatech-ensemble_point** | / | 26 |
| **Gatech-ensemble_prob** | / | 26 |
| **GH-model** | 9 | / |
| **GT-FluFNP** | 27 | / |
| **Google_SAI-FluBoostQR** | / | 3 |
| **ISU_NiemiLab-GPE** | / | 22 |
| **ISU_NiemiLab-ENS** | 18 | / |
| **ISU_NiemiLab-NLH** | 25 | / |
| **ISU_NiemiLab-SIR** | 18 | / |
| **JHUAPL-DMD** | / | 24 |
| **JHUAPL-Morris** | / | 6 |
| **JHU_CSSE-CSSE_Ensemble** | 17 | 21 |
| **LUcompUncertLab-chimera** | 30 | 25 |
| **LosAlamos_NAU-CModel_Flu** | 28 | 21 |

| | | |
|---|---|---|
| **MDPredict-SIRS** | / | 25 |
| **MIGHTE-Joint** | / | 22 |
| **MIGHTE-Nsemble** | 30 | 24 |
| **MOBS-GLEAM_FLUH** | 30 | 25 |
| **Metaculus-cp** | / | 23 |
| **NEU_ISI-AdaptiveEnsemble** | / | 25 |
| **NEU_ISI-FluBcast** | / | 24 |
| **NIH-Flu_ARIMA** | 26 | 26 |
| **OHT_JHU-nbxd** | / | 25 |
| **NU_UCSD-GLEAM_AI_FLUH** | 23 | / |
| **PSI-PROF** | 30 | 27 |
| **PSI-PROF_beta** | 20 | 25 |
| **SGroup-RandomForest** | 29 | / |
| **SigSci-CREG** | 30 | / |
| **SigSci-TSENS** | 30 | 27 |
| **Stevens-GBR** | 29 | / |
| **Stevens-ILIForecast** | / | 21 |
| **UGA_CEID-Walk** | / | 24 |
| **UGA_flucast-Copycat** | 26 | 23 |
| **UGA_flucast-INFLAenza** | 27 | 26 |
| **UGA_flucast-OKeeffe** | 3 | / |
| **UGA_flucast-Scenariocast** | / | 22 |
| **UGuelph-CompositeCurve** | 28 | 14 |
| **UGuelphensemble-GRYPHON** | 27 | 16 |
| **UI_CompEpi-EpiGen** | / | 21 |
| **UM-DeepOutbreak** | 30 | 26 |
| **UMass-AR2** | / | 25 |
| **UMass-flusion** | 30 | 25 |
| **UMass-trends_ensemble** | 30 | 27 |
| **UNC_IDD-InfluPaint** | 24 | 21 |

| | | |
|---|---|---|
| **UVAFluX-CESGCN** | / | 20 |
| **UVAFluX-Ensemble** | 29 | 27 |
| **UVAFluX-OptimWISE** | / | 9 |
| **VTSanghani-Ensemble** | 26 | / |
| **VTSanghani-PRIME** | / | 27 |
| **cfa-flumech** | 28 | / |
| **cfarenewal-cfaepimlight** | 30 | / |
| **fjordhest-ensemble** | 30 | 4 |

**Table S2: Overview of models contributing to the FluSight challenge for the 2023-2024 and 2024-2025 influenza season.** The table reports the models submitting short-term forecasts and the number of rounds covered by each model during the respective season.

### S4. Scoring rules for performance evaluation

#### S4.1 Loss functions

To ensure the robustness of results presented in the main text, we perform trajectory selection for scenario modeling using two standard loss functions: Weighted Mean Absolute Percentage Error (presented in the main text) and the Root Mean Squared Error.

##### S4.1.1 Weighted Mean Absolute Percentage Error

The Weighted Mean Absolute Percentage Error (wMAPE) is a variation of the Mean Absolute Percentage Error (MAPE) where errors are weighted by the actual values or weights, unlike MAPE, which calculates errors based on average values. The wMAPE was proposed for cases where there are values close to zero in the data. In such instances, the MAPE suffers from division by zero issues and can therefore yield infinite values. The output of the wMAPE metric is non-negative, and the optimal value is 0. Mathematically, the wMAPE is given by the formula:

$$wMAPE = \frac{\sum_{t=1}^{T} |h_t - \widehat{h}_t|}{\sum_{t=1}^{T} h_t} \times 100, \quad (1)$$

where $\widehat{h_t}$ is the projection value and $h_t$ is the actual value.

##### S4.1.2 Root Mean Squared Error

The Root Mean Squared Error (RMSE) is the quadratic mean of the differences between the observed values and predicted ones. RMSE is always non-negative, and a value of 0 indicates a perfect fit to the data. Mathematically, the RMSE is given by the formula:

$$RMSE = \sqrt{\frac{\sum_{t=1}^{T}(h_t - \widehat{h_t})^2}{T}}, \qquad (2)$$

where $\widehat{h_t}$ is the projection value and $h_t$ is the actual value.

##### S4.2 Jaccard Similarity Index

We use the Jaccard Similarity Index (JSI) [19] to measure the persistence of selected trajectories over time, evaluating how much the pool of selected trajectories changes across time steps. This index measures the similarity between two sets. Let $A = T_{t_1}$ and $B = T_{t_2}$ denote the pools of trajectories selected at time steps $t_1$ and $t_2$, respectively. The JSI is defined as the ratio between the size of the intersection of two sets of selected trajectories and the size of their union: $JSI(A,B) = \frac{|A \cap B|}{|A \cup B|}$.

#### S5. Evaluating adaptive ensemble for scenario modeling projections

##### S5.1 Comparison with individual scenario ensembles

Fig. S1 and Fig. S2 compare the performance of the adaptive ensemble with ensembles computed independently for individual scenarios A, B, C, D, E, F as well as the original ensemble[2], which combines trajectories from all models and scenarios (the benchmark used in the main text), for the long-term scenario projection modeling task of the U.S. Flu Scenario Modeling Hub Round 1 for the 2024-2025 and 2023-2024 season.

In Fig. S1A, we present the ratio of the WIS of the adaptive ensemble to that of the original, static ensembles, over all weeks on which the adaptive ensemble has been regenerated for season 2024-2025. Similar to the results presented in Fig. 2 in the main text, values below 1 indicate that the adaptive ensemble performs better than the original one in terms of WIS, and values higher than 1 indicate worse performance. The adaptive ensemble achieves median WIS values below 1 across all individual scenarios and all percentages of selected trajectories, with two exceptions: it performs worse than Scenario E at all percentages, and worse than Scenario F at the 25%, 50%, and 75% level. Fig. S1B extends the analysis by evaluating performance using the MAE of the median instead of the WIS. In this case, the adaptive ensemble outperforms all the individual scenario ensembles, showing median MAE ratios below

1 across all percentages of trajectory selection. In Fig. S2A, we report the WIS ratios for the 2023-2024 season. As with the 2024-2025 results, the adaptive ensemble shows improved performance in most configurations, except for the 25%, 50%, and 75% cases in Scenario A. Fig. S2B shows the corresponding MAE ratios, indicating that the adaptive ensemble outperforms the original ensembles across all scenarios and trajectory selection percentages, except for the 5% case in Scenario A and Scenario F.

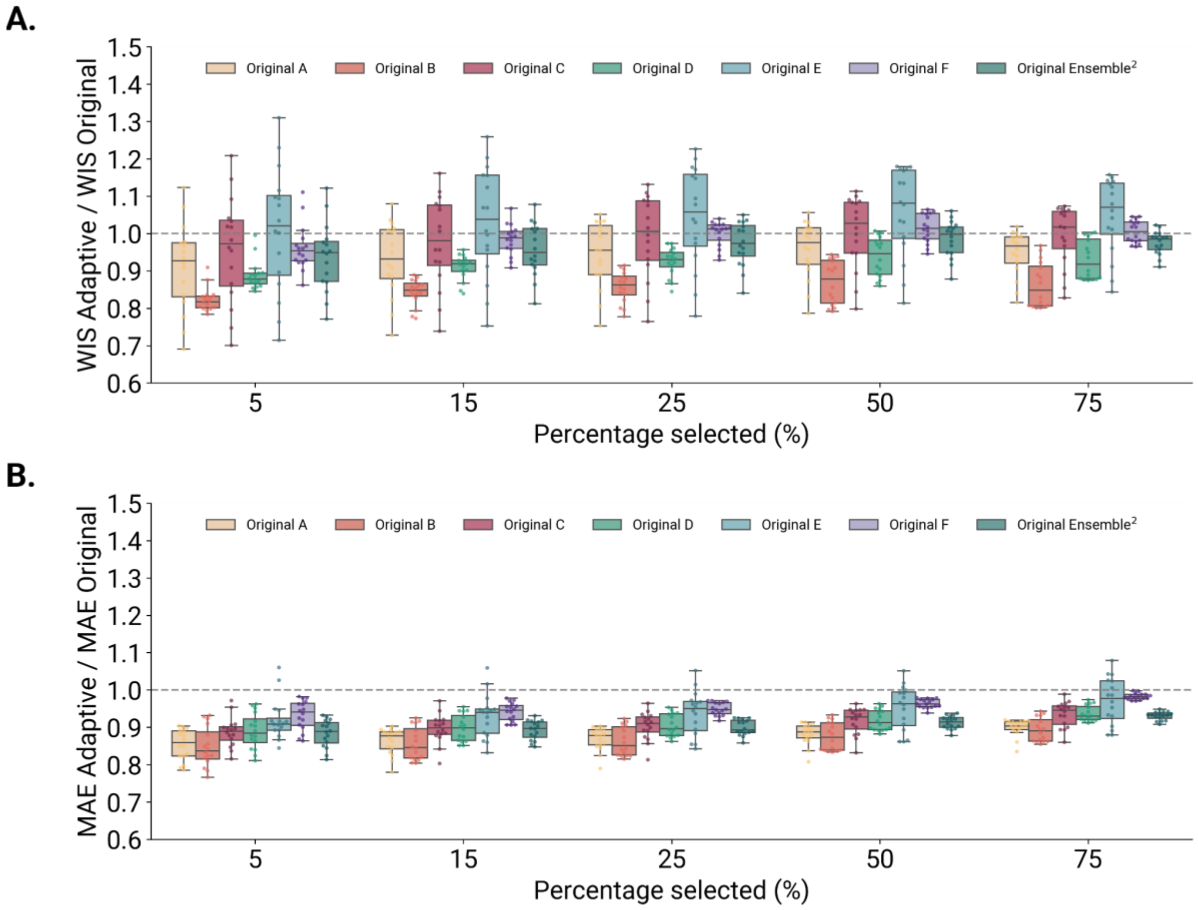


**Figure S1. Performance of the adaptive ensemble model in generating US-level projections (US Flu Scenario Modeling Hub Round 1, season 2024-2025).** A) Ratio between the WIS of the adaptive ensemble and of different original ensembles across five percentages of the trajectories selected and over all weeks. B) Ratio between the MAE of the median of the adaptive ensemble and of different original ensembles across five percentages of the trajectories selected and over all weeks. Specifically, we evaluate the performance of the adaptive ensemble against the single scenarios B, D, and F, and the original ensemble pooling together all individual scenarios. A ratio below 1 indicates better performance of the adaptive ensemble with respect to the original ensemble. Each boxplot is based on 16 data points, corresponding to the weeks in which the adaptive ensemble was generated. The overlaid swarmplot points correspond to each week and the square markers indicate the mean value. The boxplot boundaries represent the interquartile range (IQR) between the first quartile (Q1) and third quartile (Q3), and the line inside each box indicates the median. The whiskers extend to the furthest data point within 1.5 times the IQR from Q1 and Q3.

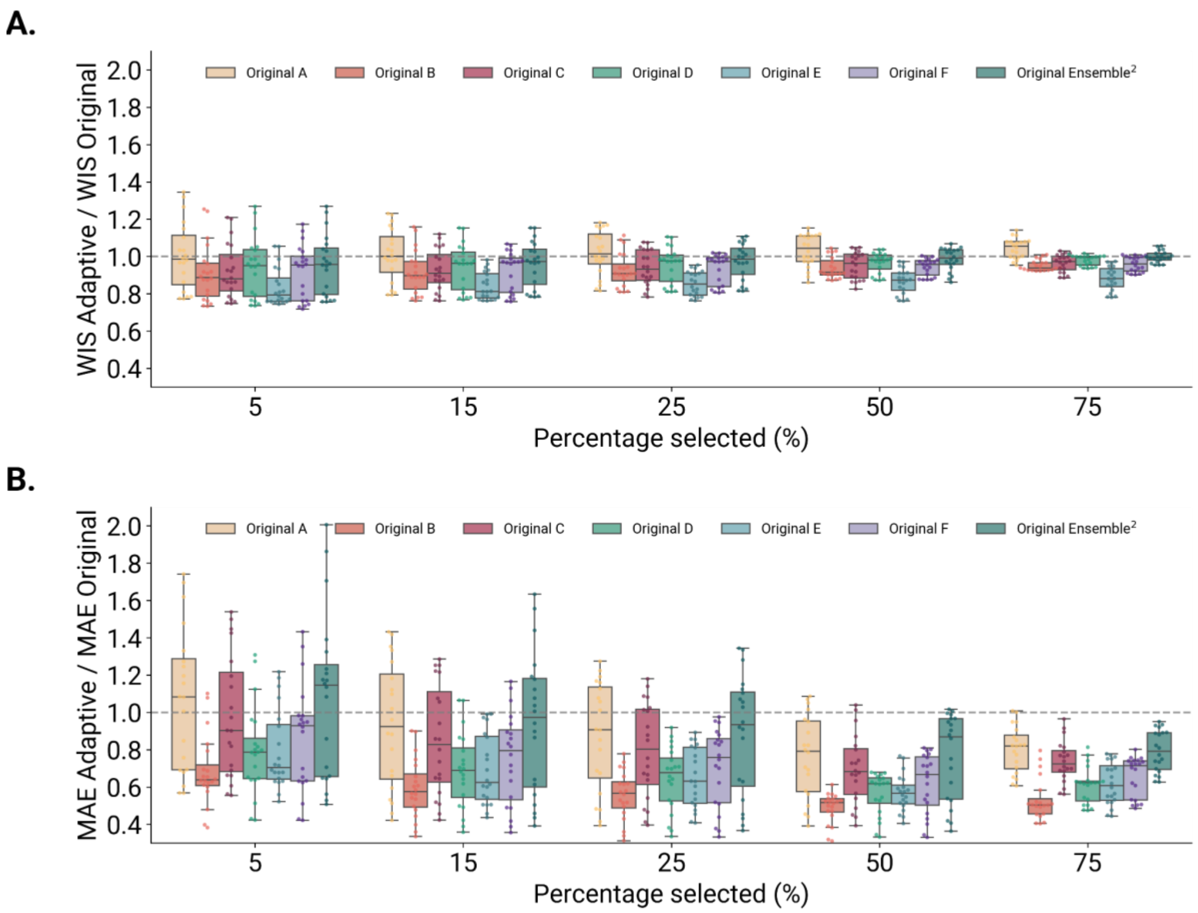


**Figure S2. Performance of the adaptive ensemble model in generating US-level projections (US Flu Scenario Modeling Hub Round 1, season 2023-2024).** A) Ratio between the WIS of the adaptive ensemble and of different original ensembles across five percentages of the trajectories selected and over all weeks. B) Ratio between the MAE of the median of the adaptive ensemble and of different original ensembles across five percentages of the trajectories selected and over all weeks. Specifically, we evaluate the performance of the adaptive ensemble against the single scenarios B, D, and F, and the original ensemble pooling together all individual scenarios. A ratio below 1 indicates better performance of the adaptive ensemble with respect to the original ensemble. Each boxplot is based on 20 data points, corresponding to the weeks in which the adaptive ensemble was generated. The overlaid swarmplot points correspond to each week and the square markers indicate the mean value. The boxplot boundaries represent the interquartile range (IQR) between the first quartile (Q1) and third quartile (Q3), and the line inside each box indicates the median. The whiskers extend to the furthest data point within 1.5 times the IQR from Q1 and Q3.

#### S5.2 Performance analysis for the 2023-2024 season

In Fig. S3, we repeat the analysis described in the manuscript for the 2023-2024 season, reporting the ratio between WIS and MAE of the adaptive and the original ensemble$^2$, respectively (Fig. S3A and Fig. S3B). We observe that, overall, the adaptive ensemble outperforms the original ensemble$^2$ at several weekly data releases, showing the median WIS ratio lower than 1 in all the tested percentages. Also, the median MAE ratio is smaller than 1 for all percentages considered, except 5%, showing a median value slightly higher than 1 (1.14). For easier interpretation of results, we report in Table S3 the median and interquartile (IQR) range of WIS and MAE ratios across different percentages of trajectories selected.

In Fig. S3C and Fig. S3D, we present the WIS and MAE ratios, respectively, comparing the adaptive ensemble to the original ensemble[2] over time for the 2023-2024 season. In the first time step, the WIS ratio exceeds 1 in the 15% case of selected trajectories. However, after this first step, WIS ratios drop below 1 for all three cases, 15%, 25%, and 50%. The adaptive ensemble does not show improvements in proximity of the peak of weekly influenza hospitalizations. Around the peak the WIS ratios increase, surpassing 1 for six and nine consecutive weeks in the 15% and 50% trajectories selected, respectively. The adaptive ensemble is then outperforming the ensemble[2] in the entire declining phase of the seasonal epidemic. For MAE ratios, a similar pattern is observed. Initially, the values are below 1, but they rise above 1 during the upward trend in incident influenza hospitalizations, persisting for ten consecutive weeks in the 15% case, and remain consistently below 1 for the 50% case except for two weeks. Subsequently, MAE ratios drop below 1 across all percentages considered, with a slight decline in performance observed toward the tail end of the epidemic.

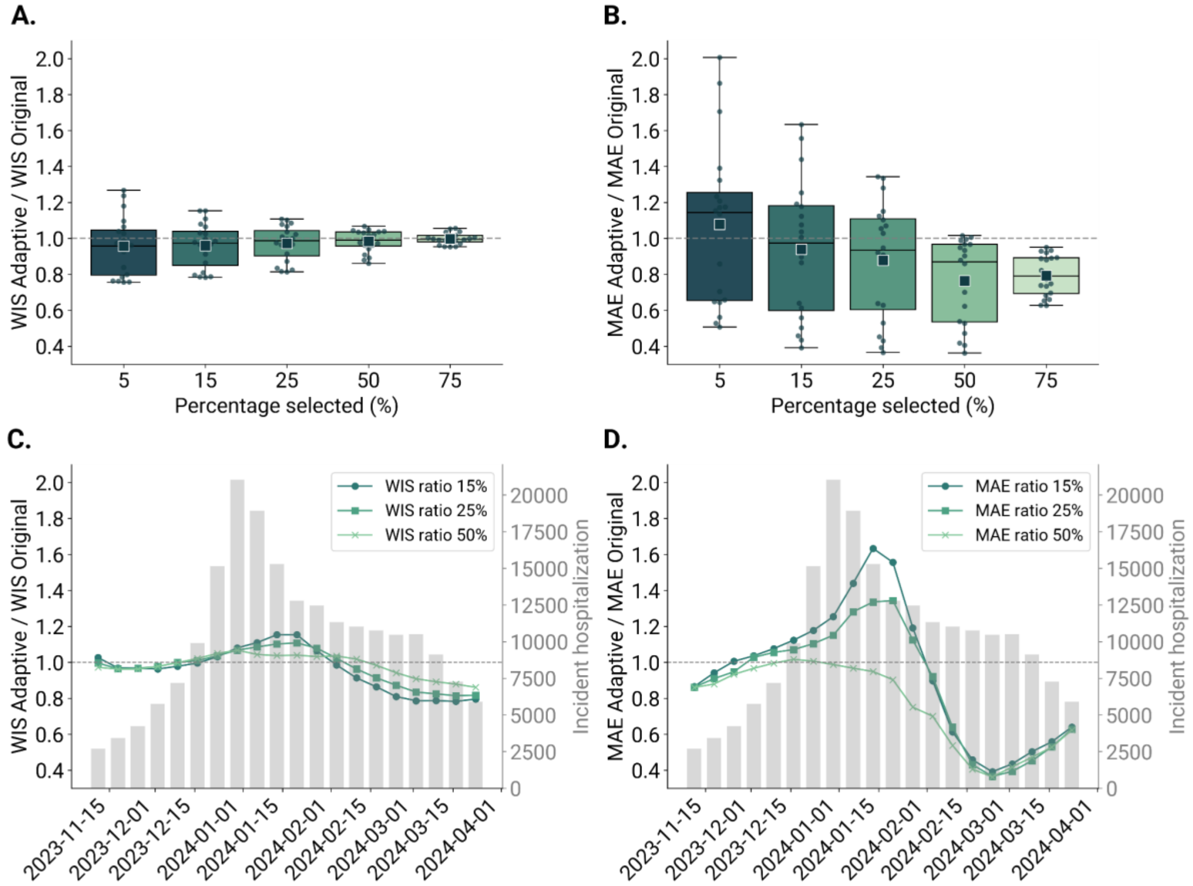


**Figure S3. Performance of the adaptive ensemble model on US-level projections from the U.S. Flu Scenario Modeling Hub Round 1, season 2023-2024.** A) Ratio between the WIS of the adaptive ensemble and of the original ensemble[2] across different percentages of trajectory selected and over all weeks. B) Ratio between the MAE of the median of the adaptive ensemble and of the original ensemble[2] across different percentages of trajectory selected and over all weeks. A ratio below 1 indicates better performance of the adaptive ensemble with respect to the original ensemble[2]. Each boxplot is based on 20 data points, corresponding to the weeks in which the adaptive ensemble was generated. The overlaid swarmplot points correspond to each week and the square markers indicate the mean value. The boxplot boundaries represent the interquartile range (IQR) between the first quartile (Q1) and third quartile (Q3), and the line inside each box indicates the median. The whiskers extend to the furthest data point within 1.5 times the IQR from Q1 and Q3. C) Ratio between the WIS of the adaptive ensemble and of the original ensemble[2] across selected percentages over time. D) Ratio between the MAE of the median of the adaptive ensemble and of the original ensemble[2] across selected percentages over time. In the background, the reported weekly incident influenza hospitalizations are shown [20].

| **Percentage**<br>*% of selected trajectories* | **WIS ratio**<br>*Median (IQR)* | **MAE ratio**<br>*Median (IQR)* |
|---|---|---|
| 5% | **0.96 (0.80-1.05)** | *1.14 (0.66-1.26)* |
| 15% | 0.97 (0.85-1.04) | 0.97 (0.60-1.18) |
| 25% | 0.99 (0.90-1.04) | 0.93 (0.60-1.11) |
| 50% | 0.99 (0.96-1.04) | 0.87 (0.54-0.97) |
| 75% | 0.99 (0.98-1.02) | **0.79 (0.69-0.89)** |

**Table S3: Performance summary of the adaptive ensemble model on US-level projections from the U.S. Flu Scenario Modeling Hub Round 1, season 2023-2024.** The table reports the median and interquartile range (IQR) of WIS and MAE of the median ratios for each percentage of trajectory selected. In bold, the best WIS and MAE ratio median values for the selected percentages; in italics, WIS and MAE ratio median values higher than 1. Values below 1 indicate better performance of the adaptive ensemble with respect to the original ensemble[2].

#### S5.3 Sensitivity analysis to the loss function considered for trajectory selection

In Figs. S4-S5, we present the ratio between the WIS and MAE of the adaptive ensemble and the original ensemble[2] for the 2024-2025 and 2023-2024 seasons, respectively, using the RMSE as the loss function to select trajectories instead of the wMAPE. This analysis was conducted to test the robustness of our findings under an alternative loss function for trajectory selection. The picture findings are analogous to those presented in the main text, with the adaptive ensemble consistently demonstrating better performance in median terms across all tested percentages according to both WIS and MAE of the median, and in several configurations.

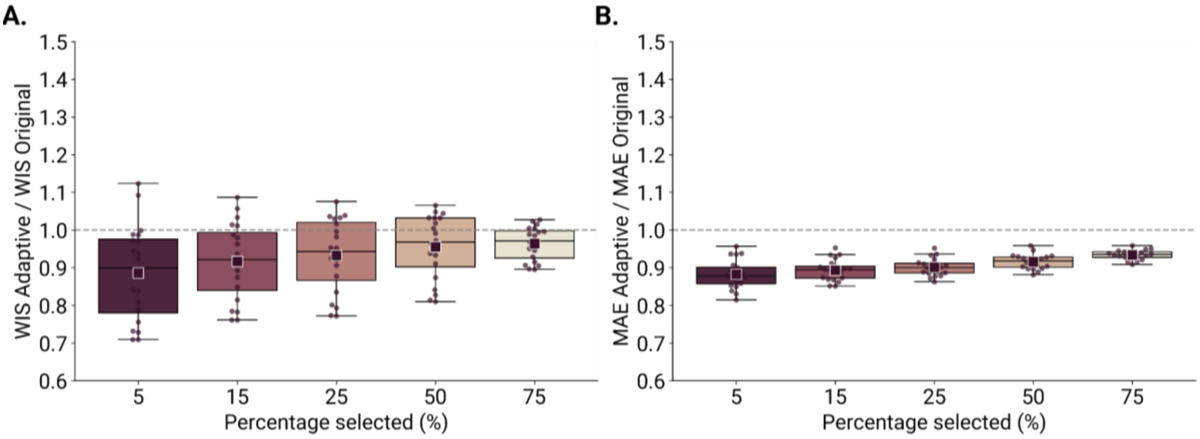


**Figure S4. Performance of the adaptive ensemble model on US-level projections from the U.S. Flu Scenario Modeling Hub Round 1, season 2024-2025.** The adaptive ensemble approach has been tested using Root-Mean-Squared Error (RMSE) as the loss function to select trajectories**.** A) Ratio between the WIS of the adaptive ensemble and of the original ensemble across different percentages of trajectory selected and over all weeks. B) Ratio between the MAE of the median of the adaptive ensemble and of the original ensemble across different percentages of trajectory selected and over all weeks. A ratio below 1 indicates better performance of the adaptive ensemble with respect to the original ensemble. Each boxplot is based on 16 data points, corresponding to the weeks in which the adaptive ensemble was generated. The overlaid swarmplot points correspond to each week and the square markers indicate the mean value. The boxplot boundaries represent the interquartile range (IQR) between the first quartile (Q1) and third quartile (Q3), and the line inside each box indicates the median. The whiskers extend to the furthest data point within 1.5 times the IQR from Q1 and Q3.

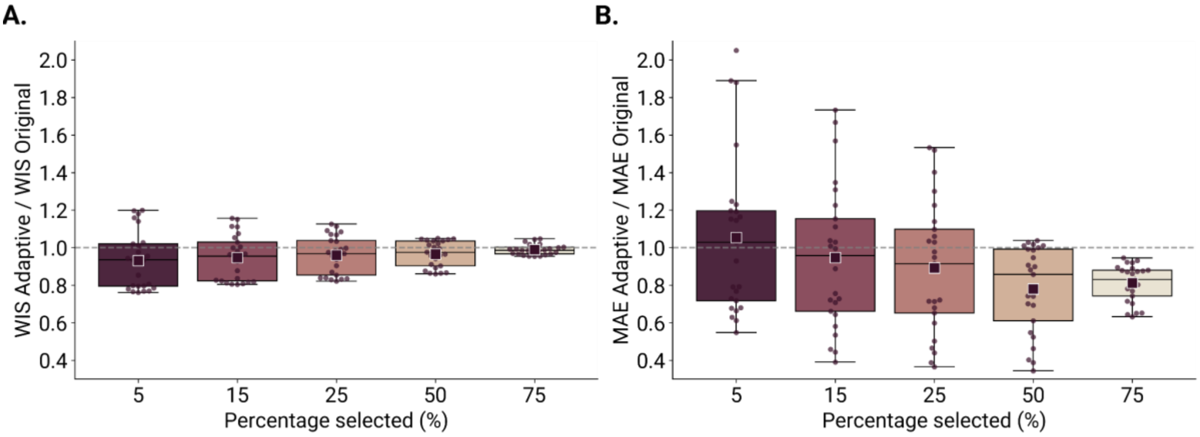


**Figure S5. Performance of the adaptive ensemble model on US-level projections from the U.S. Flu Scenario Modeling Hub Round 1, season 2023-2024.** The adaptive ensemble approach has been tested using Root-Mean-Squared Error (RMSE) as the loss function to select trajectories. A) Ratio between the WIS of the adaptive ensemble and of the original ensemble across different percentages of trajectory selected and over all weeks. B) Ratio between the MAE of the median of the adaptive ensemble and of the original ensemble across different percentages of trajectory selected and over all weeks. A ratio below 1 indicates better performance of the adaptive ensemble with respect to the original ensemble. Each boxplot is based on 20 data points, corresponding to the weeks in which the adaptive ensemble was generated. The overlaid swarmplot points correspond to each week and the square markers indicate the mean value. The boxplot boundaries represent the interquartile range (IQR) between the first quartile (Q1) and third quartile (Q3), and the line inside each box indicates the median. The whiskers extend to the furthest data point within 1.5 times the IQR from Q1 and Q3.

#### S5.4 Temporal trend of performance for additional percentages of trajectories selected

In Fig. S6, we extend the analysis of WIS and MAE ratios over time presented in the manuscript for the 2024-2025 season, comparing the adaptive ensemble to the original ensemble for the 5% and 75% trajectory selection cases. For the 5% case, we observe more pronounced deviations from a ratio of 1, with both positive and negative fluctuations, reflecting the higher variability introduced by selecting a smaller subset of trajectories. Conversely, in the 75% case, the ratio values remain consistently closer to 1 with reduced fluctuations over time. Similarly, in Fig. S7, we report WIS and MAE ratios over time for 2023-2024 season and the 5% and 75% trajectory selection cases. Again, for the 5% case, we observe higher variability in ratio values, whereas in the 75% case, the ratio values remain consistently closer to 1 with reduced fluctuations over time.

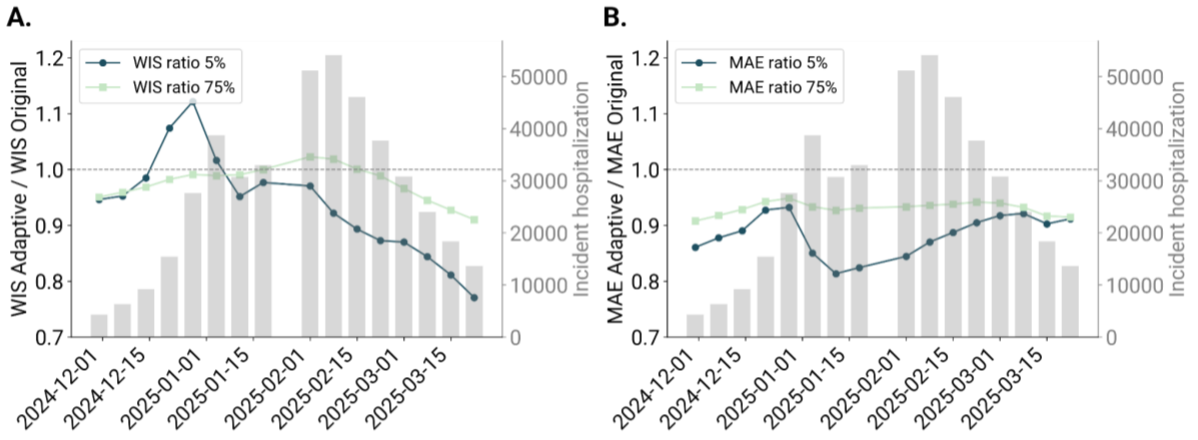


**Figure S6. Performance in time of the adaptive ensemble model in generating US-level projections (US Flu Scenario Modeling Hub Round 1, season 2024-2025).** Results are shown for 5%, and 75% of trajectories selected and for two key metrics: (A) the WIS ratio and (B) the MAE of the median ratio between adaptive and original ensembles. In the background, the reported incident hospitalization [20].

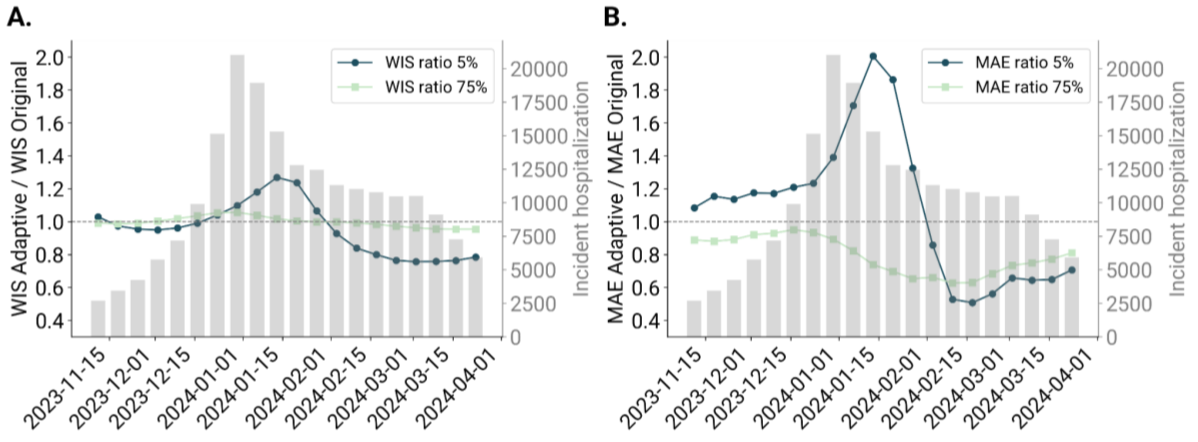


**Figure S7. Performance in time of the adaptive ensemble model in generating US-level projections (US Flu Scenario Modeling Hub Round 1, season 2023-2024).** Results are shown for 5%, and 75% of trajectories selected and for two key metrics: (A) the WIS ratio and (B) the MAE of the median ratio between adaptive and original ensembles. In the background, the reported incident hospitalizations [20].

#### S5.5 Sensitivity analysis on different numbers of trajectories across individual models

Here, we present a sensitivity analysis to explore how different numbers of submitted trajectories influence each model's contribution to the ensemble. In the adaptive ensemble, the filtering procedure selects the top X% of trajectories from each model independently. As a consequence, when all models submit the same number of trajectories, they are equally represented in the adaptive ensemble. For the 2023-2024 season, each model contributed exactly 100 trajectories for every scenario and geographic unit, and they were equally represented. In contrast, during the 2024-2025 season, the Scenario Modeling Hub allowed models to submit up to 300 trajectories, leading to variability in the number of trajectories across models. As a result, models submitting a larger number of trajectories are more represented in the ensemble, because a larger pool is selected. To address the concern of different model representations, we conducted a sensitivity analysis using a bootstrapping approach. Specifically, for models that submitted more than the minimum number of trajectories, we randomly downsampled their trajectories so that all models contributed an equal number. We then generate a static ensemble and apply the adaptive ensemble approach on the new downsampled set. The procedure has been repeated 10 times to ensure robustness. In Fig. S8, we present the WIS and MAE ratios with respect to the original static ensemble. The results show that the performance improvements of the adaptive ensemble relative to the original ensemble remain consistent even after downsampling, with a similar pattern of decreasing in WIS performance for higher thresholds (50% and 75%), and more stable improvements for MAE. This indicates that our findings are not driven by differences in the number of submitted trajectories across models.

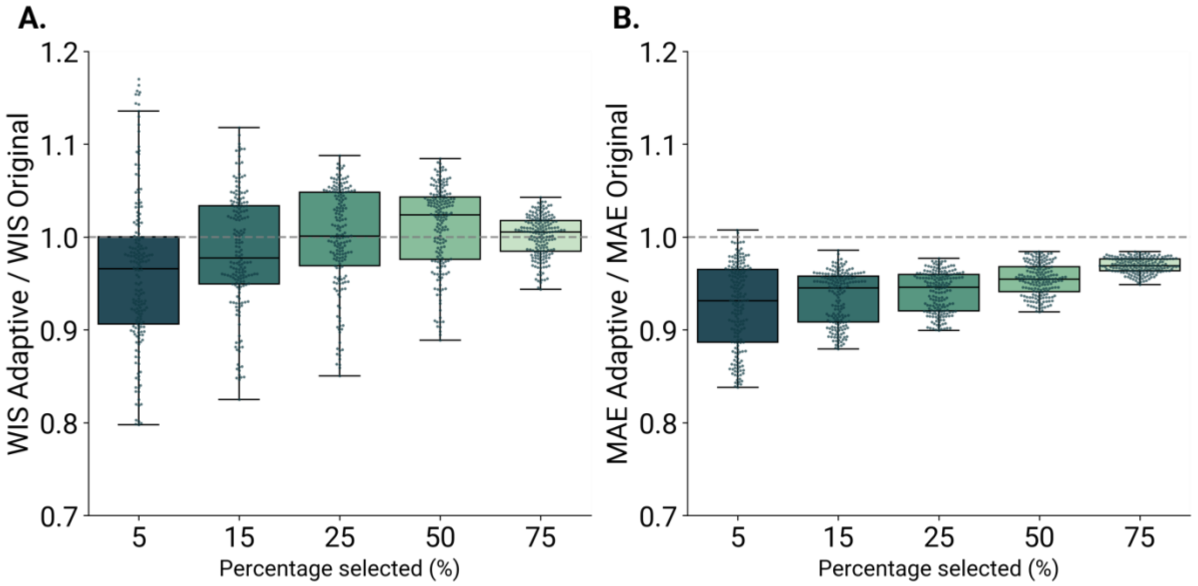


**Figure S8. Performance of the adaptive ensemble model on US-level projections from the U.S. Flu Scenario Modeling Hub Round 1, season 2024-2025.** The adaptive ensemble approach has been tested using a bootstrapping approach to downsample trajectories to equal numbers across models (10 runs have been performed). A) Ratio between the WIS of the adaptive ensemble and of the original ensemble across different percentages of trajectory selected and over all weeks. B) Ratio between the MAE of the median of the adaptive ensemble and of the original ensemble across different percentages of trajectory selected and over all weeks. A ratio below 1 indicates better performance of the adaptive ensemble with respect to the original ensemble. Each boxplot is based on 160 data points, corresponding to the 16 weeks in which the adaptive ensemble was generated across 10 bootstrapping runs. The overlaid swarmplot points correspond to each week and the square markers indicate the mean value. The boxplot boundaries represent the interquartile range (IQR) between the first quartile (Q1) and third quartile (Q3), and the line inside each box indicates the median. The whiskers extend to the furthest data point within 1.5 times the IQR from Q1 and Q3.

#### S5.6 Sensitivity analysis for age-stratified applications

Here, we present a sensitivity analysis to explore whether the adaptive ensemble improvements hold when applied to 65+ age group. We use the 65+ age group for the 2024-2025 season as a case study because a sufficient number of models for constructing an ensemble was available (5 models) [21]. We could not test the pipeline on 2023-2024 season because only three modeling teams submitted age-stratified projections, which is not a sufficient number for generating an ensemble.

Specifically, we use target data for 65+ age group collected from the Flu Scenario Modeling Hub [22]. We start the pipeline from 2024-11-23 with only three surveillance data available for calibration because the target data were available only from 2024-11-09. Also, we note that, unlike for the overall population, a proper real-time exercise is not possible because non-backfilled data are not available through the weekly target data file. Despite these differences from the main analysis, the results (Fig. S9) show that the improvements of the adaptive ensemble with respect to the original ensemble hold at different tested percentages of selections and across both performance metrics. This demonstrates the potential of our approach for age-stratified analyses.

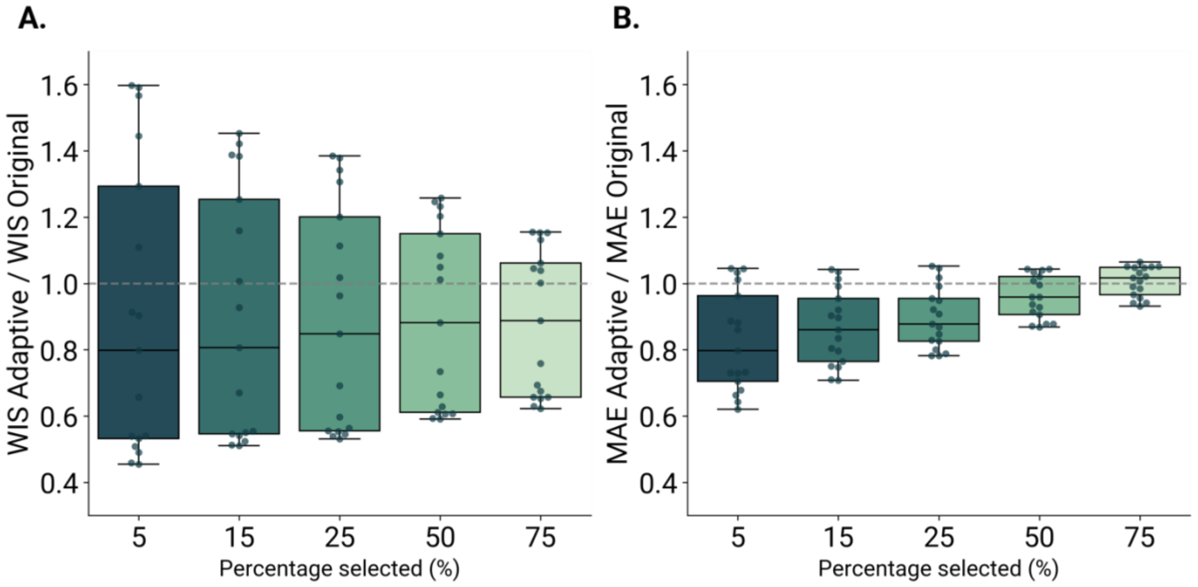


**Figure S9. Performance of the adaptive ensemble model on US-level projections from the U.S. Flu Scenario Modeling Hub Round 1, season 2024-2025.** The adaptive ensemble approach has been tested on 65+ age-stratified projections. A) Ratio between the WIS of the adaptive ensemble and of the original ensemble across different percentages of trajectory selected and over all weeks. B) Ratio between the MAE of the median of the adaptive ensemble and of the original ensemble across different percentages of trajectory selected and over all weeks. A ratio below 1 indicates better performance of the adaptive ensemble with respect to the original ensemble. Each boxplot is based on 17 data points, corresponding to the weeks in which the adaptive ensemble was generated. The boxplot boundaries represent the interquartile range (IQR) between the first quartile (Q1) and third quartile (Q3), and the line inside each box indicates the median. The whiskers extend to the furthest data point within 1.5 times the IQR from Q1 and Q3.

#### S5.7 Similarity of the sets of trajectories selected in different weeks

In Fig. S10 and Fig. S11, we show the Jaccard Similarity Index (see Sec. 4.2) of the set of trajectories selected in each week for the 2024-2025 and 2023-2024, respectively, measured in two ways: first, comparing the similarity between the pool of trajectories selected at time t and those selected at the initial time step $t_0$, JSI(t, $t_0$) (Fig. S10A and Fig. S11A); second, comparing the similarity between the pool at time t and at the previous time step t−1, JSI(t, t−1) (Fig. S10B and Fig. S11B). The evolution of JSI(t, $t_0$) highlights substantial differences with the initial set of selected trajectories, which stabilizes after 8 weeks (January 18, 2025) and 11 weeks (January 20, 2024). The trend of JSI(t, t−1) shows a gradual increase in similarity over time, suggesting that additional surveillance information contributes to stabilizing the set of trajectories in the adaptive ensemble. Notably, JSI(t, t−1) exceeds 80% within 3 weeks for the 25% case in 2024-2025 season, and 4 weeks in 2023-2024 season.

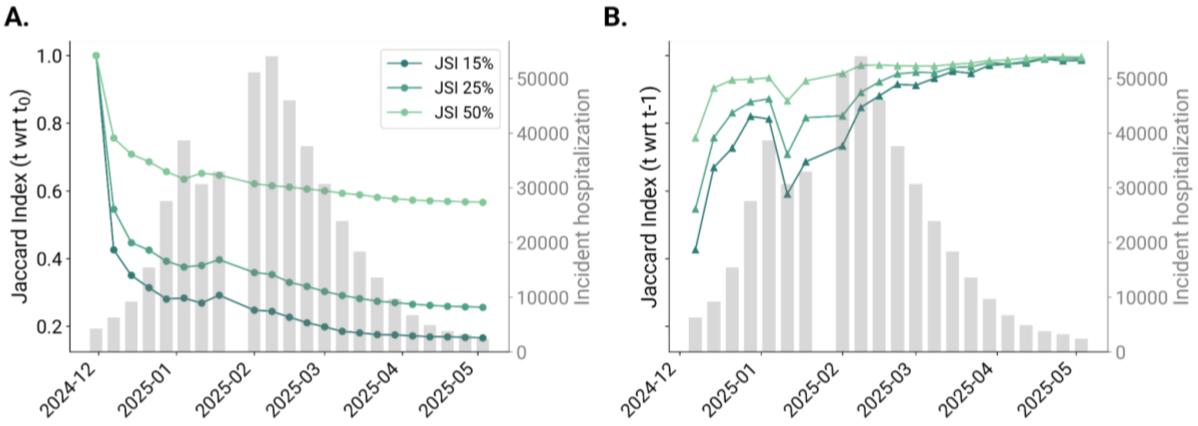


**Figure S10. Persistence analysis of the selected trajectories from the adaptive ensemble model on US-level projections from the U.S. Flu Scenario Modeling Hub Round 1, season 2024-2025.** A) Jaccard Similarity index of the set of trajectories selected each week with respect to the initial time step $t_0$ across selected percentages. B) Jaccard Similarity index of the set of trajectories selected each week with respect to the previous time step t-1 across selected percentages. In the background, the reported incident hospitalization [20].

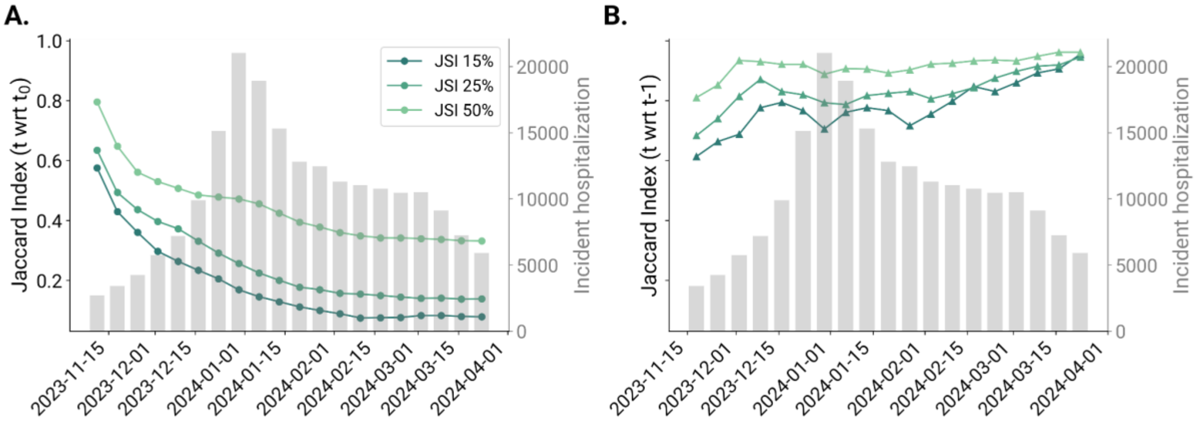


**Figure S11. Persistence analysis of the selected trajectories from the adaptive ensemble model on US-level projections from the U.S. Flu Scenario Modeling Hub Round 1, season 2023-2024.** A) Jaccard Similarity index of the set of trajectories selected each week with respect to the initial time step $t_0$ across selected percentages. B) Jaccard Similarity index of the set of trajectories selected each week with respect to the previous time step t-1 across selected percentages. In the background, the reported incident hospitalization [20].

#### S5.8 States and territories performance (additional analyses)

In Fig. S12 and Fig. S13, we present the performance analysis of projections for incident weekly influenza hospitalizations across U.S. states and territories in the 2024-2025 season, focusing on the 15% and 50% trajectory selection cases. The results are consistent with those reported for the 25% case. Specifically, for the 15% trajectory selection case, 48 out of 51 U.S. states and territories exhibit a median WIS ratio below 1, and 47 out of 51 show a median MAE ratio below 1. Similarly, for the 50% trajectory selection case, 47 out of 51 U.S. states and territories feature a median WIS ratio below 1, and 48 out of 51 show a median MAE ratio below 1.

Figs. S14-S16 present the results of the same analysis for the 2023-2024 season, using 25%, 15%, and 50% of selected trajectories, respectively. Consistent with the previous case study, the adaptive ensemble outperforms the original ensemble in over 70% of U.S. states and territories across all cases, based on both WIS and MAE metrics.

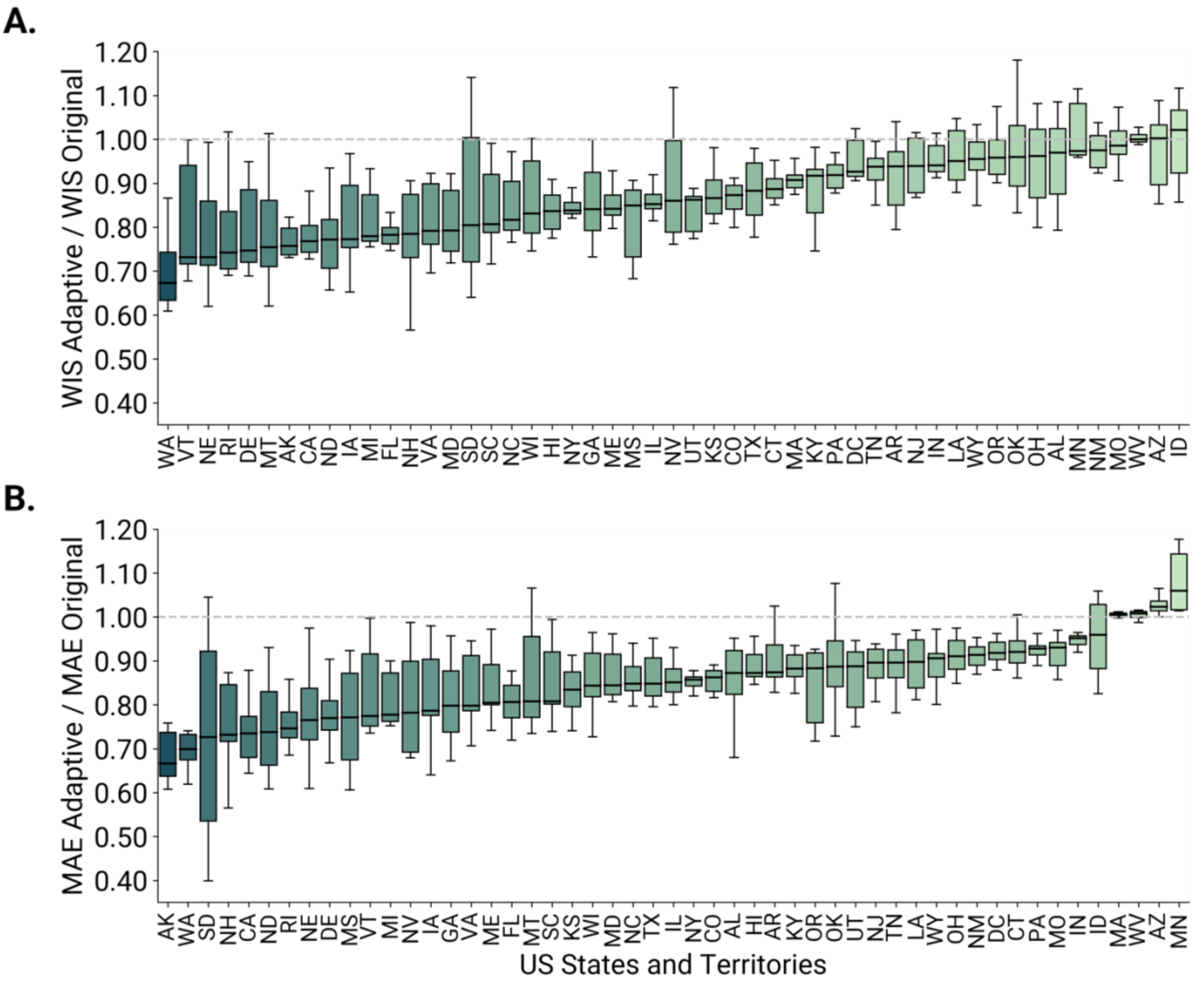


**Figure S12. Performance of adaptive ensemble (15% selected trajectories) on state-level projections (US Flu Scenario Modeling Hub Round 1, season 2024-2025).** A) For each U.S. state and territory, we show the ratio between the WIS of the adaptive ensemble and of the original ensemble for the **15%** percentage of trajectory selected and over all weeks. B) Ratio between the MAE of the median of the adaptive ensemble and of the original ensemble for the **15%** percentage of trajectory selected and over all weeks. A ratio below 1 indicates better performance of the adaptive ensemble with respect to the original ensemble. Each boxplot is based on 16 data points, corresponding to the weeks in which the adaptive ensemble was generated. The boxplot boundaries represent the interquartile range (IQR) between the first quartile (Q1) and third quartile (Q3), and the line inside each box indicates the median. The whiskers extend to the furthest data point within 1.5 times the IQR from Q1 and Q3.

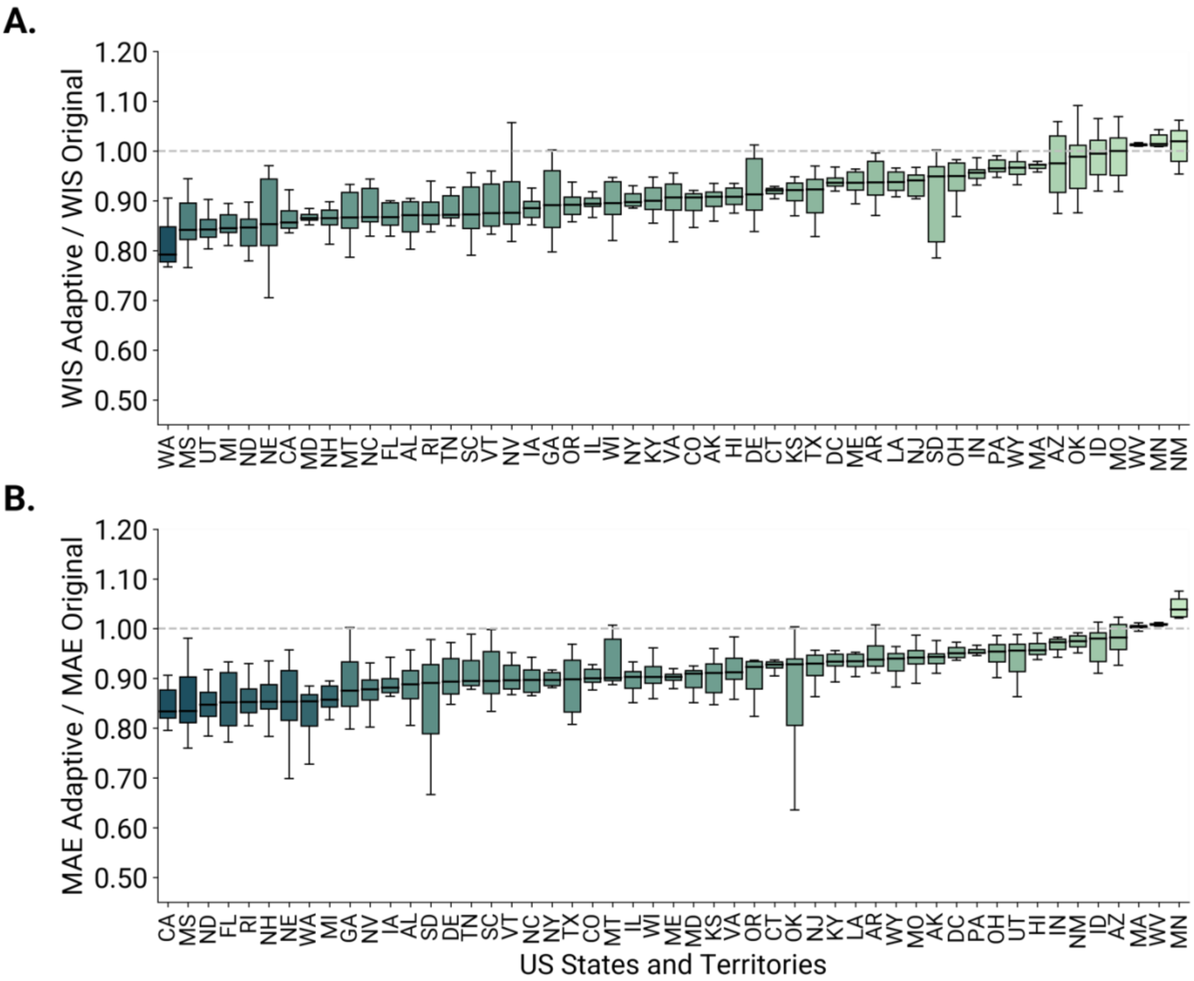


**Figure S13. Performance of adaptive ensemble (50% selected trajectories) on state-level projections (US Flu Scenario Modeling Hub Round 1, season 2024-2025).** A) For each U.S. state and territory, we show the ratio between the WIS of the adaptive ensemble and of the original ensemble for the **50%** percentage of trajectory selected and over all weeks. B) Ratio between the MAE of the median of the adaptive ensemble and of the original ensemble for the **50%** percentage of trajectory selected and over all weeks. A ratio below 1 indicates better performance of the adaptive ensemble with respect to the original ensemble. Each boxplot is based on 16 data points, corresponding to the weeks in which the adaptive ensemble was generated. The boxplot boundaries represent the interquartile range (IQR) between the first quartile (Q1) and third quartile (Q3), and the line inside each box indicates the median. The whiskers extend to the furthest data point within 1.5 times the IQR from Q1 and Q3.

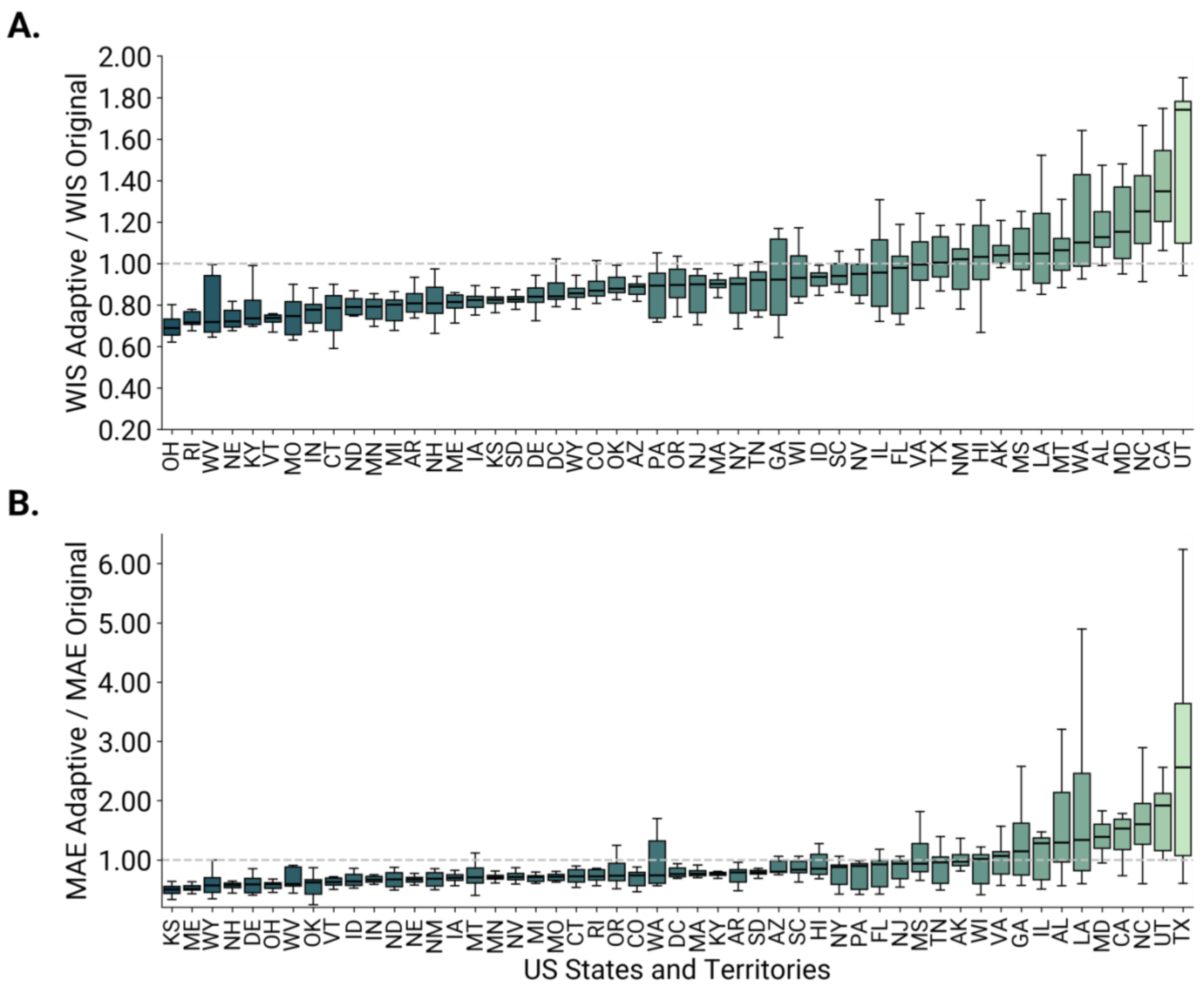


**Figure S14. Performance of adaptive ensemble (25% selected trajectories) on state-level projections (US Flu Scenario Modeling Hub Round 1, season 2023-2024).** A) For each U.S. state and territory, we show the ratio between the WIS of the adaptive ensemble and of the original ensemble for the **25%** percentage of trajectory selected and over all weeks. B) Ratio between the MAE of the median of the adaptive ensemble and of the original ensemble for the **50%** percentage of trajectory selected and over all weeks. A ratio below 1 indicates better performance of the adaptive ensemble with respect to the original ensemble. Each boxplot is based on 20 data points, corresponding to the weeks in which the adaptive ensemble was generated. The boxplot boundaries represent the interquartile range (IQR) between the first quartile (Q1) and third quartile (Q3), and the line inside each box indicates the median. The whiskers extend to the furthest data point within 1.5 times the IQR from Q1 and Q3.

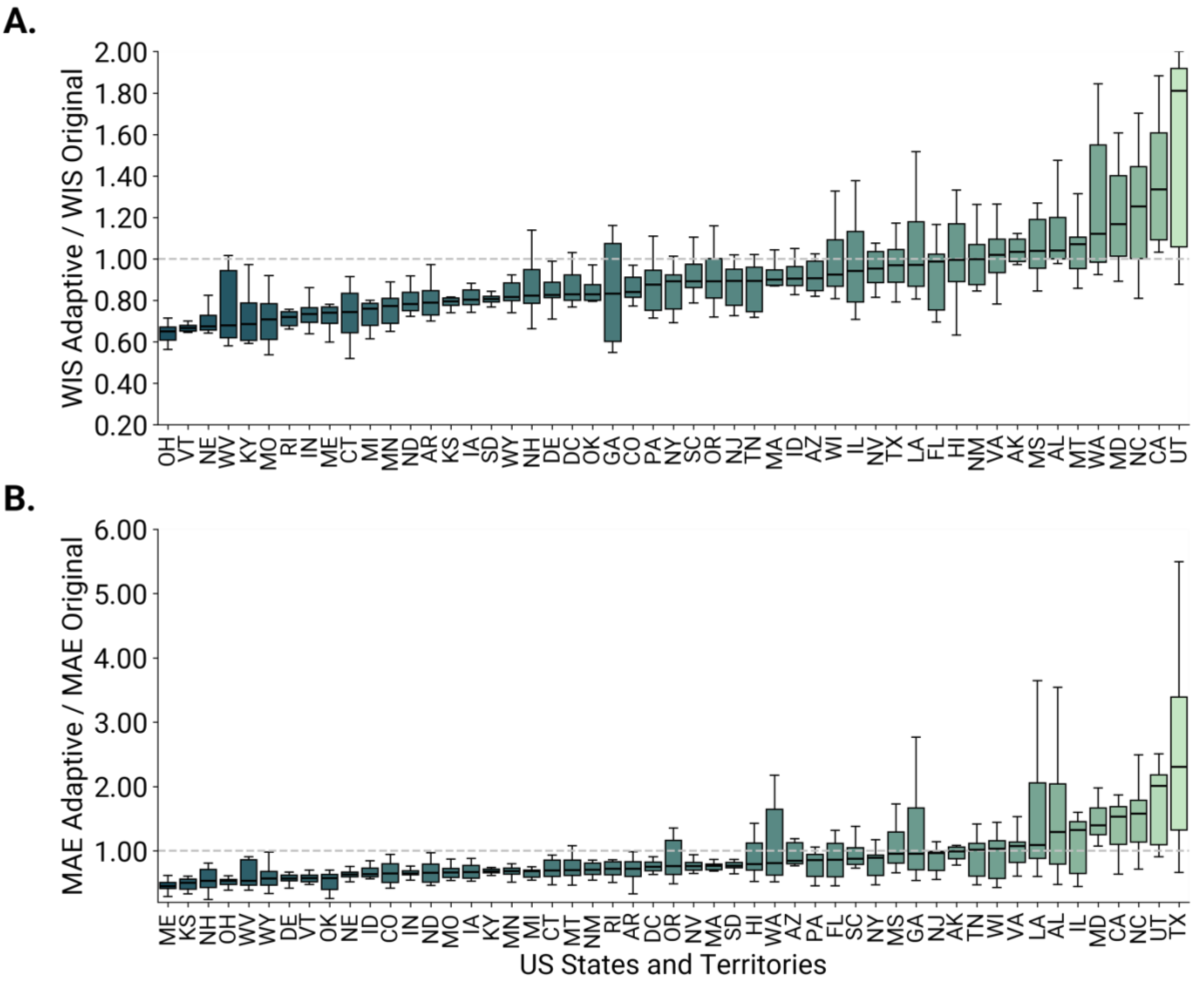


**Figure S15. Performance of adaptive ensemble (15% selected trajectories) on state-level projections (US Flu Scenario Modeling Hub Round 1, season 2023-2024).** A) For each U.S. state and territory, we show the ratio between the WIS of the adaptive ensemble and of the original ensemble for the **15%** percentage of trajectory selected and over all weeks. B) Ratio between the MAE of the median of the adaptive ensemble and of the original ensemble for the **50%** percentage of trajectory selected and over all weeks. A ratio below 1 indicates better performance of the adaptive ensemble with respect to the original ensemble. Each boxplot is based on 20 data points, corresponding to the weeks in which the adaptive ensemble was generated. The boxplot boundaries represent the interquartile range (IQR) between the first quartile (Q1) and third quartile (Q3), and the line inside each box indicates the median. The whiskers extend to the furthest data point within 1.5 times the IQR from Q1 and Q3.

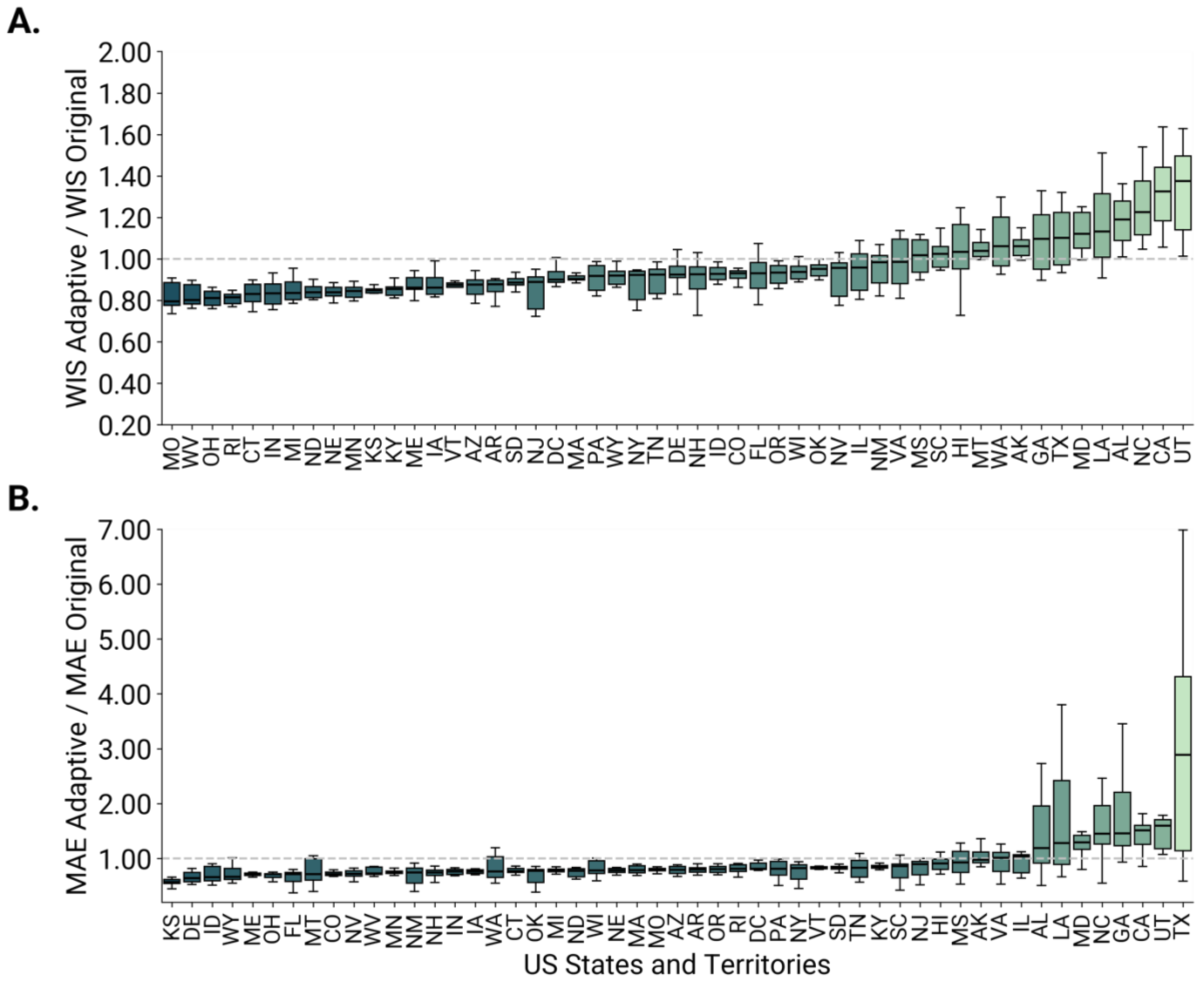


**Figure S16. Performance of adaptive ensemble (50% selected trajectories) on state-level projections (US Flu Scenario Modeling Hub Round 1, season 2023-2024).** A) For each U.S. state and territory, we show the ratio between the WIS of the adaptive ensemble and of the original ensemble for the **50%** percentage of trajectory selected and over all weeks. B) Ratio between the MAE of the median of the adaptive ensemble and of the original ensemble for the **50%** percentage of trajectory selected and over all weeks. A ratio below 1 indicates better performance of the adaptive ensemble with respect to the original ensemble. Each boxplot is based on 20 data points, corresponding to the weeks in which the adaptive ensemble was generated. The boxplot boundaries represent the interquartile range (IQR) between the first quartile (Q1) and third quartile (Q3), and the line inside each box indicates the median. The whiskers extend to the furthest data point within 1.5 times the IQR from Q1 and Q3.

#### S5.9 Exploring posterior probability of alternative epidemic scenarios (additional analyses)

Fig. S17 compares the observed cumulative subtype proportions with the posterior scenario contributions over time. The top panel shows the cumulative proportions of A/H3N2 and A/H1N1, computed from surveillance data available through the FluView Interactive dashboard [23]. The bottom panel shows the posterior probability of each scenario, where red shades correspond to scenarios assuming A/H3N2 dominance and blue shades to scenarios assuming A/H1N1 dominance. At the beginning of the season, a slightly higher proportion of A/H3N2 was observed, with the adaptive ensemble assigning higher posterior probability to A/H1N1-dominant scenarios, while still retaining a non-negligible contribution from A/H3N2-dominant ones. From week 5 of 2025 onwards, the two subtypes co-circulated with comparable cumulative positivity, a pattern that is progressively reflected in the posterior probabilities as more data become available.

In Fig. S18, we present the posterior probability of alternative scenarios over time for the 15% and 50% trajectory selection cases in the 2024-2025 season, showing the fraction of trajectories originating from each scenario that are selected weekly by the adaptive ensemble for U.S. national-level projections. The results are consistent with those observed for the 25% case in the main text. Scenario B emerges as the most likely scenario at the beginning of the influenza season in the 15% case, but it shares similar probabilities with Scenario F and Scenario D in the 50% case. Starting from week 3, 2025, Scenario E (15%) and Scenario F (50%) take place as leading scenarios for the entire period.

Figs. S19-S21 report the same analysis for the 2023-2024 season. Both the observed cumulative subtype proportions and the posterior scenario contributions consistently indicate A/H1N1 dominance throughout the season (Fig. S19). At the beginning of the winter season (October 2023), all scenarios are nearly equally likely across both percentages. However, as the season progresses, scenarios B, D, and F, associated with a season dominated by influenza subtype A/H1N1 and varying assumptions on vaccination coverage (higher-than-usual, business-as-usual, and lower-than-usual, respectively), emerge as more likely compared to scenarios dominated by influenza subtype A/H3N2. By week 52 of 2023, coinciding with the peak in incident hospitalizations, scenario F becomes the most likely for the 15% case (fraction of selected trajectories $(f_F(25\%) = 0.24,\ f_F(15\%) = 0.25)$, similar to the 25% case presented in the manuscript. For the 50% case, scenarios F and D share the highest fraction of selected trajectories $(f_F(50\%) = f_D(50\%) = 0.21)$. In all the cases, scenario B follows scenarios F and D, whereas scenarios associated with A/H3N2 dominance (A, C, E) are substantially less likely, with fractions ranging between 0.10 and 0.13. After the peak, scenarios B and D increasingly dominate, with scenario B becoming the leading scenario from week 7 of 2024 onward. However, the dominance is not so clear, with scenario D showing slightly lower probabilities compared to Scenario B.

Figure S22 shows the results of the posterior scenario analysis applied on the age group 65+, for 2024-2025 season, using the age-stratified projections. At the beginning of the season (week 2024-48), Scenario D, assuming A/H1N1 subtype dominance and business-as-usual vaccination coverage, was the most selected scenario, followed closely by Scenario E, assuming A/H3N2 dominance and lower-than-usual vaccination coverage. From week 2024-49 onwards, Scenario E was consistently the most selected scenario throughout the rest of the season, with Scenario C (A/H3N2 dominance and business-as-usual vaccination coverage) ranking second, and non-negligible posterior weight retained by Scenario D (A/H1N1 dominance and business-as-usual vaccination coverage). These results are broadly consistent with observed vaccine coverage data reported by the Centers for Disease Control and Prevention (CDC) for the 65+ age group [24]: in 2024-2025, vaccine coverage reached 49.1% in this age class, compared to 54.5% in 2022-2023 (used as the reference season), indicating a slightly lower-than-usual uptake, sitting between the lower-than-usual and business-as-usual assumptions, coherent with the dominant selection of Scenario E, followed by Scenarios C and D. Regarding subtype dominance, A/H3N2 and A/H1N1 co-circulated among individuals aged 65 and older throughout the season, with A/H3N2 slightly predominant at the beginning and A/H1N1 gaining relative dominance from week 2025-02 onwards [25, 26].

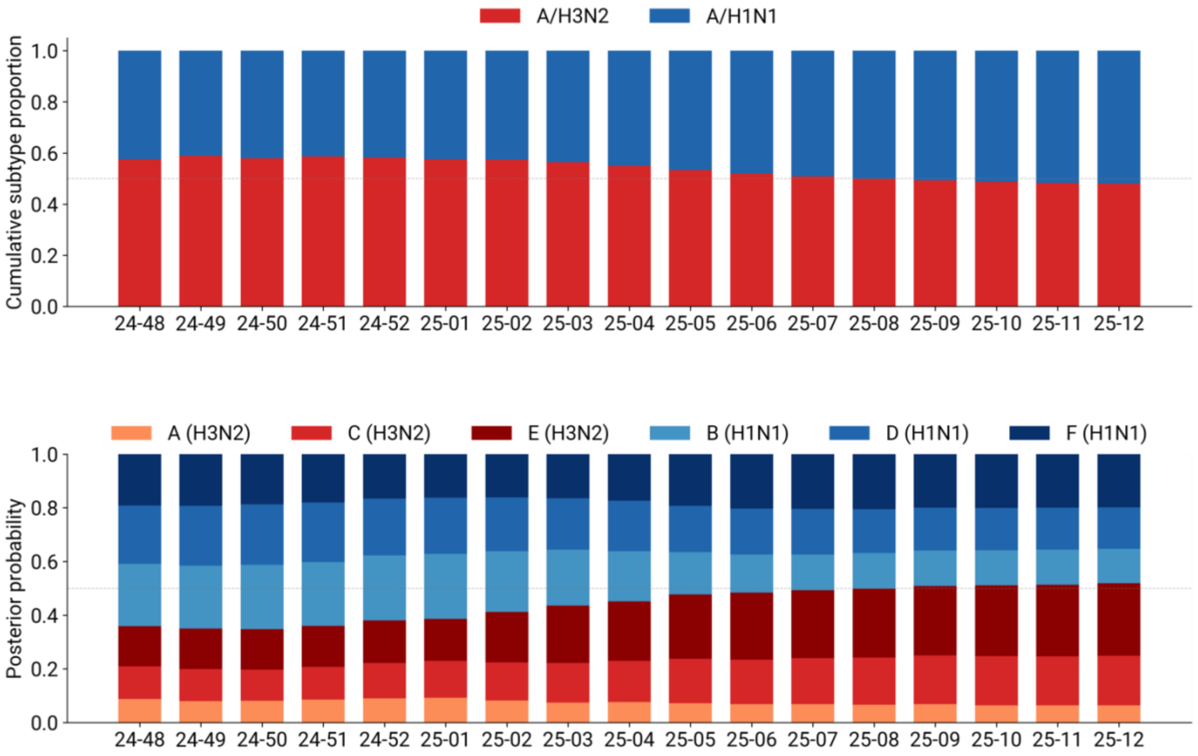


**Figure S17. Comparing observed and estimated influenza subtypes for 2024/2025 US influenza season (US Flu Scenario Modeling Hub Round 1, season 2024-2025).** Top panel: cumulative proportion of A/H3N2 and A/H1N1 subtypes over the weeks, computed from surveillance data available through the FluView Interactive dashboard [20]. Bottom panel: posterior probability of each scenario at each epidemiological week, where red shades correspond to scenarios assuming A/H3N2 dominance (Scenarios A, C, E) and blue shades to scenarios assuming A/H1N1 dominance (Scenarios B, D, F), using the selection threshold of k=25%.

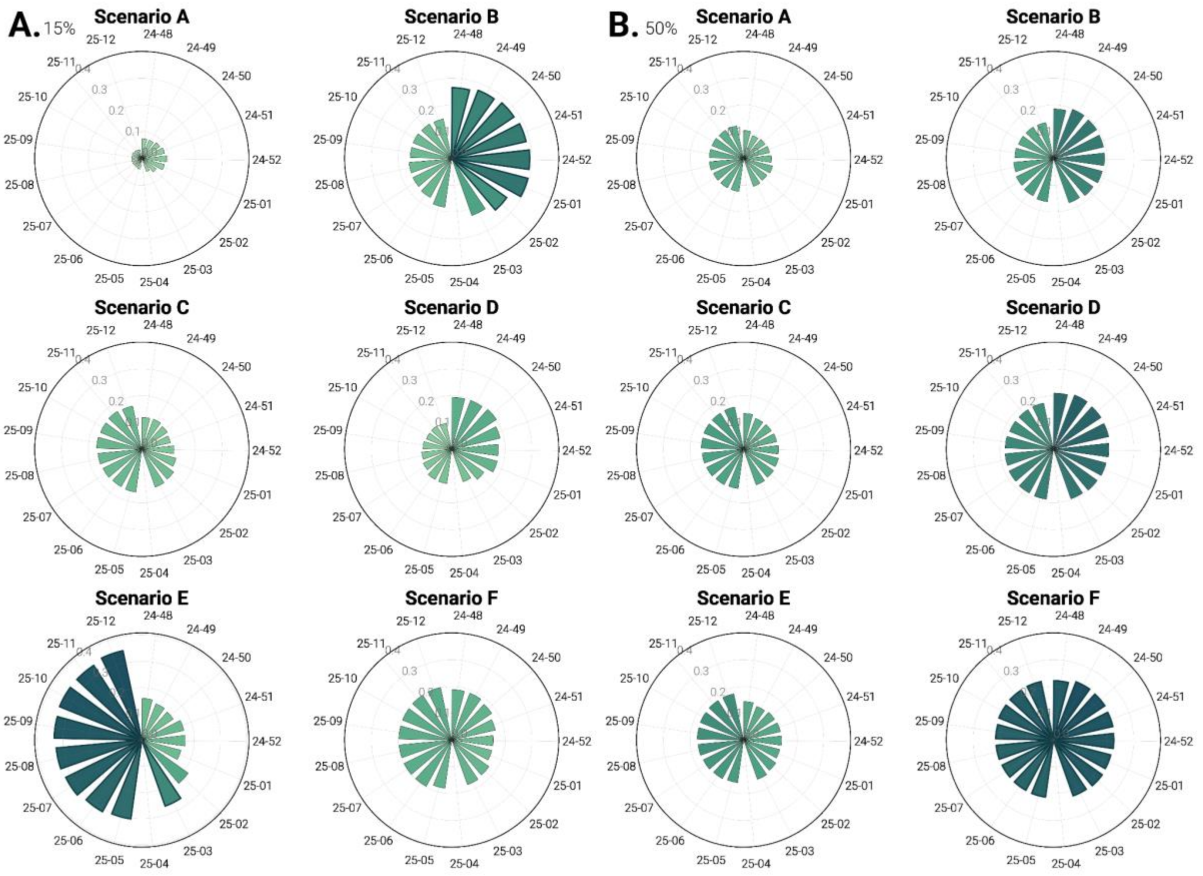


**Figure S18. Exploring scenario plausibility for 2024-2025 U.S. influenza season (U.S. Flu Scenario Modeling Hub Round 1, season 2024-2025).** The plot shows the weekly fraction of trajectories originating from each scenario by the adaptive ensemble: (A) top 15% and (B) top 50%. Scenarios assuming A/H1N1 dominance in the 2023-2024 season are shown on the right side (B, D, F), and those assuming A/H3N2 dominance are on the left side (A, C, E). Scenarios in the first row assume 20% higher vaccination coverage in all age groups and jurisdictions with respect to 2021-2022 coverage, scenarios in the second row assume business-as-usual coverage, and scenarios in the third row assume a 20% lower coverage.

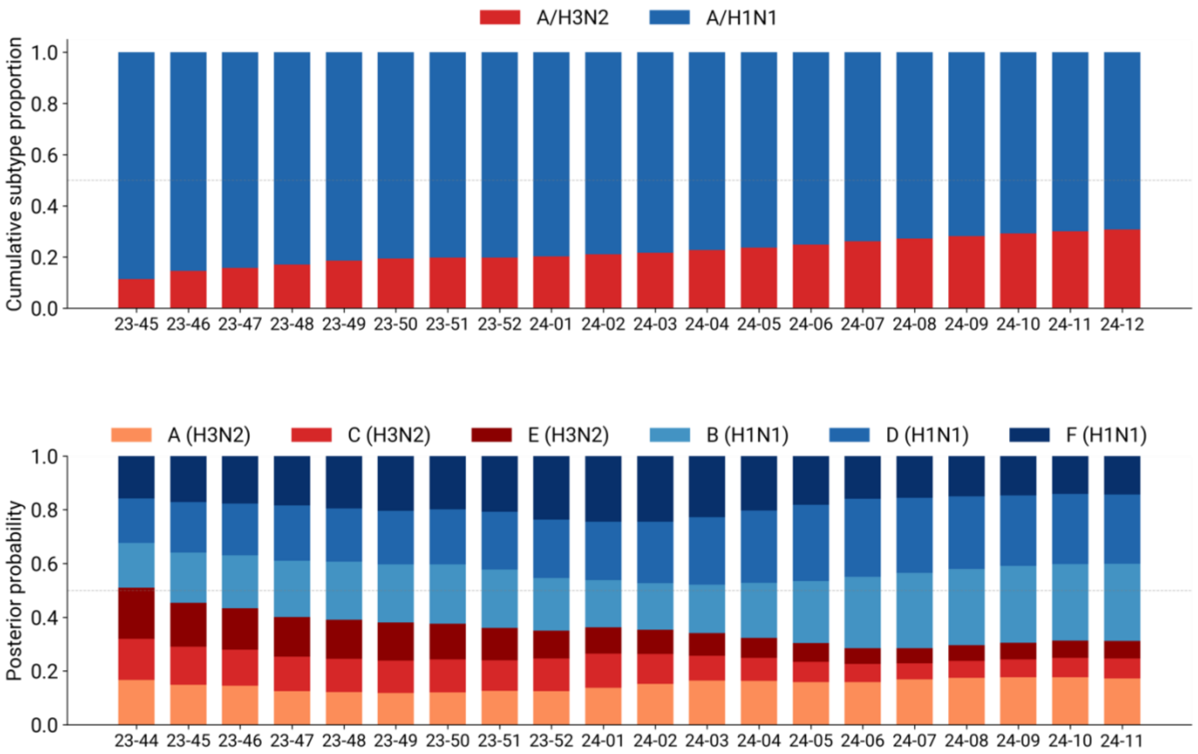


**Figure S19. Comparing observed and estimated influenza subtypes for 2023-2024 U.S. influenza season (U.S. Flu Scenario Modeling Hub Round 1, season 2023-2024).** Top panel: cumulative proportion of A/H3N2 and A/H1N1 subtypes over the weeks, computed from surveillance data available through the FluView Interactive dashboard [20]. Bottom panel: posterior probability of each scenario at each epidemiological week, where red shades correspond to scenarios assuming A/H3N2 dominance (Scenarios A, C, E) and blue shades to scenarios assuming A/H1N1 dominance (Scenarios B, D, F), using the selection threshold of k=25%.

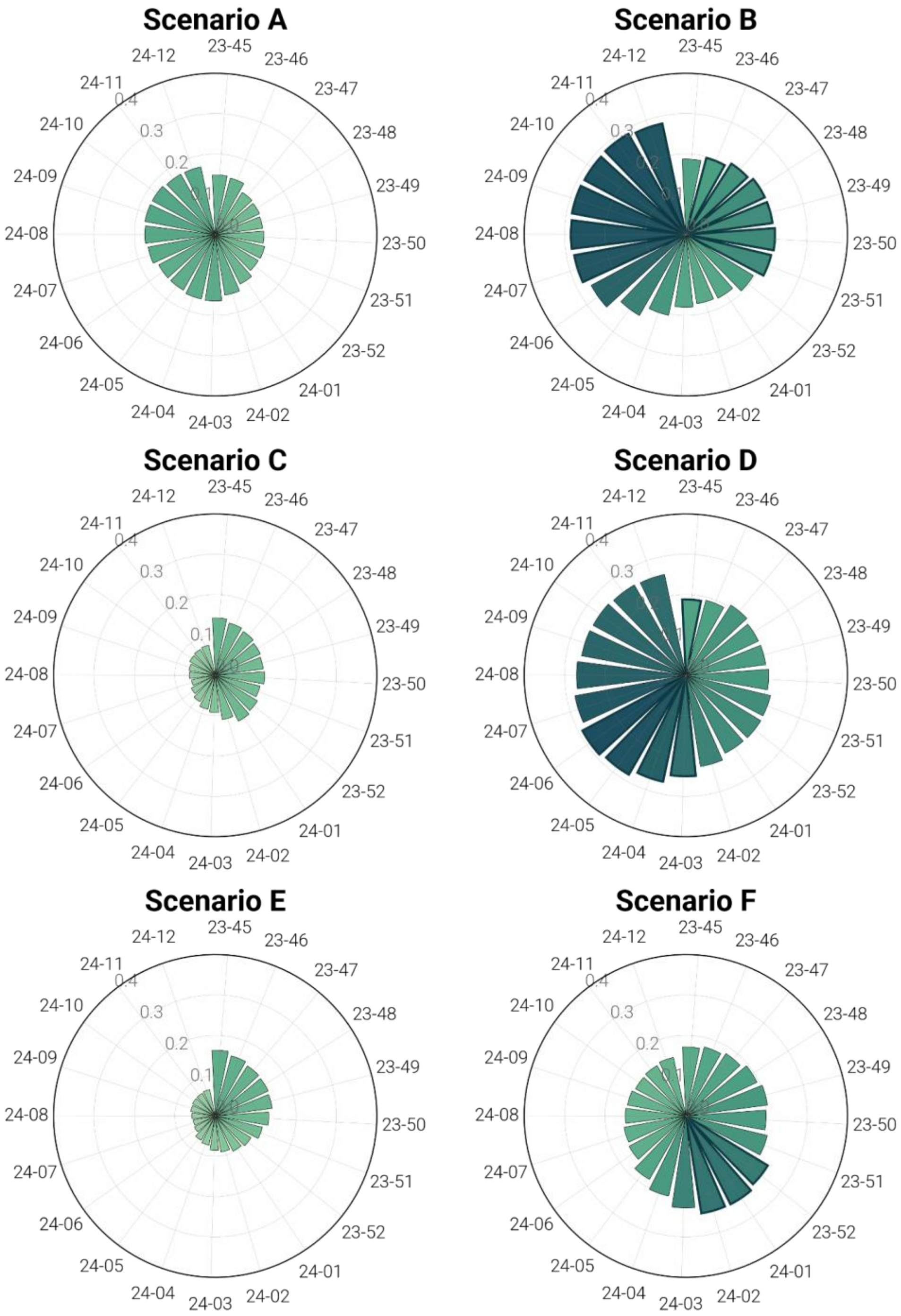


**Figure S20. Exploring scenario plausibility for 2023-2024 U.S. influenza season (U.S. Flu Scenario Modeling Hub Round 1, season 2023-2024).** The plot shows the weekly fraction of trajectories originating from each scenario by the adaptive ensemble considering the top 25% case. Scenarios assuming A/H1N1 dominance in the 2023-2024 season are shown on the right side (B, D, F), and those assuming A/H3N2 dominance are on the left side (A, C, E). Scenarios in the first row assume 20% higher vaccination coverage in all age groups and jurisdictions with respect to 2021-2022 coverage, scenarios in the second row assume a business as usual coverage, and scenarios in the third row assume a 20% lower coverage.

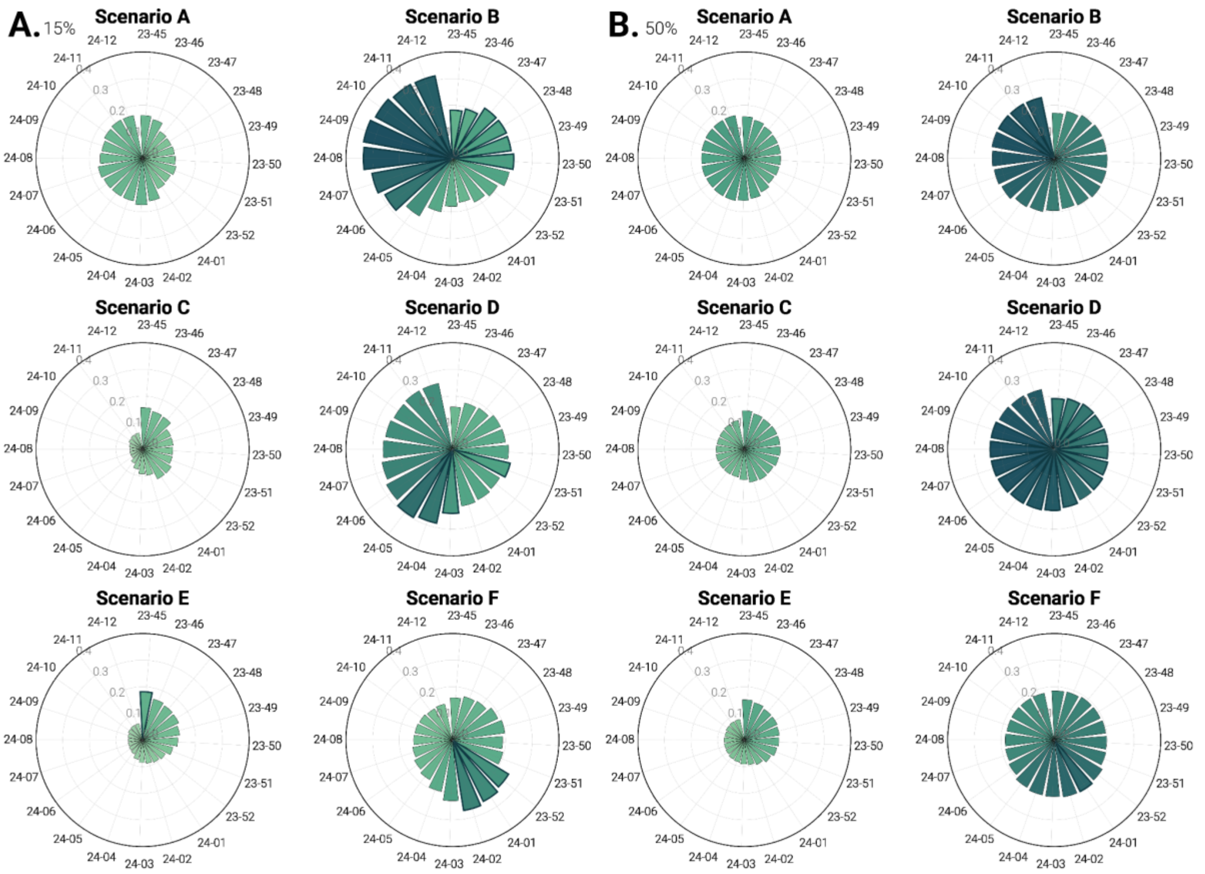


**Figure S21. Exploring scenario plausibility for 2023-2024 U.S. influenza season (U.S. Flu Scenario Modeling Hub Round 1, season 2023-2024).** The plot shows the weekly fraction of trajectories originating from each scenario by the adaptive ensemble: (A) top 15% and (B) top 50%. Scenarios assuming A/H1N1 dominance in the 2023-2024 season are shown on the right side (B, D, F), and those assuming A/H3N2 dominance are on the left side (A, C, E). Scenarios in the first row assume 20% higher vaccination coverage in all age groups and jurisdictions with respect to 2021-2022 coverage, scenarios in the second row assume a business as usual coverage, and scenarios in the third row assume a 20% lower coverage.

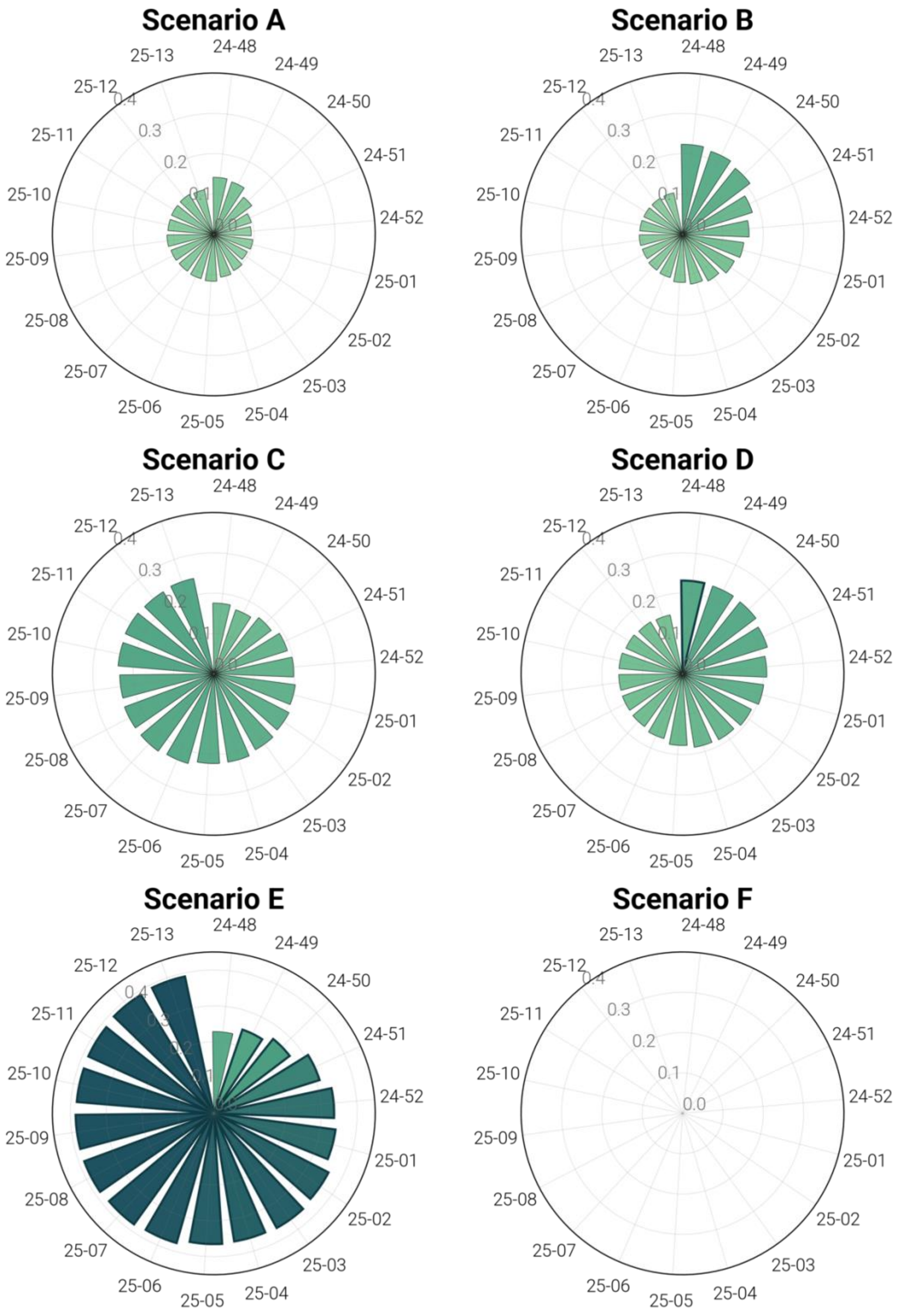


**Figure S22. Exploring scenario plausibility for 2024-2025 U.S. influenza season, age-stratification, 65+ age group (U.S. Flu Scenario Modeling Hub Round 1, season 2024-2025).** The plot shows the weekly fraction of trajectories originating from each scenario by the adaptive ensemble considering the top 25% case. Scenarios assuming A/H1N1 dominance in the 2023-2024 season are shown on the right side (B, D, F), and those assuming A/H3N2 dominance are on the left side (A, C, E). Scenarios in the first row assume 20% higher vaccination coverage in all age groups and jurisdictions with respect to 2021-2022 coverage, scenarios in the second row assume a business as usual coverage, and scenarios in the third row assume a 20% lower coverage.

### S6. Evaluating adaptive ensemble for short-term forecasting

#### S6.1 Additional analyses for 2024-2025 season

##### S6.1.1 Forecasting performance by horizon, 2024-2025

Figs. S23-S24 shows boxplots summarizing, for different percentages of selected trajectories, the distribution of the ratios between the WIS and MAE of the median of the adaptive ensemble and those of the FluSight baseline and ensemble models, separately for each forecasting horizon. In the following analysis, results are reported by forecast horizon, where horizon 0 represents the 1-week-ahead forecast, i.e., the forecast for the current week based on surveillance data available up to the previous week. Consequently, horizon 1 corresponds to the 2-week-ahead forecast, horizon 2 to the 3-week-ahead forecast, and horizon 3 to the 4-week-ahead forecast. The results are consistent with what we observed and described in the main text, where results for different horizons were averaged. Interestingly, in general, the adaptive ensemble achieves better performance in median terms at higher horizons, both compared to the FluSight baseline and FluSight ensemble models.

In Figs. S25-S26, we present the one- to four-week-ahead forecasts, showing the median, 50%, and 90% predictive intervals for the top 25%, 15%, and 50% of selected trajectories, respectively. As the percentage of selected trajectories increases, we observe a widening of the prediction intervals. These results suggest that selecting a higher percentage of trajectories may lead to reduced sharpness in forecasts.

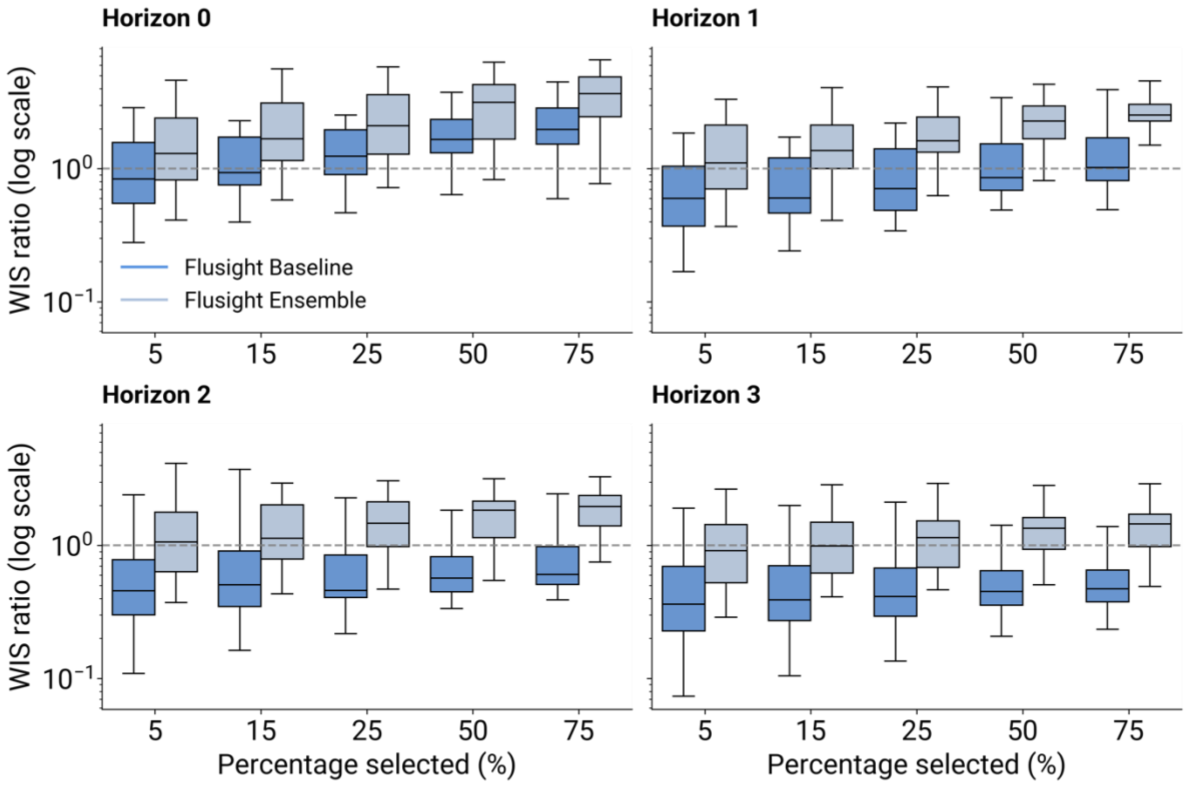


**Figure S23. Performance of the adaptive ensemble model in short-term forecasting (season 2024-2025).** Ratio (log-scale) between the WIS of the adaptive ensemble and the FluSight Baseline or FluSight ensemble models, across percentages of trajectory selection and for the four horizons separately. A ratio below 1 indicates that the adaptive ensemble outperformed the FluSight reference model. The boxplot boundaries represent the interquartile range (IQR) between the first quartile (Q1) and third quartile (Q3), and the line inside each box indicates the median. The whiskers extend to the furthest data point within 1.5 times the IQR from Q1 and Q3. Data points outside 1.5 times the interquartile range, above the upper quartile and below the lower quartile (Q1– 1.5 * IQR or Q3 + 1.5 * IQR), are considered outliers and not displayed.

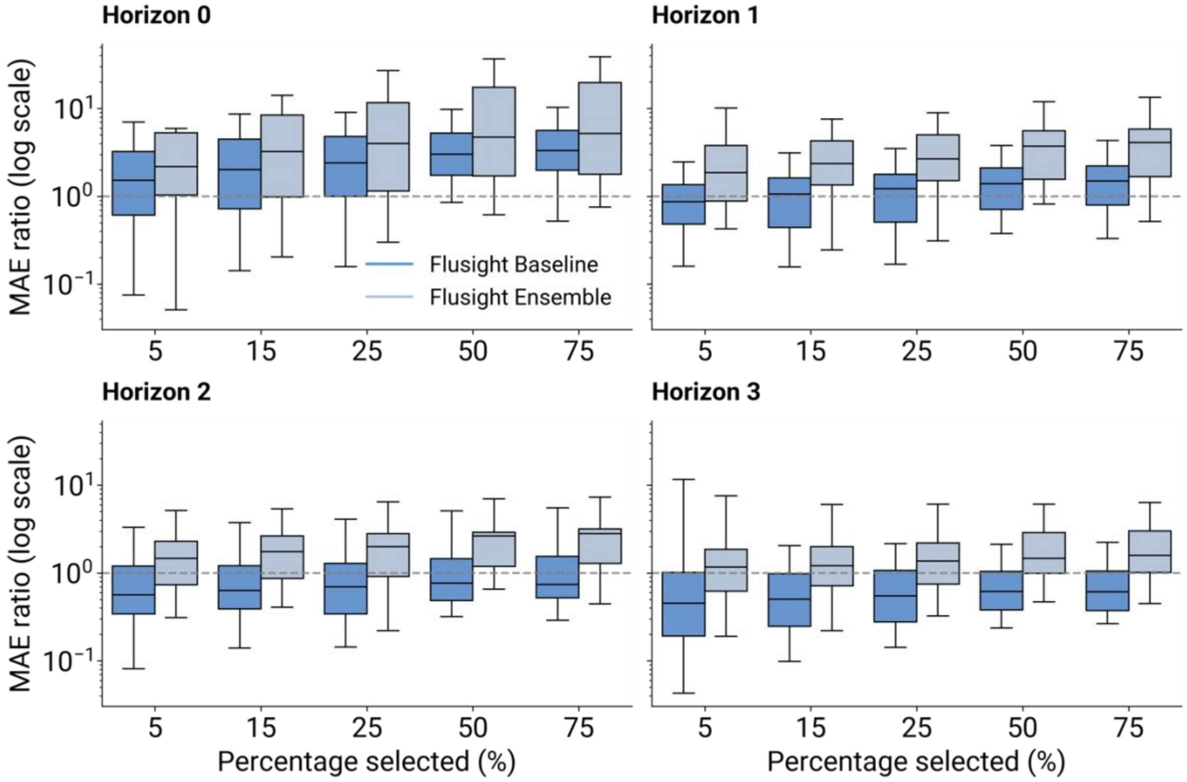


**Figure S24. Performance of the adaptive ensemble model in short-term forecasting (season 2024-2025).** Ratio (log-scale) between the MAE of the adaptive ensemble and the FluSight Baseline or FluSight ensemble models, across percentages of trajectory selection and for the four horizons separately. A ratio below 1 indicates that the adaptive ensemble outperformed the FluSight reference model. The boxplot boundaries represent the interquartile range (IQR) between the first quartile (Q1) and third quartile (Q3), and the line inside each box indicates the median. The whiskers extend to the furthest data point within 1.5 times the IQR from Q1 and Q3. Data points outside 1.5 times the interquartile range, above the upper quartile and below the lower quartile (Q1– 1.5 * IQR or Q3 + 1.5 * IQR), are considered outliers and not displayed.

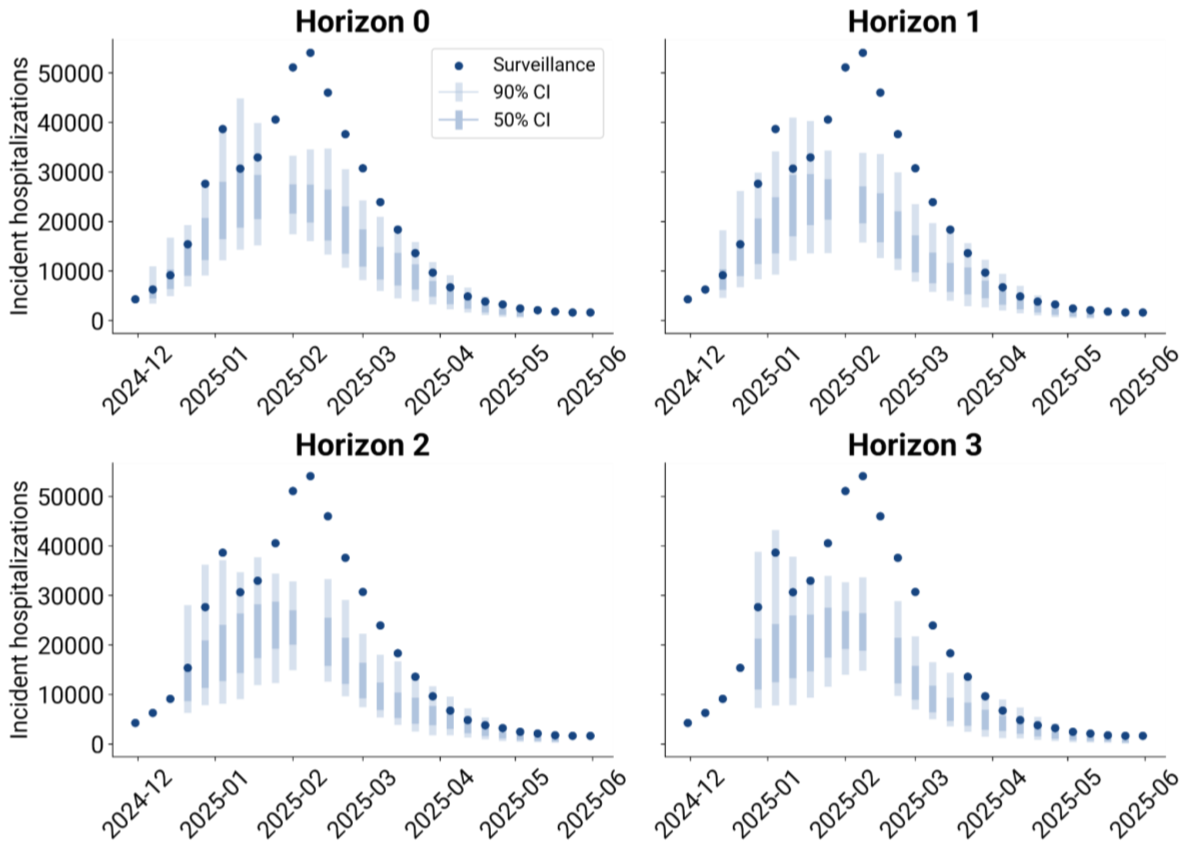


**Figure S25. Adaptive ensemble one- to four-week-ahead forecast (25% top trajectories, season 2024-2025).** The plots show one- to four-week-ahead forecasts, presenting the median, 50%, and 90% prediction intervals. These results are based on the adaptive ensemble using the top 25% of trajectories selected.

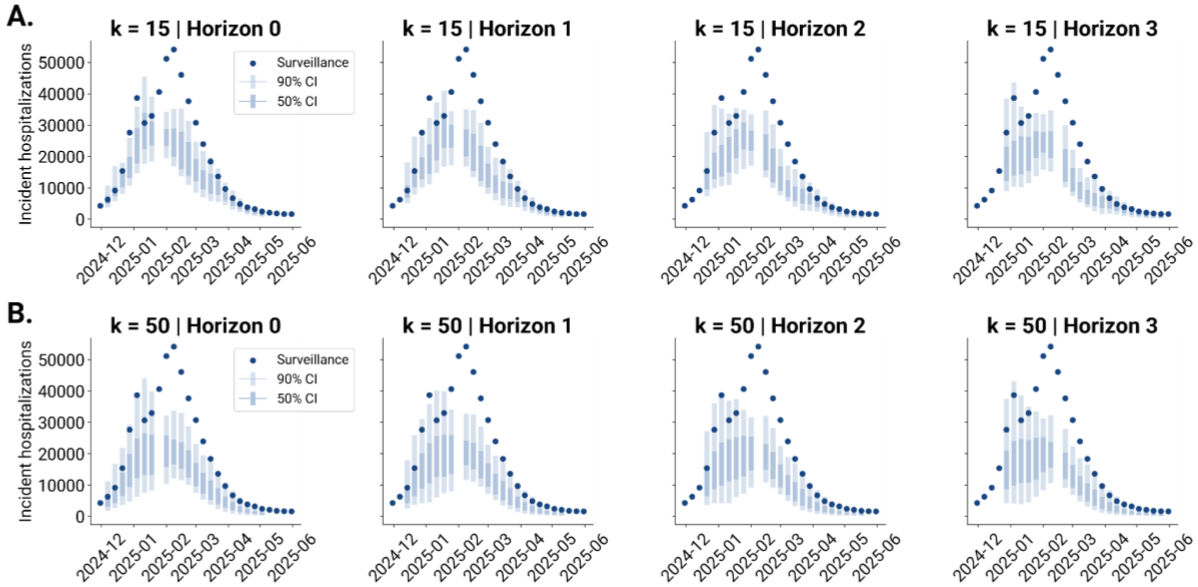


**Figure S26. Adaptive ensemble one- to four-week-ahead forecast (season 2024-2025).** The plot shows the one- to four-week-ahead forecasts, presenting the median, 50%, and 90% prediction intervals considering the 15% top trajectories for the adaptive ensemble generation (A) and the top 50% trajectories (B).

##### S6.1.2 Prediction coverage - 2024-2025

In Fig. S27, we compare the prediction coverage of the adaptive ensemble to that of the FluSight baseline and FluSight ensemble models, focusing on the 25% case for trajectory selection and 2024-2025 season. The four panels represent the coverage results for the four horizons individually, aggregated across all prediction time points. The adaptive ensemble demonstrates overconfidence across all horizons, with coverage values falling below the nominal levels. This indicates that its prediction intervals are narrower than those of a perfectly calibrated model. Similarly, the FluSight baseline model exhibits overconfidence for all horizons, with coverage values falling below the nominal levels. The FluSight ensemble performs the best in terms of coverage, showing values across different alpha levels that are closely aligned with the dashed diagonal, indicative of an ideally calibrated model. This finding is supported by results presented in Table S4, showing the coverage mismatch for the 0.5 and 0.9 nominal coverage levels (i.e. the absolute difference between the nominal values and the predicted coverage). In Fig. S28 we show the equivalent coverage plots for the 15% and 50% trajectory selection cases. Notably, the 15% case shows improved coverage results, particularly for longer time horizons, whereas for the 50% case we observe an increasing overconfidence across all horizons.

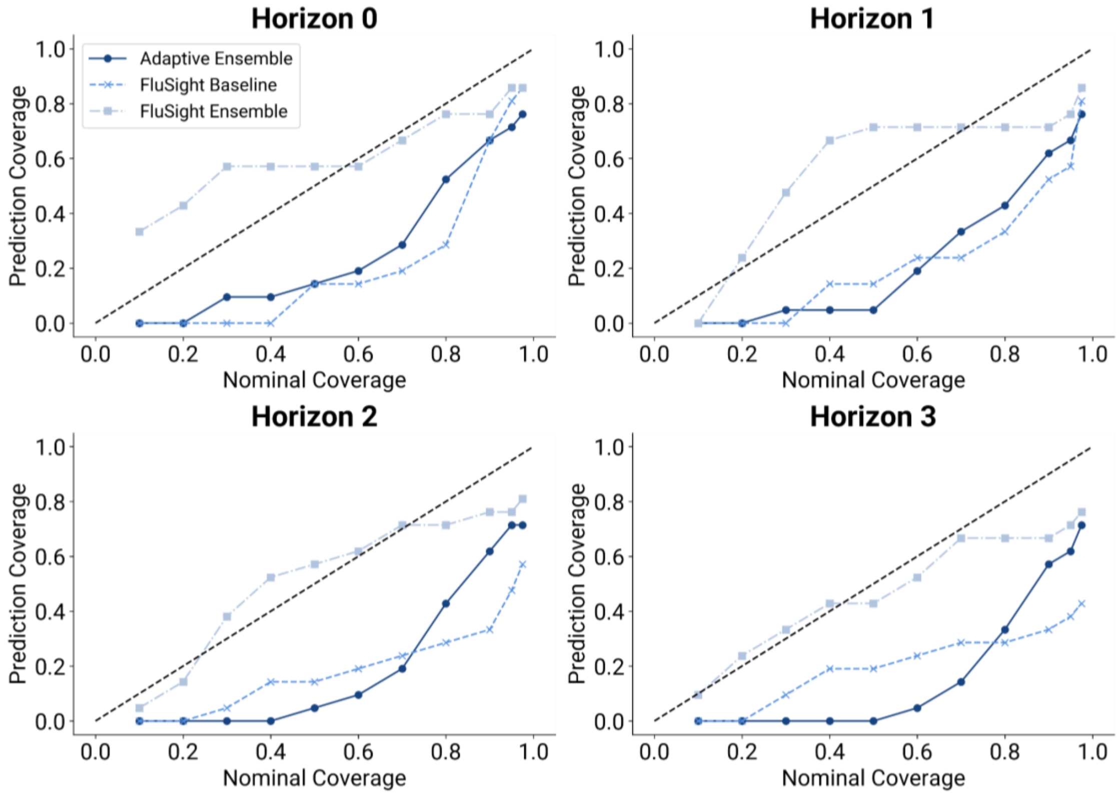


**Figure S27. Performance of the adaptive ensemble model in short-term forecasting (season 2024-2025).** The figure shows the coverage across different time horizons for the adaptive ensemble model, the FluSight baseline model, and the FluSight ensemble model. The adaptive ensemble has been generated using the 25% top trajectories. Each data point represents the coverage value for a specific alpha level (0.10, 0.20, 0.30, 0.40, 0.50, 0.60, 0.70, 0.80, 0.90, 0.95, 0.975). The black dashed diagonal serves as a reference, indicating the coverage of an ideally calibrated model.

| Metric | Horizon | Adaptive Ensemble | FluSight Baseline | FluSight Ensemble |
|---|---|---|---|---|
| *Coverage mismatch alpha = 0.5* | 0 | 0.31 | 0.20 | **0.17** |
| | 1 | 0.38 | 0.27 | **0.19** |
| | 2 | 0.38 | 0.3 | **0.10** |
| | 3 | 0.42 | 0.29 | **0.08** |
| *Coverage mismatch alpha = 0.9* | 0 | 0.23 | 0.15 | **0.06** |
| | 1 | 0.28 | 0.28 | **0.13** |
| | 2 | 0.30 | 0.46 | **0.10** |
| | 3 | 0.32 | 0.53 | **0.23** |

**Table S4: Coverage mismatch summary of adaptive ensemble in short-term forecasting (season 2024-2025).** The table presents the absolute differences between nominal and predicted coverage values (coverage mismatch) for alpha levels 0.5 and 0.9, across four time horizons. Results are reported for the adaptive ensemble, FluSight baseline, and FluSight ensemble models. The best-performing values for each alpha level and horizon are highlighted in bold.

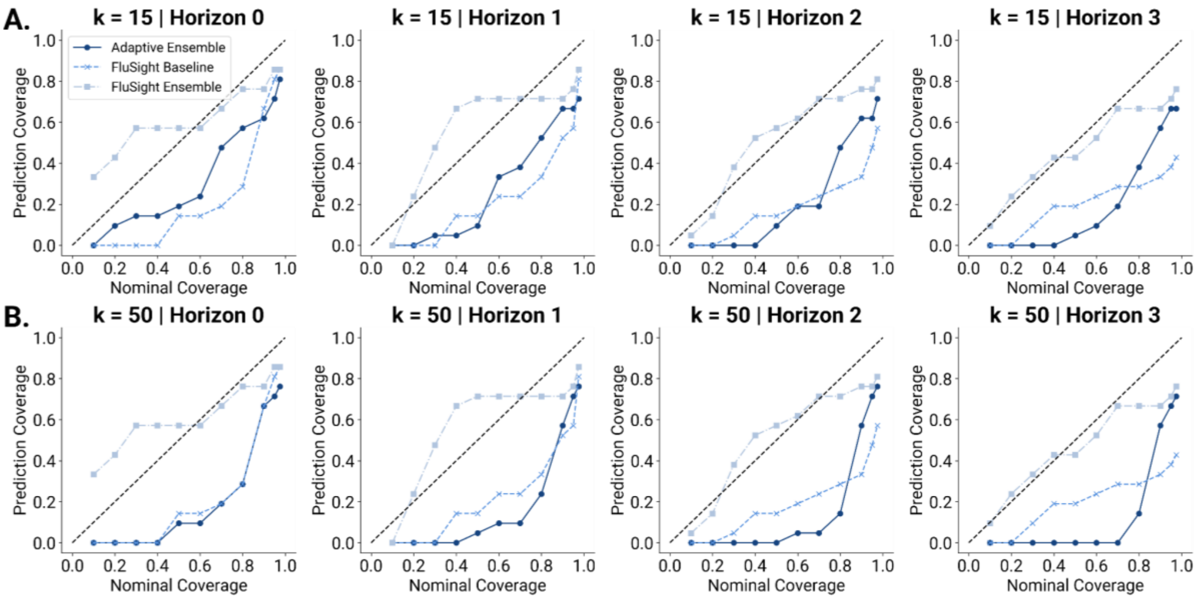


**Figure S28. Performance of the adaptive ensemble model in short-term forecasting (season 2024-2025).** A) coverage across different time horizons for the adaptive ensemble model, the FluSight baseline model, and the FluSight ensemble model, considering the top 15%. B) coverage across different time horizons for the adaptive ensemble model, the FluSight baseline model, and the FluSight ensemble model, considering the top 50%. Each data point represents the coverage value for a specific alpha level (0.10, 0.20, 0.30, 0.40, 0.50, 0.60, 0.70, 0.80, 0.90, 0.95, 0.975). The black dashed diagonal serves as a reference, indicating the coverage of an ideally calibrated model.

#### S6.2 Forecasting performance for 2023-2024 season

We repeated the analysis for the 2023-2024 season, with forecasts generated between October 14, 2023, and April 20, 2024, aligning with the rounds of the FluSight forecasting challenge. As before, forecasts produced by the adaptive ensemble approach were compared to both the FluSight ensemble model, an aggregate of all eligible contributed models, and the FluSight baseline model. Overall, the adaptive ensemble shows similar results to the ones obtained with the 2024-2025 season in terms of WIS aggregated across horizons (Fig. S29A). However, MAE ratios show slightly different results (Fig. S29B), consistently falling below 1 across all percentages when compared to both the FluSight baseline and FluSight ensemble models. Table S5 presents the median and interquartile ranges of WIS and MAE ratios for all percentages of trajectories selected, relative to the FluSight baseline and ensemble models. This contrasts with the 2024-2025 season, where the adaptive ensemble underperformed relative to the FluSight ensemble. In the 2023-2024 season, the adaptive ensemble demonstrated competitive performance to the FluSight ensemble for lower percentage of selected trajectories for WIS, and across all percentages for MAE. A breakdown of performance by forecast horizon (Figs. S30-S31) further supports the 2024-2025 pattern: the adaptive ensemble generally performs better at longer horizons in terms of median scores, outperforming both the FluSight baseline and FluSight ensemble models. Additionally, when visualizing the one-to-four-week-ahead forecasts across the three percentage configurations (Figs. S32-S33), we observe notable differences compared to the 2024-2025 season. Specifically, the adaptive ensemble in 2023-2024 shows reduced underprediction of incident hospitalizations. This is further reflected in the coverage metrics: the adaptive ensemble appears underconfident during the 2023-2024 season (Figs. S34-S35), whereas it exhibits an overconfidence tendency in 2024-2025 (Fig. S26). Similar to the 2024-2025 season, the FluSight ensemble consistently shows coverage rates that are closely aligned with nominal values. Notably, when examining the mismatch for 50% and 90% coverage levels (Table S6), the FluSight ensemble demonstrates the smallest deviations across most of the configurations.

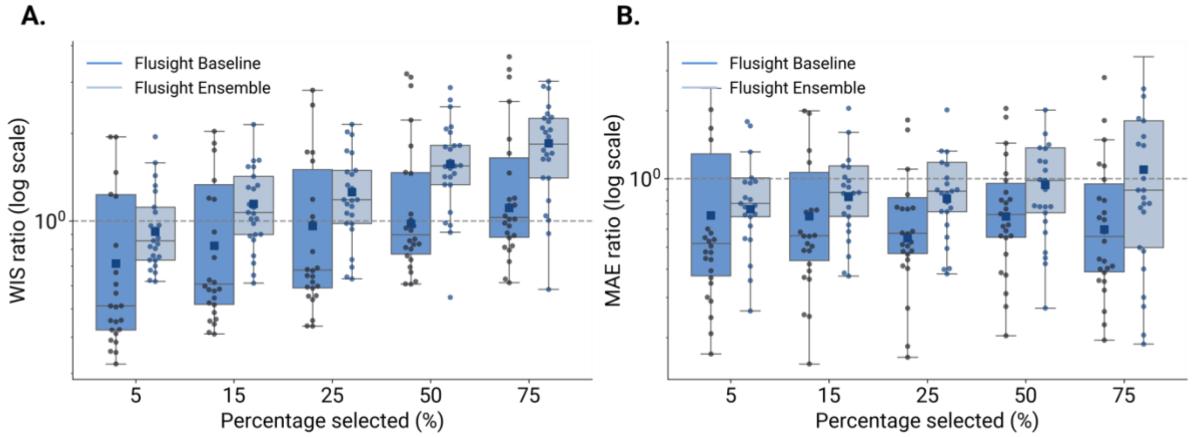


**Figure S29. Performance of the adaptive ensemble model in short-term forecasting, data from FluSight season 2023-2024.** A) Ratio (log-scale) between the WIS of the adaptive ensemble and FluSight Baseline or

FluSight Ensemble model across different percentages of trajectory selected and over all weeks. B) Ratio between the MAE of the median of the adaptive ensemble and FluSight Baseline or FluSight Ensemble model across different percentages of trajectory selected and over all weeks. A ratio below 1 indicates better performance of the adaptive ensemble with respect to the FluSight Baseline or FluSight Ensemble. Each boxplot is based on 30 data points, corresponding to the weeks in which the adaptive ensemble was generated. The overlaid swarmplot points correspond to each week. The boxplot boundaries represent the interquartile range (IQR) between the first quartile (Q1) and third quartile (Q3), and the line inside each box indicates the median. The whiskers extend to the furthest data point within 1.5 times the IQR from Q1 and Q3.

| **Percentage** *% of selected trajectories* | **WIS** **(Adaptive / FluSight Bas.)** *Median (50% CI)* | **WIS** **(Adaptive / FluSight Ens.)** *Median (50% CI)* | **MAE** **(Adaptive / FluSight Bas.)** *Median (50% CI)* | **MAE** **(Adaptive / FluSight Ens.)** *Median (50% CI)* |
|---|---|---|---|---|
| 5% | **0.51 (0.42-1.22)** | **0.86 (0.73-1.11)** | **0.50 (0.37-0.70)** | **0.78 (0.68-1.01)** |
| 15% | 0.60 (0.51-1.20) | *1.07 (0.90-1.42)* | 0.56 (0.42-0.73) | 0.87 (0.68-1.14) |
| 25% | 0.68 (0.59-1.19) | *1.18 (0.98-1.49)* | 0.57 (0.46-0.75) | 0.88 (0.72-1.18) |
| 50% | 0.87 (0.76-1.46) | *1.55 (1.33-1.82)* | 0.69 (0.55-0.87) | 0.98 (0.71-1.37) |
| 75% | *1.02 (0.88-1.46)* | *1.84 (1.41-2.25)* | 0.58 (0.39-0.99) | 0.89 (0.50-1.80) |

**Table S5: Performance summary of adaptive ensemble in short-term forecasting, data from FluSight season 2023-2024.** The table reports the median and interquartile ranges of WIS and MAE ratios for each percentage of trajectory selected. In bold, the best WIS and MAE ratio median values of the selected percentages; in italics, WIS and MAE ratio median values higher than 1. Values below 1 indicate better performance of the adaptive ensemble with respect to the reference (FluSight Baseline or Ensemble).

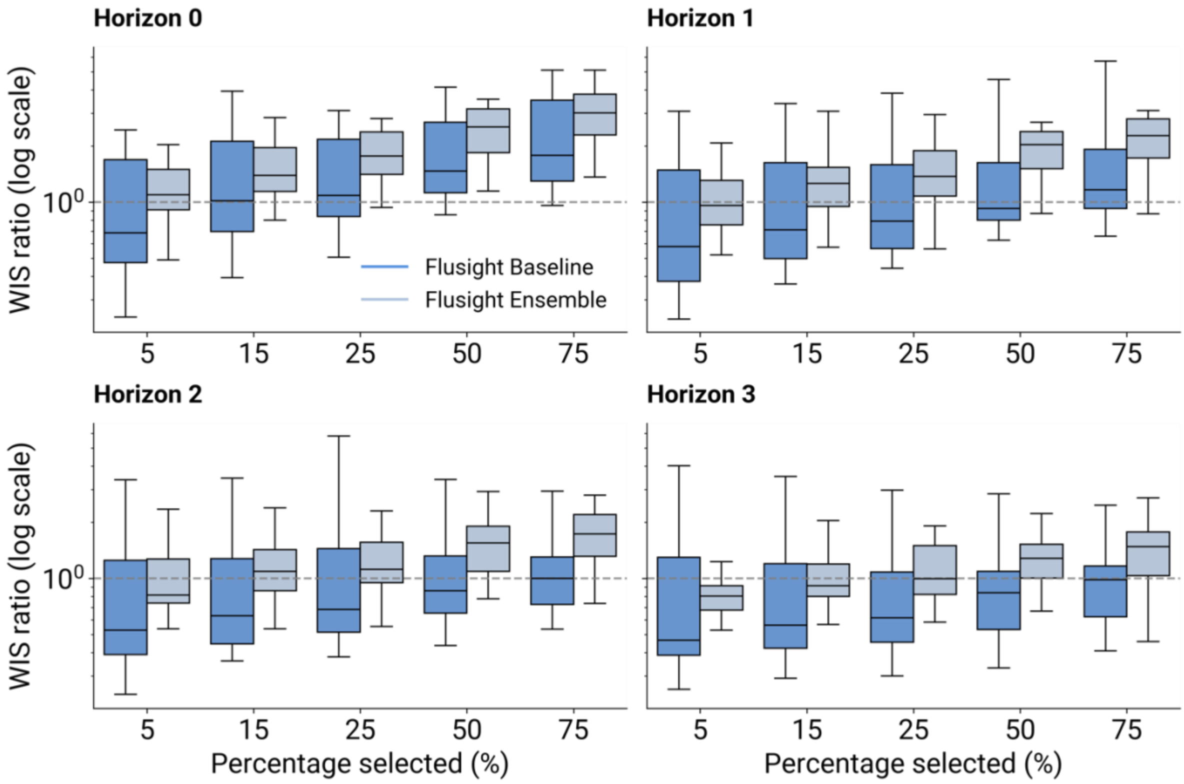


**Figure S30. Performance of the adaptive ensemble model in short-term forecasting (season 2023-2024).** Ratio (log-scale) between the WIS of the adaptive ensemble and the FluSight Baseline or FluSight ensemble models, across percentages of trajectory selection and for the four horizons separately. A ratio below 1 indicates that the adaptive ensemble outperformed the FluSight reference model. The boxplot boundaries represent the interquartile range (IQR) between the first quartile (Q1) and third quartile (Q3), and the line inside each box indicates the median. The whiskers extend to the furthest data point within 1.5 times the IQR from Q1 and Q3. Data points outside 1.5 times the interquartile range, above the upper quartile and below the lower quartile (Q1– 1.5 * IQR or Q3 + 1.5 * IQR), are considered outliers and not displayed.

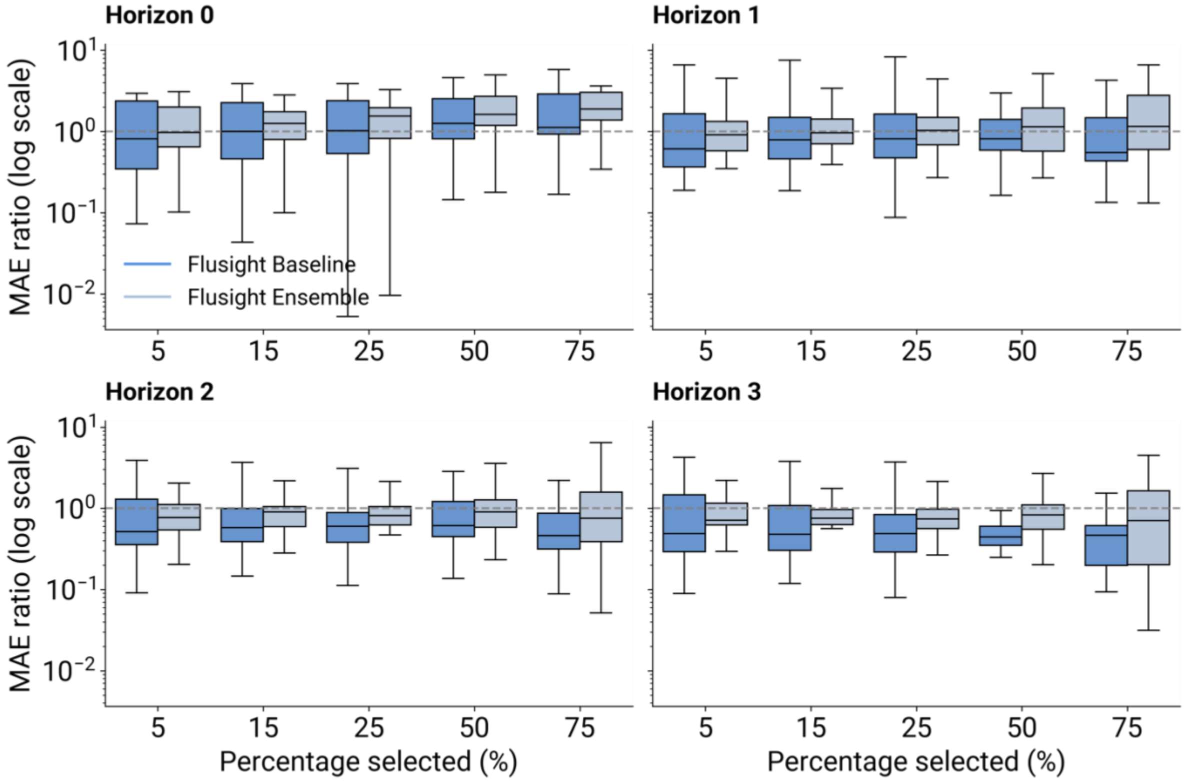


**Figure S31. Performance of the adaptive ensemble model in short-term forecasting (season 2023-2024).** Ratio (log-scale) between the MAE of the adaptive ensemble and the FluSight Baseline or FluSight ensemble models, across percentages of trajectory selection and for the four horizons separately. A ratio below 1 indicates that the adaptive ensemble outperformed the FluSight reference model. The boxplot boundaries represent the interquartile range (IQR) between the first quartile (Q1) and third quartile (Q3), and the line inside each box indicates the median. The whiskers extend to the furthest data point within 1.5 times the IQR from Q1 and Q3. Data points outside 1.5 times the interquartile range, above the upper quartile and below the lower quartile (Q1– 1.5 * IQR or Q3 + 1.5 * IQR), are considered outliers and not displayed.

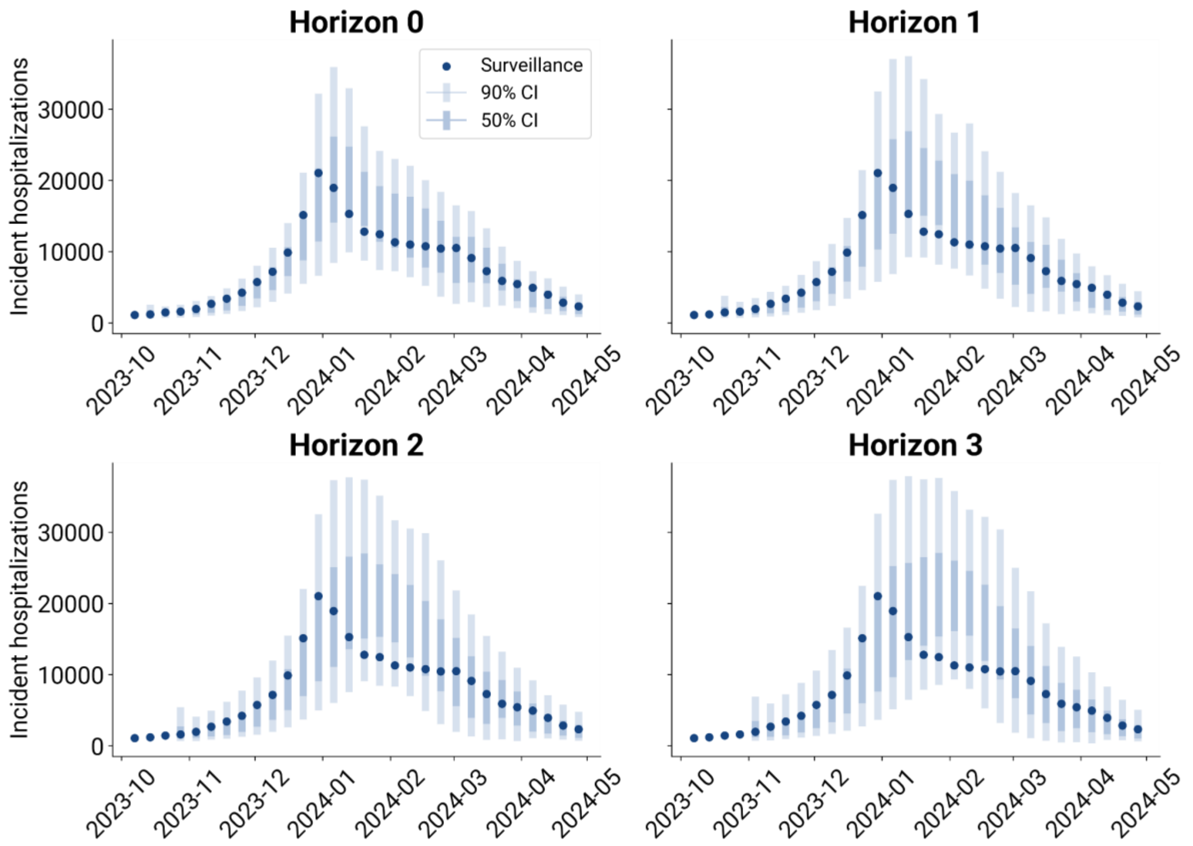


**Figure S32. Adaptive ensemble one- to four-week-ahead forecast (25% top trajectories, season 2023-2024).** The plots show one- to four-week-ahead forecasts, presenting the median, 50%, and 90% prediction intervals. These results are based on the adaptive ensemble using the top 25% of trajectories selected.

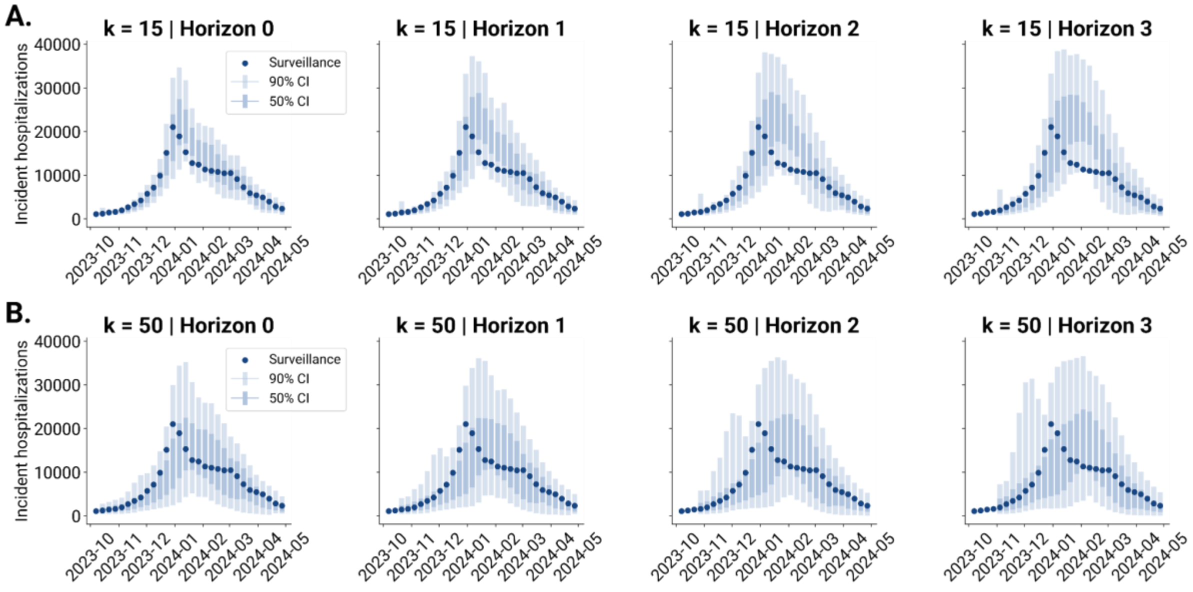


**Figure S33. Adaptive ensemble one- to four-week-ahead forecast (season 2023-2024).** The plot shows the one- to four-week-ahead forecasts, presenting the median, 50%, and 90% prediction intervals considering the 15% top trajectories for the adaptive ensemble generation (A) and the top 50% trajectories (B).

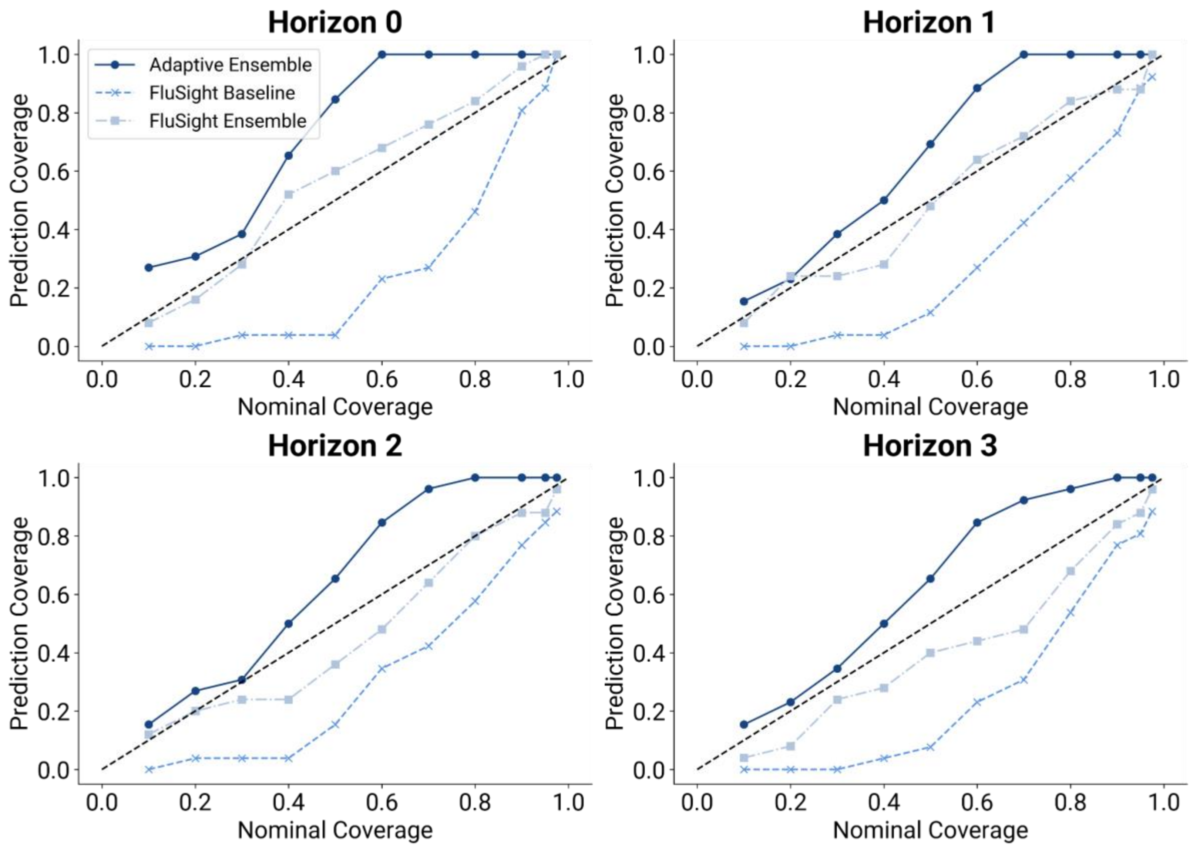


**Figure S34. Performance of the adaptive ensemble model in short-term forecasting (season 2023-2024).** The figure shows the coverage across different time horizons for the adaptive ensemble model, the FluSight baseline model, and the FluSight ensemble model. The adaptive ensemble has been generated using the 25% top trajectories. Each data point represents the coverage value for a specific alpha level (0.10, 0.20, 0.30, 0.40, 0.50, 0.60, 0.70, 0.80, 0.90, 0.95, 0.975). The black dashed diagonal serves as a reference, indicating the coverage of an ideally calibrated model.

| Metric | Horizon | Adaptive Ensemble | FluSight Baseline | FluSight Ensemble |
|---|---|---|---|---|
| *Coverage mismatch alpha = 0.5* | 0 | 0.35 | 0.46 | **0.10** |
| | 1 | 0.19 | 0.38 | **0.02** |
| | 2 | 0.15 | 0.35 | **0.14** |
| | 3 | 0.15 | 0.42 | **0.10** |
| *Coverage mismatch alpha = 0.9* | 0 | 0.10 | 0.09 | **0.06** |
| | 1 | 0.10 | 0.17 | **0.02** |
| | 2 | 0.10 | 0.13 | **0.02** |
| | 3 | 0.10 | 0.13 | **0.06** |

**Table S6: Coverage mismatch summary of adaptive ensemble in short-term forecasting (season 2023-2024).** The table presents the absolute differences between nominal and predicted coverage values (coverage mismatch) for alpha levels 0.5 and 0.9, across four time horizons. Results are reported for the adaptive ensemble, FluSight baseline, and FluSight ensemble models. The best-performing values for each alpha level and horizon are highlighted in bold.

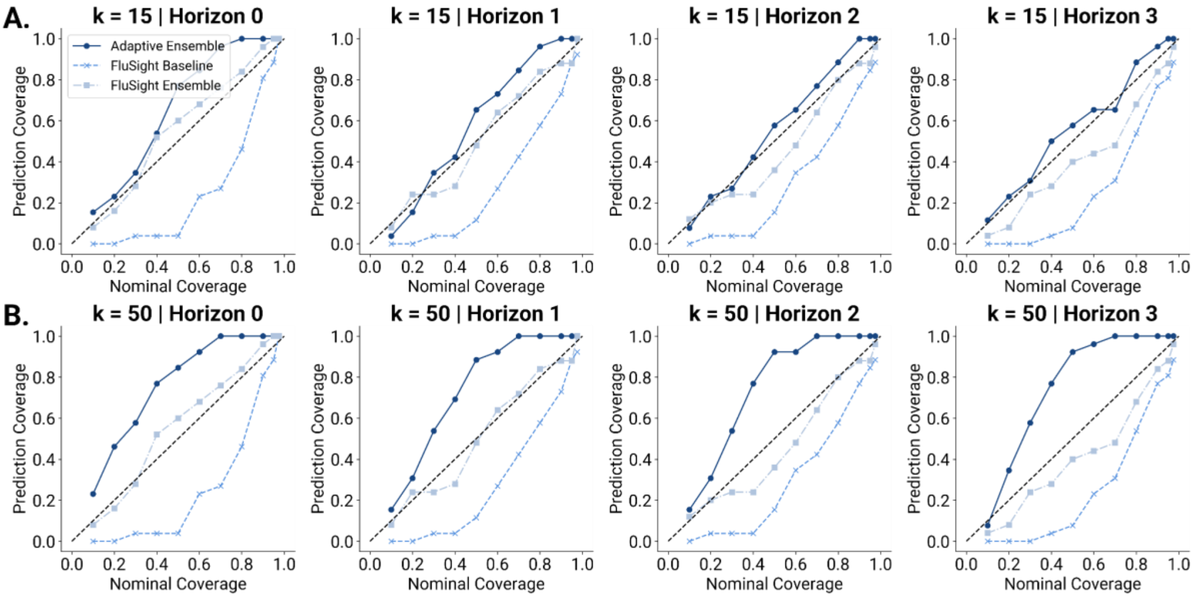


**Figure S35. Performance of the adaptive ensemble model in short-term forecasting (season 2023-2024).** A) coverage across different time horizons for the adaptive ensemble model, the FluSight baseline model, and the FluSight ensemble model, considering the top 15%. B) coverage across different time horizons for the adaptive ensemble model, the FluSight baseline model, and the FluSight ensemble model, considering the top 50%. Each data point represents the coverage value for a specific alpha level (0.10, 0.20, 0.30, 0.40, 0.50, 0.60, 0.70, 0.80, 0.90, 0.95, 0.975). The black dashed diagonal serves as a reference, indicating the coverage of an ideally calibrated model.
